# A Multimodal Vision-text AI Copilot for Brain Disease Diagnosis and Medical Imaging

**DOI:** 10.1101/2025.01.09.25320293

**Authors:** Guoxun Zhang, Zebin Gao, Caohui Duan, Jiaxin Liu, Yuerong Lizhu, Yaou Liu, Qian Chen, Ling Wang, Kailun Fei, Tianyun Wang, YuJia Chen, Yanchen Guo, Yuchen Guo, Xin Lou, Qionghai Dai

## Abstract

Integrating non-invasive brain imaging techniques, particularly computed tomography (CT) and magnetic resonance imaging (MRI), coupled with the advancement of artificial intelligence, is forging a key pathway for brain disease diagnosis, playing a vital role in safeguarding human health*^1–4^*. A robust artificial intelligence copilot is essential for clinical emergencies, functioning as the central processing unit for brain medical imaging systems, aiming to revolutionize the imaging process, expedite the diagnosis of diseases, and support treatment*^5–7^*. In this study, we developed an advanced multi-modal brain medical imaging foundational model named Brainfound, utilizing AI-generated content and image-text alignment technology, pre-trained on over 3 million brain CT images and over 7 million brain MRI images with their paired reports. As a clinical brain medical imaging multi-modal model, Brainfound achieved state of the art on seven downstream tasks, including brain disease diagnosis, brain lesion segmentation, MRI image enhancement, MRI cross-modality translation, automatic report generation, zero-shot brain disease classification, and free human-AI conversation. After thorough human-machine validation, Brainfound surpassed the current leading model by 51.75% in automatic report generation for brain imaging. In multiple-choice questions related to brain imaging, the accuracy of Brainfound outstripped GPT-4V by 47.68%, comparable to experienced doctors. We anticipate Brainfound, a clinical model with flexible visual and text input-output capabilities, will provide substantial support in brain medical imaging, clinical education, and human-in-the-loop medical diagnosis.

## Introduction

Brain computed tomography (CT) and magnetic resonance imaging (MRI) have become essential non-invasive diagnostic tools in clinical scenarios. These imaging studies are critical for accurately diagnosing various brain conditions, including tumors, strokes, and neurodegenerative diseases^8–11^. CT is renowned for its rapid assessment capabilities, especially in acute settings, providing more reliable detection of hemorrhagic stroke. In contrast, MRI excels at producing high-resolution images of soft tissues, significantly increasing early detection rates for brain tumors and neurodegenerative changes. Collectively, these advancements highlight the significance of brain imaging in safeguarding patient health and facilitating timely medical interventions^12–14^.

Artificial intelligence (AI) plays a multifaceted role in augmenting the capabilities of brain CT and MRI^15,16^. AI technologies enable automated image analysis, rapidly identifying abnormalities such as lesions, tumors, and hemorrhages, thereby significantly enhancing diagnostic efficiency^17–20^. Deep learning models improve diagnostic accuracy by recognizing complex image features, sometimes matching or even surpassing the assessments of expert radiologists^21,22^. Additionally, AI supports clinical decision-making by integrating the clinical and imaging data of patients and providing personalized treatment recommendations^23,24^. Predictive analytics further enhance the early identification of high-risk patients and potential disease progression. Overall, AI optimizes resource allocation, ensuring that high-risk cases receive prompt and effective care. These advancements highlight the critical role of AI in improving diagnostic accuracy, clinical decision-making, and patient outcomes in neuroimaging^25^.

The challenges posed by limited labeled data and the complexities of acquiring multi-modal annotations significantly impact the development of AI models^26^. In medical imaging, the scarcity of high-quality labeled data hampers training efficacy, preventing models from achieving optimal performance^27,28^. The annotating of multi-modal data, such as the combination of brain CT and MRI, necessitates advanced expertise, leading to increased time and costs associated with data preparation. This complexity not only heightens the risk of human error but also limits the diversity and richness of the training datasets, restricting the capacity of the model for generalization to unseen data. Additionally, the issue of data imbalance, where common conditions often overshadow rare diseases, exacerbates the risk of overfitting and diminishes overall model robustness. Addressing these challenges necessitates innovative methodologies, including semi-supervised and transfer learning approaches, to enhance model training and performance despite data limitations^29,30^. Such strategies are essential for ensuring that AI applications in clinical practice remain effective and reliable.

Large AI models have emerged as a promising solution to these challenges. Inspired by breakthroughs in large language and vision models like ChatGPT^31,32^, CLIP^33^, SimCLR^34^, and DINO^35^, medical foundational models are thriving and pushing the frontiers in computational pathology^36^, ophthalmic disease diagnosis^37^, ultrasonography^38^, and cancer biomarker innovation^2^. These advancements bolster diagnostic accuracy, facilitate knowledge sharing, and advance medical education^39^. By leveraging large-scale pre-training on diverse, unannotated datasets, foundation models capture robust feature representations, enabling effective performance even when labeled data is limited^6,40^. Their capability to integrate multimodal information further enhances their reliability in clinical applications. In addition, foundational models support advanced techniques such as self-supervised learning and generative models, which can synthesize annotated data, thereby expanding the training datasets and improving model generalization. Consequently, the introduction of foundation models not only provides innovative strategies for overcoming the challenges posed by the scarcity of annotated data and the integration of multimodal information but also opens new avenues for enhancing CT and MRI image analysis to identify brain diseases.

In this study, we introduce Brainfound, a multimodal AI copilot for brain medical imaging based on the AIGC and image-text alignment technology (Fig. 1), which has been pre-trained on BrainCT-3M and BrainMRI-7M (Supplementary Fig. 1, Supplementary Fig. 2) collected from the Chinese PLA General Hospital to achieve the aforementioned purpose. BrainCT-3M is a massive brain CT scan dataset containing 107,754 brain CT scans and corresponding diagnostic reports, totaling over 3 million images. BrainMRI-7M is a brain multi-sequence MRI scan dataset containing 68,653 brain multi-sequence MRI scans and corresponding diagnostic reports, totaling over 7 million images. By harnessing these two datasets, we pre-train image encoders and decoders based on the Denoising Diffusion Probabilistic Model (DDPM, Fig. 1b) strategy, as well as text decoders based on the LLaMa^41^ framework (Fig. 1b). We aligned the visual module and language module of Brainfound, which comprehended brain medical imaging and knowledge during pre-training, empowering Brainfound to tackle diverse and flexible downstream tasks. In comparison with multimodal models like GPT-4V, Brainfound achieved the best performance across seven types of tasks, including brain disease diagnosis, brain lesion segmentation, MRI image enhancement, MRI cross-modality translation, automatic generation of reports from images, zero-shot brain disease classification, and free human-AI conversation. Especially in two tasks: one is in automatic report generation, where Brainfound scored about 50% higher than the current top model in human-machine evaluation; the other is in multiple-choice question answering, where Brainfound outperformed GPT-4V by 47.68% in accuracy, comparable to experienced doctors.

**Fig. 1.**
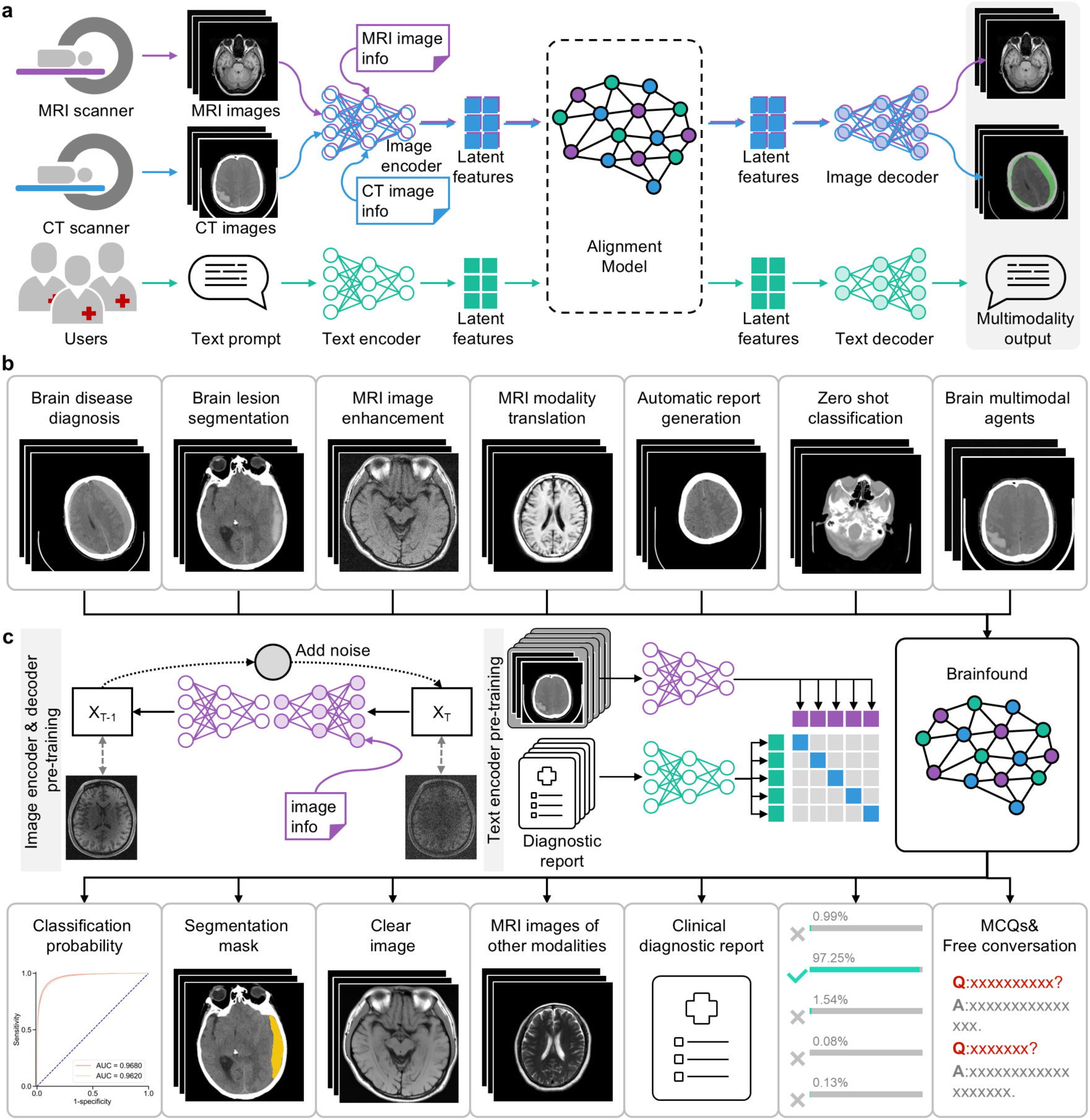
Overview of Brainfound. **a**, Brainfound aims to develop an AI medical copilot capable of processing brain CT or MRI scan sequences and user instructions as inputs and delivering either processed images or textual outputs. CT or MRI image sequences and the basic information of the images are encoded by the image encoder to obtain latent space features, which are aligned with the features produced by the text encoder in the alignment model. Then, the alignment model is connected to two decoders to obtain the output of Brainfound. **b**, Downstream task evaluation for Brainfound. As an AI medical copilot, Brainfound excels in several downstream tasks, including brain disease diagnosis, brain lesion segmentation, MRI image enhancement, MRI cross-modality translation, automatic generation of reports from images, zero-shot brain disease classification, and free human-AI conversation. These tasks include all tasks related to brain medical imaging, from single-modality pixel-level tasks to multi-modality diagnostic-related conversations. **c**, Stepwise and modular pre-training strategy for Brainfound. The image encoder and decoder of Brainfound undergo pre-training through the DDPM strategy on the BrainCT-3M and BrainMRI-7M, with fundamental information like image modality randomly masked and then input into the image encoder and decoder. The text encoder is pre-trained using the CLIP strategy, aligning paired diagnostic reports and image sequences in the latent feature space.

## Results

### Brainfound serving as a multimodal AI copilot for brain medical imaging

Large-scale datasets are essential for building robust AI models. We assembled a comprehensive national Brain medical imaging dataset, to our knowledge, this is the largest multimodal brain medical imaging dataset available, containing BrainCT-3M and BrainMRI-7M collected from the Chinese PLA General Hospital. BrainCT-3M originates from a larger multicenter dataset, which catalogs 630,992 scans (enrolled between 2008 and 2022, comprising 366,123 males and 264,869 females) (Supplementary Fig. 1). The dataset was refined based on image quality, specifically the level of signal-to-noise ratio (SNR) and the diagnostic details provided in the reports (Methods), resulting in 105,184 CT scans (59,935 males and 45,249 females) and paired diagnostic reports. This refined dataset includes approximately 46,066 cases of normal individuals, 25,197 cases of ischemia, 20,798 cases of hemorrhage, 19,497 cases of fractures, and 3,282 cases of tumors. Additionally, we gathered 68,653 MRI scans and paired diagnostic reports, including 36,002 males and 32,651 females, with admission dates from 2018 to 2023, covering ages from 1 to 105 years (Supplementary Fig. 2). The BrainMRI-7M encompasses several primary modalities: T1-weighted imaging (T1WI), T2-weighted imaging (T2WI), Diffusion-weighted imaging (DWI, low-b-value and standard-b-value), Fluid attenuated inversion recovery (FLAIR), etc. We employed the GPT-4 API to automatically tokenize MRI reports, allowing us to count the image types and quantities involved in BrainMRI-7M. The statistical results revealed the top 10 most frequent terms: Ischemia 32,021 times, Softening Focus 6,574 times, Normal 4,273 times, Inflammation 3,936 times, Cyst 3,565 times, Atherosclerosis 3,035 times, Senile Brain Changes 2,865 times, Cerebral Infarction 2,751 times, Hemorrhage 1,934 times, Vascular Stenosis 1,498 times (Supplementary Fig. 2). Meanwhile, we collected a wide range of data for downstream validation tasks. These include publicly available datasets and hospital-acquired data. The publicly available datasets include the RSNA Intracranial Hemorrhage Classification dataset^42^ for validating classification tasks and the BraTS MRI^8^ dataset for image quality enhancement. The hospital-acquired data consists of physician-annotated intracranial hemorrhage segmentation data, midline shift segmentation data, MCQ data, report generation data, zero-shot classification data from both internal and external centers, and MRI data for modality conversion.

In the training phase of the image encoder and decoder (Brainfound-v), we utilized a U-net architecture augmented by transformer blocks featuring cross-attention^43^ mechanisms, comprising approximately 78 million trainable parameters (Supplementary Fig. 3, Methods). Paired clinical information, such as image modality, was randomly selected and encoded using BERT^44^, then integrated with corresponding brain images to serve as input for the primary network architecture of Brainfound-v. The self-supervised pre-training of Brainfound-v is anchored on the fundamental principles of DDPM^45^(Methods). A two-dimensional Brain CT or MRI image, along with its basic information, is randomly selected from the BrainCT-3M and BrainMRI-7M datasets and subjected to data augmentation. In the course of forward propagation, Gaussian noise with a defined intensity is systematically introduced into the image. Upon reaching 1000 iterations, this procedure culminates in the conversion of the image into pure noise (Fig. 1b, Supplementary Fig. 4a). Throughout backward propagation, the neural network meticulously learns to perform denoising and reconstructs the clean image (Supplementary Fig. 5), thereby cultivating advanced representation learning capabilities (Supplementary Fig. 6). Modality information from brain CT or MRI images serves as guidance and is concurrently input into the model. To bolster the robustness of the main backbone network against this modality information, we randomly occlude portions or the entirety of it as input. In the process of report generation, we employed the text encoder and decoder of BERT (102 million trainable parameters) and initialized them with pre-trained weights, along with diagnostic reports to pre-train the text encoder and decoder (Supplementary Fig. 7, Methods). For the task of open conversation, we utilized a LLaMA-like model (8 billion parameters) and its weights for initialization, using Lora^46^ for fine-tuning with our self-collected instruction datasets about brain medical imaging (Supplementary Fig. 8, Methods).

During the inference phase, we design task-specific adapters to fully harness the capabilities of Brainfound. For the cerebral hemorrhage classification task, a trainable multilayer perceptron (MLP) is utilized to convert the final output features of the Brainfound image encoder into diagnostic labels (Supplementary Fig. 4b, Methods). In the segmentation tasks for cerebral hemorrhage and midline shift, a set of learnable MLP classifiers is employed to independently classify the intermediate features extracted by the image encoder-decoder (Supplementary Fig. 4c, Methods). For the modality transfer task, the diffusion model is fine-tuned using the image encoder-decoder with modality-conditioned images. In the denoising task, we develop a zero-shot learning denoising diffusion model based on the image encoder-decoder module. For contrastive learning, a set of aggregation modules is introduced to merge the features from the image encoder and achieve scan-level alignment with reports (Supplementary Fig. 7, Methods). Finally, in the construction of the AI assistant, the features extracted by the image encoder are transformed into tokens and integrated with text inputs into a LLaMa-like large language model for unified processing (Supplementary Fig. 8, Methods).

### Brainfound performs precise diagnosis and localization of brain diseases

As a potentially life-threatening condition, intracranial hemorrhage requires an accurate diagnosis for proper treatment. We compared the full parameter fine-tuning performance and only the tail MLP fine-tuning performance of Brainfound with other methods on the publicly available RSNA intracranial hemorrhage classification dataset. We used 222,218 images from the RSNA intracranial hemorrhage dataset, with half used for model training and the other half for model testing. We conducted four experiments. The first experiment involved full parameter fine-tuning of models with different amounts of training data to compare the accuracy of intracranial hemorrhage classification. We compared Brainfound, MAE pre-trained on natural images (MAE)^47^, MAE pre-trained on medical images (MAE pre-trained), and models pre-trained on RadImagenet^48^. The models underwent full fine-tuning using the entire training set (about 110,000 images) and reduced portions of the training set (1/2, 1/4, 1/8, 1/16, and 1/32). Brainfound achieved the best AUC across all training data volumes (Fig. 2a). For instance, with fine-tuning on the 1/32 training set (Fig. 2b, Supplementary Fig. 9a), Brainfound achieved an AUC of 0.8739 (95% CI 0.8690-0.8769), with ResNet^49^ in second place reaching an AUC of 0.8739 (95% CI 0.8690-0.8769). The second experiment focuses on fine-tuning just the tail MLP under varying training data volumes, with the pre-trained model serving as a feature extractor for brain CT images. Brainfound achieved the highest AUC across all training data volumes (Fig. 2c). For instance, with 1/32 of the training set (Fig. 2d, Supplementary Fig. 9b), Brainfound achieved an AUC of 0.7776 (95% CI 0.7731-0.7817), whereas ResNet, in second place, reached an AUC of 0.7267 (95% CI 0.7215-0.7317). In addition, we incorporated the state-of-the-art method from the RSNA Brain Hemorrhage Classification competition to demonstrate the plug-and-play performance of Brainfound as the foundation model. This method is based on an ensemble learning approach with three different backbones. During comparison, we replaced one of the backbones (Densenet121^50^) with Brainfound. In all experiments, models based on Brainfound achieved the highest AUC scores (Supplementary Fig. 9c-d).

**Fig. 2.**
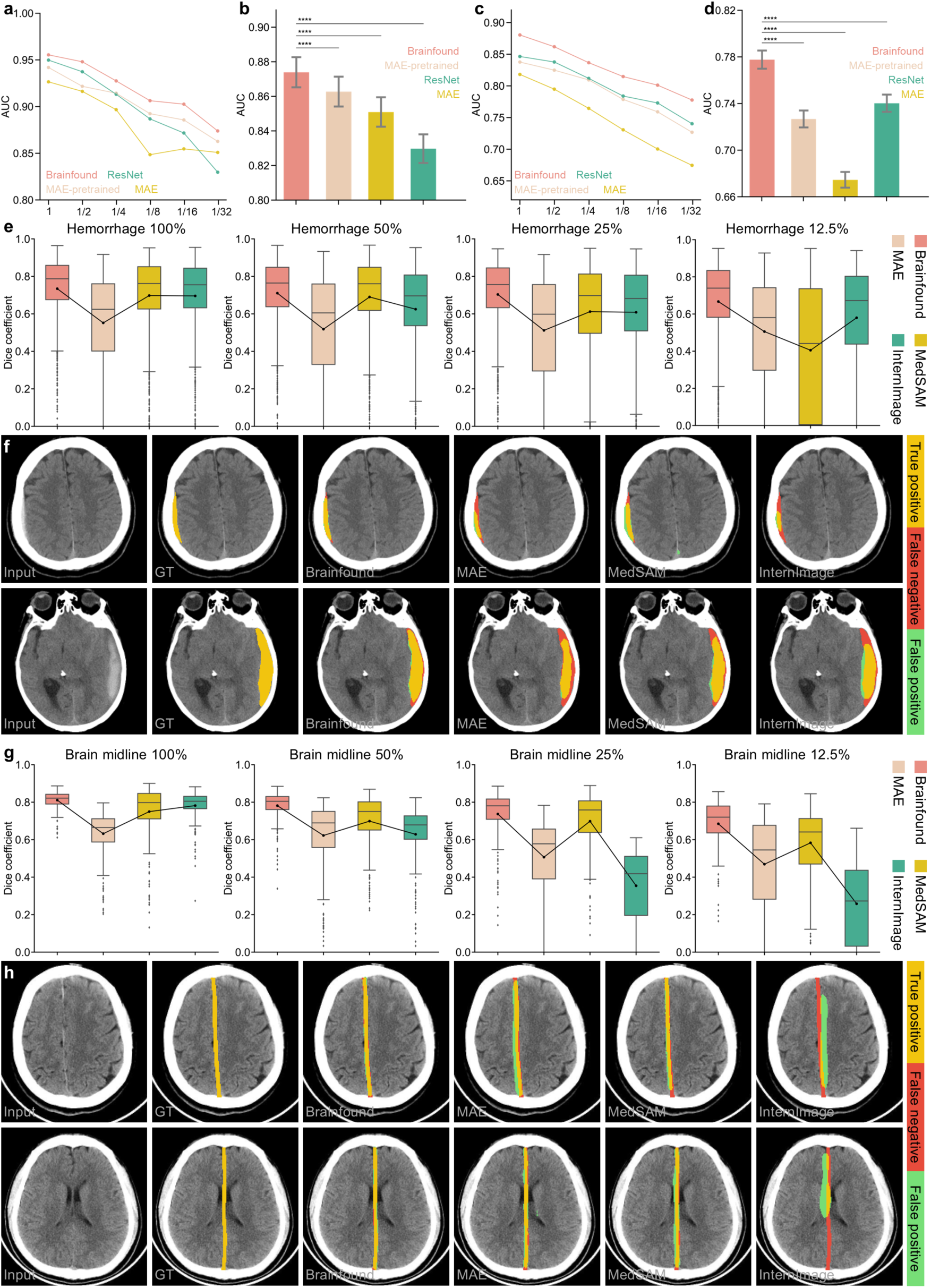
Performance of Brainfound in diagnosing and localizing brain diseases. **a,** The AUC results for four methods in full parameter fine-tuning for brain hemorrhage classification. “1” denotes fine-tuning using the complete training set (110,000 training images), “1/2” signifies using half of the dataset, and so forth. The four methods include Brainfound, ResNet, MAE-pretrained, and MAE. **b**, The AUC results for the four methods using 1/32 of the training set for full parameter fine-tuning in brain hemorrhage classification. **c**, The AUC results for four methods when only the final MLP is fine-tuned for brain hemorrhage classification. The four networks serve as feature extractors, with parameters frozen during fine-tuning. **d**, The AUC results for four methods using 1/32 of the training set with only the final MLP fine-tuned for brain hemorrhage classification. **e**, Comparing the accuracy of four methods with full parameter fine-tuning on brain hemorrhage segmentation using different training set sizes. “100%” indicates using the entire training set (220 brain CT scans) to fine-tune the four methods. The four methods are Brainfound, MedSAM, MAE, and InternImage. **f**, Two cases of brain hemorrhage segmentation. From left to right are the original CT image, ground truth, and segmentation results from Brainfound, MedSAM, MAE, and InternImage. **g**, The accuracy comparison of four approaches with full parameter fine-tuning for midline brain segmentation across varying training set sizes. “100%” denotes using the complete training set (439 images) for fine-tuning. The methods compared include Brainfound, MedSAM, MAE, and InternImage. **h**, Two cases of midline brain segmentation. From left to right are the original CT image, ground truth, and segmentation results by Brainfound, MedSAM, MAE, and InternImage.

The visual module of Brainfound is built upon DDPM, a robust image generation tool that effectively learns the prior knowledge needed for dense prediction. We evaluated this in brain hemorrhage segmentation and midline shift detection tasks. For the brain hemorrhage segmentation task, we gathered 2060 brain CT scans from the Chinese PLA General Hospital, with 220 cases in the training set (1397 images), 760 cases in the validation set (5440 images), and 1080 cases in the test set (7917 images). In the full parameter fine-tuning process for Brainfound and three other comparative methods, we established four groups of training data volumes, using 12.5%, 25%, 50%, and 100% of the training set data (Fig. 2e). In all segmentation tasks for intracerebral hemorrhages, Brainfound achieved the best performance (Fig. 2e). For example, when fine-tuning with only 12.5% of the training data, the Dice coefficient of Brainfound was 0.6671 (95% CI 0.6536-0.6805), surpassing the second-place InternImage by approximately 31.99%. Brainfound excels at segmenting small hemorrhages located near the skull (Fig. 2f). The shift in the brain midline is a reliable indicator of the severity of brain diseases. For the task of brain midline localization and segmentation, we collected data from 301 patients, including 12 cases in the training set (439 images), 50 cases in the validation set (1629 images), and 239 cases in the test set (7743 images). In the experiment, we tested four different training data volumes: 12.5% training set (1 case, 72 images), 25% training set (3 cases, 138 images), 50% training set (6 cases, 236 images), and 100% training set (12 cases, 439 images), to assess the performance of four models in few-shot learning (Fig. 2g). In the comparison with MedSAM^51^, MAE, and InternImage^52^, Brainfound outperformed the other models, achieving the highest Dice coefficient (Fig. 2g). The best performance was observed with 12.5% of the training data, where Brainfound scored 0.6848 (95% CI 0.6682-0.7014), exceeding the second-place model by 17.47% (Fig. 2g). With a pixel-level visual model, Brainfound delivers more accurate masks in brain midline segmentation (Fig. 2h). From the results of the saliency map analysis, it is evident that the attention of Brainfound effectively concentrated on the necessary hemorrhage area or midline region for the task (Supplementary Fig. 10-12).

### Brainfound boosts the imaging capability of brain medical equipment

MRI is a fundamental tool in clinical neuroimaging applications. Unlike CT, MRI is non-invasive, non-ionizing, inherently quantitative, and multi-parametric. By utilizing strong magnetic fields and radiofrequency signals, MRI generates high-contrast images of soft tissues, which makes it particularly advantageous in detecting brain conditions such as tumors, inflammation, and vascular abnormalities. From the standpoint of the main magnetic field strength, MRI covers a range from low-field to high-field and even ultra-high-field. Low-field MRI (such as 0.3T) has the advantages of lower equipment and maintenance costs and higher patient comfort, but the acquired images have lower spatial resolution and image quality, with longer acquisition times^53–55^. High-field MRI (such as 3T) provides higher signal-to-noise ratio (SNR) and spatial resolution with shorter scan times. However, it is expensive and has limited availability. 5T MRI is gradually entering clinical practice and is helping to solve many clinical problems by improving resolution^56^. From the standpoint of acquisition sequences, MRI allows for the acquisition of high-resolution brain images with different contrasts. For MRI imaging, reducing scanning time would greatly expand its clinical application scenarios. Along with the rapid development of AI-enhanced MRI imaging, current AI methods can both recover clear images from low signal-to-noise ratio MRI images taken with fast acquisition and enable virtual multi-modal MRI imaging through modality transformation. The visual module of Brainfound, based on DDPM, has an inherent advantage in pixel-level tasks, as we have validated its performance in zero-shot learning for MRI denoising and few-shot learning for modality translation.

We first utilized the publicly available 3T MRI high SNR image dataset: the Brain Tumor Segmentation (BraTS) Challenge 2023, randomly selecting 10 brain T1WI scans (a total of 1380 images) to construct a zero-shot learning test set. Different intensities of simulated Rican noise were added to the clear MRI images to produce six test sets with varying noise levels (Methods), with average image SNRs of 9.6808 dB, 11.7425 dB, 14.7979 dB, 15.6190 dB, 16.5326 dB, and 17.5389 dB. We constructed a zero-shot learning iterative denoising architecture using Brainfound (Supplementary Fig. 13, Methods) and compared it with pre-trained SCUnet^57^, Neighbor2neighbor^58^ (Nei2nei), and Noise2self^59^. We assessed the quality of the enhanced images using PSNR, RMSE, SNR, and SSIM(Methods), which are commonly used metrics in computer vision. Brainfound achieved optimal scores on all four metrics across six different noise levels in the test sets (Fig. 3a-d). For instance, when the average SNR of test images is 14.7979 dB (95% CI 14.7171-14.8786 dB), the SNR of the images enhanced by Brainfound is 19.7580 dB (95% CI 19.6456-19.8704 dB), surpassing the second-place SCUnet by 6.89% (Fig. 3c), whose enhanced image SNR is 18.4848 dB (95% CI 18.3855-18.5842 dB). Brainfound manages to remove noise while retaining details (Supplementary Fig. 14). Moreover, we gathered paired MRI images with both high and low SNR from various sequences spanning low field (0.3T) to high field (5T) to assess the denoising capabilities of Brainfound. The 0.3T low-field MRI images were sourced from the M4Raw dataset, including T1WI (25 scans, 450 images), T2WI (25 scans, 450 images), and FLAIR (25 scans, 450 images). The 5T ultra-high field MRI images were collected at Beijing Friendship Hospital, including T2WI (1 scan, 10 images) and T1WI (1 scan, 19 images), and 25 images were collected from a 5T MRI at Shanghai United Imaging as the external test set (Methods). Brainfound secured the best scores on almost all datasets and metrics (Fig. 3e-h). For instance, in terms of PSNR, Brainfound surpassed the second place by up to 5% (Fig. 3e). Detailed comparisons and case studies are available in the supplementary materials (Supplementary Fig. 15-20).

**Fig. 3.**
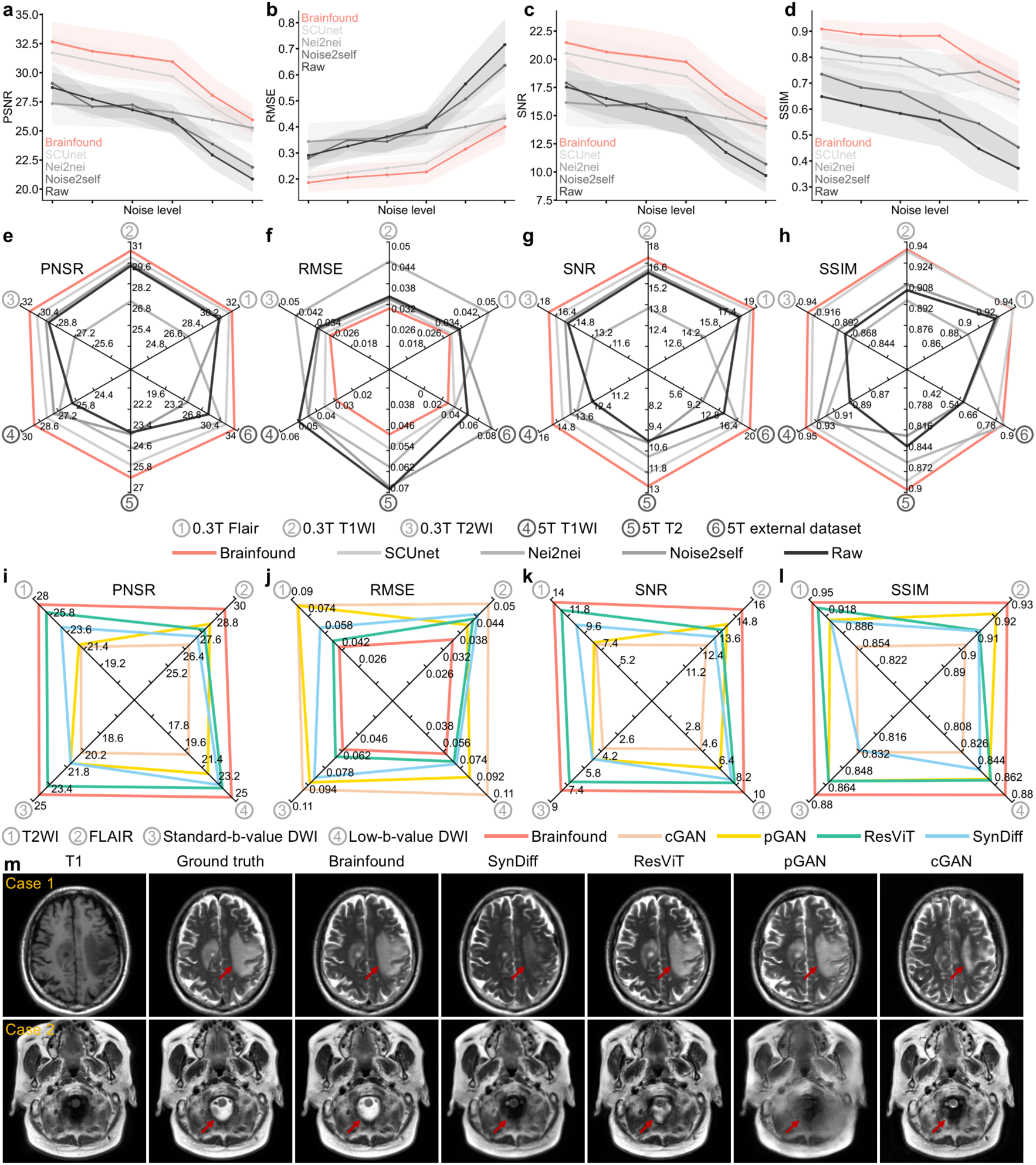
Estimation of Brainfound in MRI image enhancement and cross-modality translation. **a-d,** The zero-shot learning denoising performance of four methods was quantitatively assessed on the simulated dataset. PSNR, RMSE, SNR, and SSIM were calculated on the test dataset(n=1380). Simulated datasets with six different noise levels were employed to evaluate the image enhancement capabilities of the four methods. Different colored curves indicate the results of different methods. Brainfound consistently achieved superior denoising effects across all noise conditions. **e-f**, High SNR, and low SNR images were collected from the low-field MRI (0.3 T) and the ultra-high-field MRI (5 T) to validate the zero-shot learning denoising performance of four methods. The test set comprises 0.3 T FLAIR scans(n=450), 0.3 T T1WI scans(n=450), 0.3 T T2WI scans (n=450), 5 T T1WI scans(n=19), 5 T T2WI scans (n=10), and 5 T external test set (n=25). PSNR, RMSE, SNR, and SSIM were calculated on six real-world test datasets. Each radar chart represents the results of one metric. Different colored curves illustrate the denoising results of each method. Brainfound achieved the best scores on almost all metrics. **i-l**, Assessment of the cross-modality translation capability of five methods on clinically common sequences. The original sequence modality is T1WI, while the conversion target sequences include T2WI, FLAIR, low-b-value DWI, and standard-b-value DWI. The training set comprises 94 head 3T MRI scans (2205 images), and the test set includes 88 head 3T MRI scans (1936 images). Each radar chart represents the results of one metric. Different colored curves indicate the modality translation results of each method. Brainfound achieved the best scores on all metrics. **m**, two cases of T1WI to T2WI modality translation. In case 1, the lesion indicated by the red arrow is translated more accurately in the result of Brainfound. In case 2, the structures within the foramen magnum (indicated by the red arrow) are demonstrated in the result of Brainfound, with cerebrospinal fluid exhibiting high signal intensity, presenting distinct contrast against the skull, medulla oblongata, and vertebral arteries.

We then validated the ability of Brainfound in MRI image modality transformation on clinically common MRI sequences. We collected 182 cases of brain 3T MRI scan data from the Chinese PLA General Hospital, including T1WI, T2WI, FLAIR, low-b-value DWI, and standard-b-value DWI modalities. We used AI methods to virtually generate images of the other four modalities from T1WI. We used 94 scans (2205 images) as the training set and 88 scans (1936 images) as the test set. We compared Brainfound with SynDiff^60^, ResViT^61^, pGAN^62^, and cGAN^63^ in modality transformation, which covers the commonly used MRI modality transfer network structures and methods. We employed PSNR, RMSE, SNR, and SSIM to quantify the comparison of modality transformation results. Brainfound achieved the best results across four modality transformation tasks (Fig. 3i-l). In SNR comparisons (Fig. 3k), Brainfound showed improvements of 8.17% for T1WI to T2WI, 7.52% for T1WI to FLAIR, 12.23% for T1WI to standard-b-value DWI, and 11.17% for T1WI to low-b-value DWI over the second place. In PSNR comparisons (Fig. 3i), Brainfound showed improvements of 3.90% for T1WI to T2WI, 3.78% for T1WI to FLAIR, 4.17% for T1WI to standard-b-value DWI, and 3.40% for T1WI to low-b-value DWI over the second place. The T1WI to T2WI modality transformation results show that the output of Brainfound more clearly generates lesion areas and highlights important structural regions (Fig. 3j). Additional examples of modality conversion are available in the supplementary materials (Supplementary Fig. 21-24).

### Brainfound autonomously drafts high-quality clinical reports

Automatically generating brain imaging reports has the potential to improve the medical experience of patients and the work efficiency of radiologists. Based on the BrainCT-3M and BrainMRI-7M datasets, we pre-trained the text encoder and decoder using a strategy similar to Bidirectional Encoder Representations from Transformers (BERT). Then, we aligned the image encoder and text encoder using the Contrastive Language-Image Pre-Training (CLIP) strategy, thereby training strategy the capability of Brainfound for automatic report generation (Supplementary Fig. 7, Methods). For a comprehensive understanding of a brain CT or MRI scan, Brainfound processes an entire brain CT or MRI scan stack when drafting reports (Fig. 4a, Methods). The alignment of text encoders and image encoders also serves the significant purpose of enabling zero-shot classification of brain CT or MRI scans based on text tokens.

**Fig. 4.**
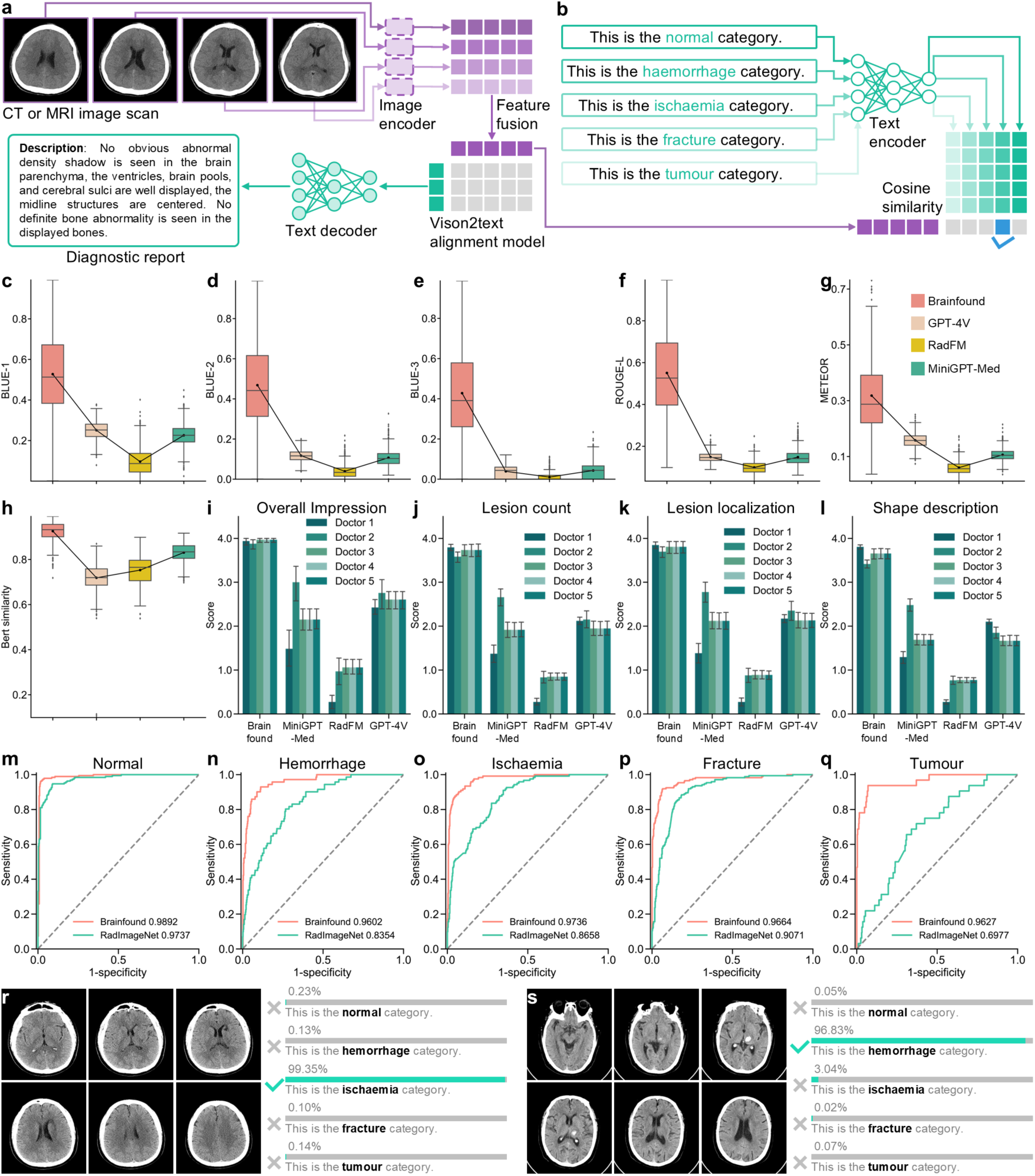
Assessment of Brainfound in automatic report generation. **a**, The image encoder extracts features from CT or MRI image sequences, producing latent space features. Diagnostic reports are processed through the text encoder to acquire latent space features. The features of both the image sequences and their corresponding diagnostic reports are aligned in the latent space of Brainfound. **b**, The alignment model of Brainfound serves directly as a zero-shot classification model. Brainfound can be instructed in natural language to perform brain medical imaging classification. **c-h**, Quantitative comparison of the report generation outcomes for Brainfound, GPT-4V, RadFM, and MiniGPT-Med (n=990). The metrics used for comparison include BLUE-1, BLUE-2, BLUE-3, ROUGE-L, METEOR, and Bert similarity. The higher the scores, the closer the generated reports are to the ground truth reports and the greater the accuracy. **i-l**, Under clinical standards, the reports generated by four models were evaluated by five experienced doctors (n=33). The scoring criteria include overall assessment, number of lesions, location of lesions, and description of lesion shapes. More scoring results can be found in Supplementary Fig. 30. **m-q**, The zero-shot classification results of Brainfound, with RadImageNet as the comparison method. The AUC curves, arranged from left to right, correspond to normal, hemorrhage, ischemia, fracture, and tumor categories. **r**, Classification output probabilities for ischemia type. **s**, Classification output probabilities for hemorrhage type.

We first evaluated the capabilities of Brainfound with GPT-4V, RadFM^64^, and MiniGPT-Med^65^ in terms of automatic report generation. The default prompt was utilized to generate brain CT reports in the final three methods (Supplementary Fig. 25). We collected 990 brain CT scans and their corresponding reports, authored by experienced clinicians, from the Chinese PLA General Hospital to serve as a test set for these four methods. Data for evaluation in this dataset is not included in the pretraining dataset. We employed commonly used natural language processing metrics to quantitatively evaluate the report quality generated by each method, including BLUE-1, BLUE-2, BLUE-3, ROUGE-L, METEOR, and Bert similarity. Brainfound secured the highest scores in all metrics: for BLUE-1(Fig. 4c), Brainfound scored 0.5275 (95% CI 0.5144-0.5405), outperforming the second-place GPT-4V by 110.24%, which scored 0.2509 (95% CI 0.2482-0.2537). In METEOR (Fig. 4f), Brainfound scored 0.3182 (95% CI 0.3098-0.3266), surpassing GPT-4V by 101.46%, which scored 0.1579 (95% CI 0.1563-0.1594). In Bert similarity (Fig. 4h), Brainfound scored 0.9258 (95% CI 0.9233-0.9283), exceeding MiniGPT-Med by 11.42%, which scored 0.8309 (95% CI 0.8284-0.8334). Similar trends are observed in the results of other metrics (Fig. 4d,f,h). A report for a normal brain CT, a report for an ischemic brain CT, and a report for a hemorrhagic brain CT are presented in the supplementary materials(Supplementary Fig. 26-28). In parallel, we randomly selected 33 cases from the results to form a test set and established a human evaluation framework to assess the accuracy of reports generated by four methods (Methods). Based on the 3D slicer, we developed a human scoring framework for report evaluation (Supplementary Fig. 29). Five radiologists from three different hospitals, with an average practice duration of 6.4 years (5, 3, 2, 5, and 17 years respectively), participated in scoring the reports. The reports were evaluated on nine aspects according to clinical guidelines: the overall impression and completeness of the report, the descriptions of lesions for count, localization, morphology, boundary, density, type, and normal structure. For the overall impression (Fig. 4i), Brainfound scored 3.9455 (95% CI 3.9104-3.9805), surpassing the second-place GPT-4V by 51.75%, which scored 2.600 (95% CI 2.4943-2.7057). For the lesion count (Fig. 4j), Brainfound scored 3.4848 (95% CI 3.3382-3.6315), surpassing the second-place GPT-4V by 51.75%, which scored 1.5394 (95% CI 1.4118-1.667). For the lesion localization (Fig. 4k), Brainfound scored 3.4788 (95% CI 3.3322-3.6254), surpassing the second-place MiniGPT-Med by 113.33%, which scored 1.6303 (95% CI 1.4595-1.8011). For the shape description (Fig. 4l), Brainfound scored 3.4788 (95% CI 3.2955-3.6621), surpassing the second-place GPT-4V by 118.91%, which scored 1.5879 (95% CI 1.4156-1.7601). Detailed scoring results are available in Supplementary Fig. 30. We also leveraged the understanding of medical knowledge by current large language models, having GPT-4 and GPT-4o score the reports generated by the four methods like doctors. Reports written by experienced doctors were used as the reference standard for GPT-4 and GPT-4o scoring. GPT-4 and GPT-4o each scored three times, obtaining results close to human scores (Supplementary Fig. 31-32).

Once the image and text encoders are aligned, zero-shot classification tasks can be seamlessly accomplished using the tokens from the text encoder(Fig. 4b). We developed internal and external test sets to evaluate the performance of Brainfound and RadImageNet in zero-shot brain CT classification tasks. The internal test set was gathered at the Chinese PLA General Hospital, containing 588 brain CT scans with corresponding diagnostic results, including 190 normal brain CTs, 71 cases of cerebral hemorrhage, 122 cases of cerebral ischemia, 173 cases of skull fracture, and 32 cases of brain tumor. The external test set was collected in Brains Hospital of Hunan Province and contains 363 brain CT scans with corresponding diagnostic results, including 92 normal brain CTs, 62 cases of cerebral hemorrhage, 160 cases of cerebral ischemia, 24 cases of skull fracture, and 25 cases of brain tumor. Brainfound achieved the highest AUC scores for the classification of the five types of CT scans in both the internal and external test sets (Fig. 4m-q, Supplementary Fig. 33). In internal dataset testing, the AUC of Brainfound for normal brain CT is 0.9892, while RadImageNet is 0.9737, with a difference of 1.59%(Fig. 4m); for brain hemorrhage CT, the AUC of Brainfound is 0.9602, and RadImageNet is 0.8354, with a difference of 14.94%(Fig. 4n); for brain ischemia CT, the AUC of Brainfound is 0.9736, and RadImageNet is 0.8658, with a difference of 12.45%(Fig. 4o); for brain fracture CT, the AUC of Brainfound is 0.9664, and RadImageNet is 0.9071, with a difference of 6.54%(Fig. 4p); for brain tumor CT, the AUC of Brainfound is 0.9627, and RadImageNet is 0.6977, with a difference of 37.98%(Fig. 4q). We visualized the probability distributions of Brainfound for five labels in the brain ischemia and hemorrage CT classification(Fig. 4m-n), where it is evident that the image encoder and text encoder assign higher confidence to the correct label token afterward. Additional visualization results are available in Supplementary Fig. 34. In external dataset tests, the AUC difference between Brainfound and RadImageNet is more pronounced, with the AUC of Brainfound nearly twice that of RadImageNet (Supplementary Fig. 33). Through significance analysis, the attention of Brainfound is concentrated on the regions that determine the image category (Supplementary Fig. 35). The label probability visualizations for the external test set are shown in Supplementary Fig. 33.

### Brainfound handles medical MCQs and free conversations

Finally, as an AI copilot tailored for brain imaging clinical applications, we assessed the capability of Brainfound via more flexible and challenging tasks. By leveraging the image sequences and corresponding diagnostic reports from BrainCT-3M and BrainMRI-7M, in combination with the strong text understanding capabilities of the current large language model (GPT-4), we created an instruction dataset named BrainInstru-1M, which contains 1,003,732 cases (Fig. 4a). BrainInstru-1M includes multiple-choice questions (MCQs) or free conversations: for MCQs, each instruction data contains a brain CT or MRI sequence, the question stem and options, and the correct answer; for free conversations, each instruction data contains a brain CT or MRI sequence along with three rounds of conversations. We employed different prompts to guide GPT-4 in generating data from various knowledge depths and perspectives, enriching the data diversity of BrainInstru-1M.

To evaluate the performance of Brainfound in answering MCQ, we created a test set called BrainMCQ. BrainMCQ consists of 70 brain CT scan samples: 12 normal brain CTs, 14 brain hemorrhage CTs, 20 brain ischemia CTs, 12 brain fracture CTs, and 12 brain tumor CTs (Fig. 5d). Each CT scan contains 3-4 MCQs, with a total of 229 MCQs, which include 3 three-option questions, 215 four-option questions, and 11 five-option questions (Fig. 5e). In terms of the options, we ensure the correct answer is randomly distributed, with no bias towards any specific option (Fig. 5f). We recruited three experienced doctors from three different hospitals, with an average of 3.3 years of professional experience (2, 3, and 5 years, respectively), to compare their accuracy in answering BrainMCQ questions with that of Brainfound and GPT-4V. To evaluate the performance, we asked both Brainfound and GPT-4V to independently answer BrainMCQ three times and calculated the average accuracy. The average accuracy of Brainfound is 0.7846, the average accuracy of GPT-4V is 0.5313, the accuracy of Doctor 1 is 0.5749, the accuracy of Doctor 2 is 0.5072, and the accuracy of Doctor 3 is 0.7393. Brainfound achieved the same level of accuracy as human doctors (Fig. 5a). In the time statistics for completing BrainMCQ, both Brainfound and GPT-4V finished the test within half an hour, while the three doctors took around one hour or longer (Fig. 5b). Some MCQ cases are shown in detail (Fig. 5h-i, Supplementary Fig. 36-37).

**Fig. 5.**
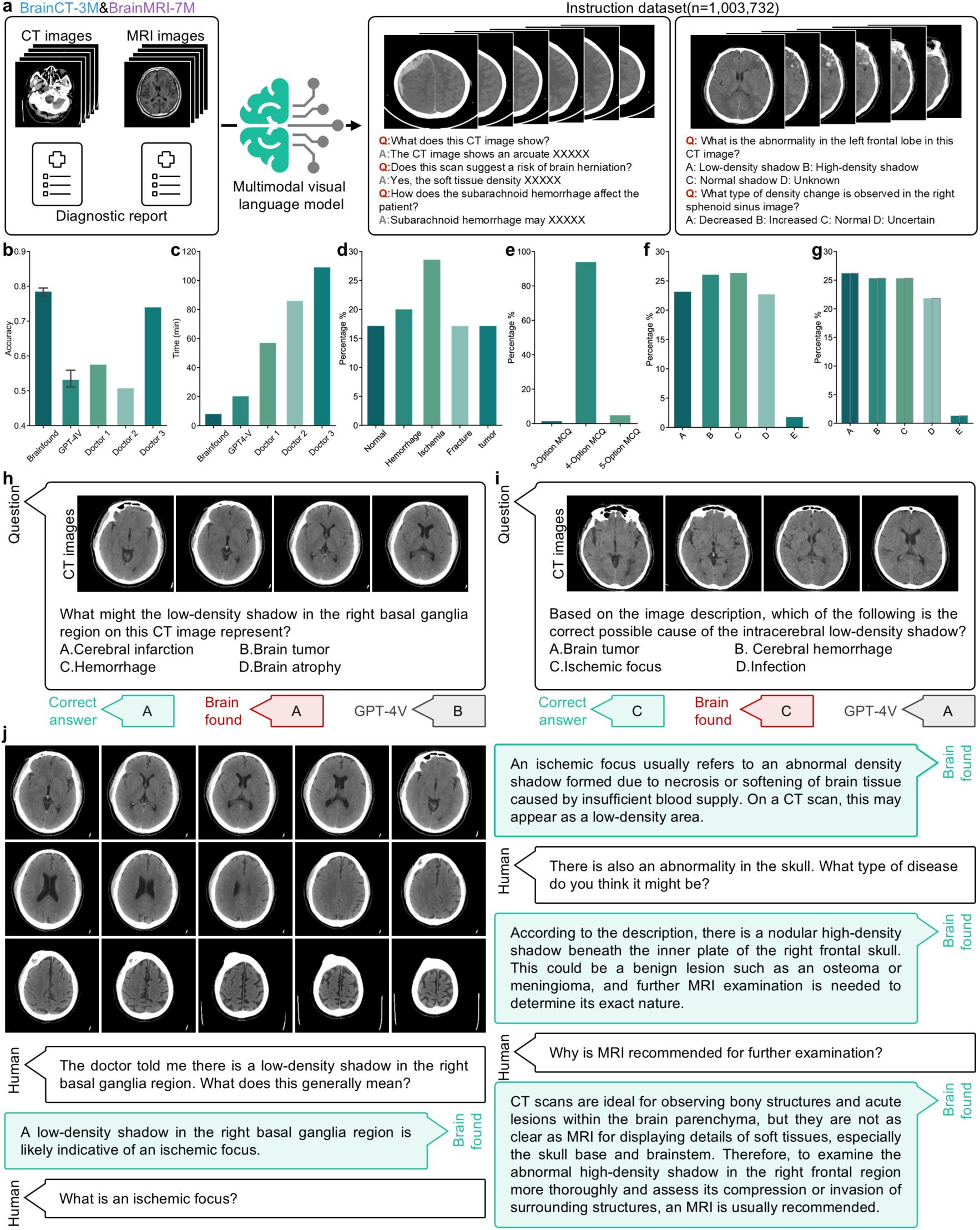
Evaluation of Brainfound on multiple-choice questions and free conversations. **a**, Leveraging the advanced capabilities of GPT-4, a multimodal brain imaging dataset comprising 1,003,732 instructions and corresponding responses has been constructed. Every scan sequence and its paired report from BrainCT-3M and BrainMRI-7M are utilized to generate MCQs and multi-turn conversations on various aspects. **b**, The response accuracy of Brainfound, GPT-4V, and two proficient doctors on BrainMCQ. Both Brainfound and GPT4-V underwent evaluation three times. Error bars represent the 95% confidence interval. **c**, The time taken by Brainfound, GPT-4V, and two skilled doctors to complete BrainMCQ. The average time for three evaluations by Brainfound and GPT-4V is displayed. **d**, The percentage of questions related to normal, cerebral hemorrhage, cerebral ischemia, brain tumor, and fracture types in BrainMCQ. **e**, The proportion of three-option, four-option, and five-option multiple-choice questions in BrainMCQ. **f**, The proportion of each option in the correct answers of BrainMCQ. **g**, The proportion of each option in the answers of Brainfound and GPT-4V. **h-i**, Two cases of Brainfound and GPT-4V answering MCQs. Brain CT sequences and questions are fed into the models, which subsequently provide the chosen answers. **j**, A case of Brainfound in the free conversation. Brain CT image sequences serve as input for Brainfound, allowing humans to engage in multiple rounds of conversation based on the image information.

Besides testing on BrainMCQ, we explored the potential of using Brainfound as a specialized brain imaging AI copilot in free conversations. Brainfound is capable of accurately diagnosing disease conditions by integrating brain CT images, answering complex medical concepts, and even offering further examination suggestions for diseases currently not fully determined (Fig. 5j). During the conversation about cerebral infarction CT scans, Brainfound explained the causes of cerebral infarction and noted a hemorrhage in another area (Supplementary video 1). In the conversation about cerebral hemorrhage CT scans, Brainfound analyzed the effects of the hemorrhage on the lateral ventricles and the subarachnoid space, aiding patients in understanding their condition more comprehensively (Supplementary video 1).

## Discussion

In this study, we established a comprehensive specialized multimodal model for brain medical imaging called Brainfound (Supplementary Fig. 38). We collected two foundational datasets aligned with scans and reports, BrainCT-3M and BrainMRI-7M, covering common brain medical imaging modalities and disease types. Based on this, we generated the instruction dataset related to brain medical imaging, BrainInstru-1M, which includes 1,003,732 cases of brain medical imaging and corresponding instruction texts. Using the DDPM strategy, we pre-trained image encoders and decoders with text embedding enhancement via the cross-attention module as the visual module of Brainfound. Based on the RSNA dataset, we verified that Brainfound achieved the highest AUC in the intracranial hemorrhage classification task across different training data volumes. In the tasks of hemorrhage segmentation and midline shift segmentation, the pre-trained Brainfound also achieved the highest Dice coefficient across various training data volumes. For the task of automatic report generation, we created a robust human-machine evaluation system and recruited five experienced doctors from different hospitals to assess the report generation results. In comparison with current common medical report generation methods, Brainfound outperformed the second-best method by 51.75% in evaluation scores. Additionally, we found that when the text encoder and image encoder are aligned, zero-shot brain medical imaging classification is possible using the input text. Brainfound achieved superior AUCs on both internal and external test datasets compared to RadImageNet. We evaluated the multimodal understanding capabilities of Brainfound through tasks requiring flexibility and a deep understanding of medical images and knowledge, like multiple-choice questions and free conversation. In the multiple-choice question task, the accuracy of Brainfound exceeded GPT-4V by 47.68%, rivaling experienced human doctors. In the free conversation around brain medical imaging, Brainfound is able to correctly answer medical knowledge, diagnose diseases in images, and proactively provide subsequent diagnostic opinions or treatment plans.

Clinical medical AI models are inherently multimodal big data regression tasks. In the future, we plan to integrate additional modalities into Brainfound (such as EEG and electronic medical records), positioning patient records as a central component of prompts to enhance the precision and complexity of disease diagnosis. By incorporating medication information, we aim to train Brainfound to automatically offer medication recommendations. We expect that with further training using extensive multimodal datasets (including medical guidelines and research papers), Brainfound will develop emergent insights related to brain medical imaging. In the high-dimensional space, brain imaging information is often fragmented and isolated; Brainfound will act as an exceptional interpolator to bridge these gaps. Brain diseases often progress over time for patients. Therefore, Brainfound needs to prioritize developing its temporal causal reasoning abilities, allowing it to deduce the disease course by integrating transformations in medical images at several time points, offering more precise prognoses and treatment strategies. On the other hand, medical AI models are responsible for doing everything possible to enhance human health in global clinical applications. By employing techniques like model distillation, quantization, pruning, and leveraging powerful cloud computing platforms, we will deploy Brainfound in the cloud. This will allow Brainfound to function as an AI copilot for clinicians working with brain medical imaging.

## Methods

### Pretrain strategy of Brainfound

To establish BrainFound as an outstanding multimodal AI assistant for brain imaging analysis, our model is designed with three main components: an image encoder, an image decoder, and a large language model. We further propose a three-stage training strategy to enhance its performance. In the first stage, we adopt a diffusion model-based training approach as the pretraining strategy for the image encoder and decoder. This stage enables the model to effectively capture low-level features from medical images, which is essential for tasks such as segmentation, denoising, and modality conversion. In the second stage, we implement contrastive learning based on the pre-trained image encoder and the BERT model. This stage equips the model with the capabilities of report generation and zero-shot classification. In the third stage, we fine-tune the image encoder from the second stage and the large language model InternLM^66^ using multimodal dialogue datasets and multiple-choice question datasets. This enables the model to serve as an AI assistant capable of answering questions effectively.

**Self-supervised training for feature representation:** To accommodate a broader range of low-level downstream tasks, we adopted the training methodology of diffusion models as our first-stage pretraining strategy. Diffusion models are widely recognized for their ability to generate realistic images from Gaussian noise. Recent research has shown that these models can effectively capture stable prior knowledge, leading to improved performance across a variety of downstream tasks. Therefore, we leverage diffusion models as a self-supervised pretraining approach. Before training, the images were preprocessed by converting them into different window widths and window levels. The training process of diffusion models consists of two key phases: the forward diffusion process and the reverse diffusion process. During the forward diffusion phase, noise is gradually added to the data. The objective of DDPM is to train a model capable of reconstructing the original data from these noisy observations. For our training, we adhered to the standard settings^67^. To enhance control over the model’s generated content and expand its range of applications, we adopt a cross-attention-based DDPM model. The window width, window level, and modality information of the image are used as conditional inputs to guide the learning process during generation. To improve robustness, the conditional information may be randomly dropped out during training.

**Contrastive Learning:** Contrastive Learning is a self-supervised learning technique that trains models on unlabeled data by learning meaningful representations through the similarities between data samples. This approach is particularly effective in scenarios with limited labeled data. By utilizing contrastive learning, the image encoder extracts features that align with semantically meaningful text in the feature space. This alignment facilitates applications such as image-text retrieval and medical image captioning. In this stage, we use the encoder from the pre-trained model in the first stage to extract image features from an image sequence. These features are concatenated and then passed through an aggregation module and a projection module to generate a feature vector. This feature vector is compared with the features extracted by a text encoder to calculate similarity, and the loss is computed accordingly. For the text encoder, we employ a BERT structure fine-tuned on Chinese-language corpora. Using a rule-based report analysis method, we extract CT image categories, including normal, hemorrhage, cerebral infarction, fracture, and tumor. Since MRI scans encompass multiple distinct modality sequences, the report content is characterized by its comprehensive and summarized nature, and we leverage ChatGPT to extract key disease-related terms from the reports to ensure an accurate and comprehensive representation of disease information. When constructing the text for contrastive learning, we concatenate the extracted disease categories with the original reports.

**Multimodal Fine-Tuning Phase:** To enable the multimodal assistant to fully understand both images and text, we fine-tune the model using the image encoder from the contrastive learning stage and a large language model (LLM) based on the open-source LLaMA architecture. When constructing the multimodal training dataset, we utilized ChatGPT to clean and organize the report data. By designing prompts, we transformed the reports into conversational text of various styles and created multiple-choice questions. Detailed prompts are provided in the supplementary materials. Using this approach, we generated a total of *N* rounds of dialogue text and *N* sets of multiple-choice question text. Given the significant number of parameters in large models, fully fine-tuning all parameters for downstream tasks requires substantial computational resources and is prone to overfitting. Moreover, full fine-tuning can lead to severe forgetting issues, causing the model to lose many of its original capabilities. To address these challenges, we adopted the PEFT (Parameter-Efficient Fine-Tuning) method based on LoRA (Low-Rank Adaptation) to fine-tune both the LLaMA language model and the image encoder model.

### Network architecture

Our BrainFound framework consists of an image encoder, an image decoder, and a foundational large language model. BrainFound leverages diffusion models for pretraining to obtain robust and meaningful feature representations. During the self-supervised diffusion phase, the image encoder and image decoder are connected in a UNet-like architecture. To better capture feature representations across multiple levels, we chose a pixel-to-pixel space diffusion model instead of the Latent Diffusion Model (LDM)^68^. Specifically, the image encoder in BrainFound consists of five downsampling modules and one deep feature extraction module, while the image decoder comprises five upsampling modules. Each of these modules incorporates residual structures to ensure effective gradient optimization during training. Moreover, certain downsampling and upsampling blocks are enhanced with a cross-attention Transformer module. This mechanism enables the encoded textual information to directly influence the image generation process. Such textual information includes, but is not limited to, parameters like the image’s window width and window level, as well as disease category information extracted from reports.

In the contrastive learning phase, we additionally designed an aggregation module to fuse the features of multiple images within an image sequence. The aggregation module adopts a transformer architecture consisting of two layers of transformer encoders with layer normalization. Positional encoding parameters are also included to enhance the model’s ability to process sequential information.

During the fine-tuning phase of the multimodal assistant, we selected LLaMA as the foundation for the large language model. LLaMA is a highly optimized large-scale language model based on the Transformer architecture, designed for performance and efficiency. Its core structure incorporates multi-head self-attention (MHSA) to capture long-range dependencies efficiently, enabling parallel processing of contextual relationships and maintaining semantic and syntactic consistency during generation. To enhance contextual understanding, LLaMA employs rotary position embedding (RoPE), which provides strong generalization capabilities for modeling long sequences. In this work, we utilized the 7B-parameter version of the model as a balance between performance requirements and computational resources. The pre-trained weights were sourced from InternLM. Subsequently, we adopted an instruction learning approach to fine-tune the image encoder and the LLaMA model using multimodal dialogue data and multimodal multiple-choice question datasets. Specifically, for a given dialogue, we concatenate the image-encoded features with the text-extracted tokens and input them into the LLaMA model to generate outputs. A detailed process flowchart can be found in the supplementary materials. (Supplementary Fig. 1).

### Fine-tuning Brainfound to downstream tasks

To fully unlock the potential of Brainfound across diverse tasks, we incorporated multiple state-of-the-art deep learning techniques and designed experiments tailored to various downstream applications.

**RSNA intracranial hemorrhage classification task:** Intracranial hemorrhage classification is essential for identifying the underlying cause of bleeding, guiding treatment decisions, and optimizing management strategies. It provides a foundation for prognosis evaluation, personalized treatment planning, and advancing medical research. In this task, we utilize an image encoder to extract image features and perform classification through an additional linear layer. Specifically, the image at t = 0 is input into the image encoder to extract features, which are then passed through a dropout layer and an activation function before being fed into the linear layer for prediction. As this is a multi-label classification problem, binary cross-entropy (BCE) is employed to calculate the loss. We evaluated three experimental setups on this dataset: Full-parameter fine-tuning: Both the image encoder and the linear layer parameters are updated during training. Linear-layer fine-tuning: The image encoder is frozen, and only the linear layer weights are fine-tuned. Ensemble integration: Our image encoder was incorporated into the winning ensemble strategy of the RSNA competition for further evaluation.

**Intracerebral hemorrhage and midline structure segmentation task:** Brainfound contains rich prior knowledge of brain medical images. To fully exploit this prior knowledge for segmentation, which is a dense prediction task, we adopted the approach^69^ that utilized MLP to classify each pixel’s label. In summary, we used the image encoder and image decoder of Brainfound to extract image features and trained an MLP classifier to classify the features extracted from each spatial location. For each image, we obtained four features at different scales from Brainfound, which were then upsampled to match the input resolution and concatenated. The feature vector corresponding to each spatial location was then fed into the MLP classifier to predict the class of that pixel, and the loss was computed with the segmentation labels to update the network. During training, we used single-center data and split it into training, validation, and testing sets. The best model was selected based on the validation set, and results were reported on the test set. The AdamW optimizer was used with a weight decay of 1e-3 and an initial learning rate of 1e-3. The training lasted for 20 epochs.

**MRI imaging modality translation task:** For the task of MRI modality conversion, we conducted four experiments, converting T1WI into T2WI, FLAIR, and DWI, with the latter further divided into two classes: b<500 and b>500. For this task, we employed a straightforward conditional diffusion model to perform the modality conversion. Specifically, the input to the diffusion model consisted of both the noise channel and an additional T1WI as the condition. We acknowledge that more advanced diffusion-based mechanisms might achieve better results in this task. For our experiments, we curated a dataset of 200 cases containing these modalities. Among them, 100 cases were used for training, and the remaining 100 for validation. The model was trained for 200 epochs, and the final model was evaluated on the validation set.

**Low-quality medical image enhancement task:** The visual module of Brainfound employs the DDPM strategy for pretraining, which provides strong representation learning capabilities for pixel-level semantic information. This capability is leveraged to develop a zero-shot denoising framework. For the detailed algorithmic process, see Supplementary Fig. 13.

**Zeroshot classification task:** To validate the effectiveness of contrastive learning, we collected two classification datasets. The first dataset, consisting of 588 cases, was sourced from an internal center and is entirely separate from the training data used for contrastive learning. The second dataset, obtained from an external center, contains 363 cases. Both datasets include five categories: normal, hemorrhage, ischemia, fracture, and tumor. For zero-shot classification, we constructed textual features for the five categories using short descriptive phrases. The cosine similarity between the textual features and the image features was then computed. After normalizing the similarity scores, softmax probabilities were calculated to predict the final category.

**Medical image report generation task:** During the training of contrastive learning, we additionally designed a text decoder module. This module is based on a pre-trained Chinese BERT architecture with six hidden layers and a vocabulary size of 21,128. The module takes image features as input and predicts the probability distribution of the corresponding text. The predicted results are further refined using a beam search algorithm to generate the final version of the medical image report. In this task, we compared our approach with several baseline models (RadFM, MiniGPT-Med, and GPT-4V), all of which can generate reports based on images. For RadFM and MiniGPT-Med, we utilized the prompts provided in the authors’ examples to generate report outputs from medical images. For GPT-4V, we designed custom prompts as input. Detailed prompts can be found in the supplementary materials. For evaluation, we adopted standard quantitative metrics such as BLEU, ROUGE-L, and METEOR. Additionally, we invited five physicians to rank the reports generated by the four methods. The evaluation criteria were designed based on physicians’ suggestions and included 10 subcategories. The rank method is similar to GPA calculation, where the best items are assigned 4 points, the worst are given 1 point, and certain unacceptable cases are assigned 0 points.

**AI assistant assessment task:** To validate the functionality of the multimodal assistant, we designed two experiments: multiple-choice question evaluation and free-form question-answering. For the multiple-choice questions, we compared our model, BrainFound, with GPT-4o. Two physicians were invited to complete the questions as well. Both BrainFound and GPT-4o were generally able to provide consistent and well-formatted answers. However, GPT-4o occasionally failed to answer certain questions. For these cases, we repeatedly queried the API until stable responses were obtained. We compared the performance of BrainFound, GPT-4o, and the two physicians in terms of answer accuracy and response time. The detailed results are presented in Figure 5.

### Evaluation metrics for pixel-level tasks

Several metrics are commonly used to evaluate the performance of image enhancement task. Among them, the Peak Signal-to-Noise Ratio (PSNR) is widely recognized as a standard for assessing image quality. A higher PSNR value indicates better image fidelity. If the ground truth image is *y*, and the raw image is *x*, then the definition of PSNR is as follows:

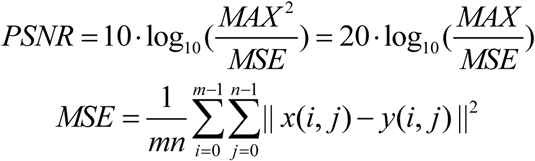

Here *MAX* is the maximum pixel value. For normalized images *MAX* = 1. *m* and *n* are the two dimensions of the image.

We also compute the Signal-to-Noise Ratio (SNR) to evaluate the performance of various methods. Let *I*_(*i*,*j*)_ represent the raw image pixel values and *K*_(*i*,*j*)_ denote the model output, and the formula is as follows:

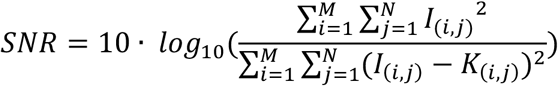

Here, *M* and *N* represent the width and height of the image, respectively.

Root Mean Square Error (RMSE) directly quantifies the variance between two images. An RMSE value approaching 0 indicates better preservation of visual information between the reconstructed image and the ground truth. RMSE is defined as follows:

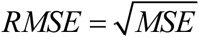

Structural Similarity Index (SSIM) is a widely used metric for quantifying the similarity between two images. SSIM evaluates similarity by independently comparing three key components: luminance, contrast, and structural information. These components are then weighted and combined into a single score to represent the overall similarity. The calculation of SSIM is performed using a sliding window applied across the image. In this process, a window with dimensions *a* × *a* is selected from the image for each calculation, and the SSIM is computed for that specific window. The overall SSIM for the image is then obtained by averaging the SSIM values from all such windows after the entire image has been scanned. A higher SSIM score indicates superior image quality. SSIM is defined as follows:

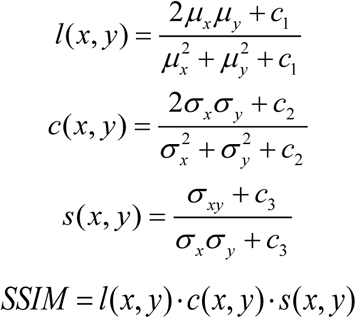

Here, *μ_x_* and *μ _y_* represent the mean values of *x* and *y*; *α _x_* and *α _y_* represent the variances of *x* and *y*, *α _xy_* represents the covariance between *x* and *y*. *c*_1_, *c*_2_, *c*_3_ are three constants.

### Evaluation metrics for report generation

BLEU is a widely used metric for evaluating the quality of machine-generated text, particularly in tasks such as machine translation and text summarization. The BLEU score is calculated based on n-gram precision, which measures the overlap between n-grams in the generated text and those in the reference text. BLEU-1 to BLEU-4 represent the scores computed using unigrams, bigrams, trigrams, and 4-grams, respectively, capturing different levels of linguistic context. The following is the formula:

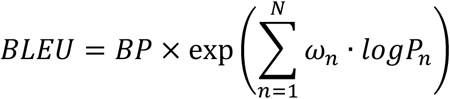

*BP* is the brevity penalty addresses the issue of overly short translations. *P*_*n*_ is The precision for *n* grams. *n* ranges from 1 to 4 for BLEU-1 to BLEU-4. *ω*_*n*_ is the weight assigned to each *n* gram precision.

ROUGE-L (Recall-Oriented Understudy for Gisting Evaluation - Longest Common Subsequence) is a widely used metric for evaluating the quality of machine-generated text, particularly in summarization tasks. Unlike n-gram-based metrics, ROUGE-L measures the overlap between the candidate text and reference text based on their longest common subsequence (LCS). This approach takes into account both the order and presence of words, making it well-suited for capturing fluency and relevance in text generation. The following is the formula:

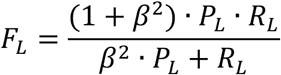

where 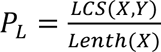 is precision and 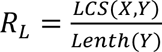 *is recall. LCS*(*X*, *Y*) represents the length of the longest common subsequence between the candidate text *X* and the reference text *Y*. And *β* is a weighting parameter (usually set to 1) to balance the importance of precision and recall.

METEOR (Metric for Evaluation of Translation with Explicit ORdering) is a popular evaluation metric for machine-generated text, particularly in machine translation and text generation tasks. Unlike n-gram-based metrics such as BLEU, METEOR focuses on aligning words in the candidate and reference texts using advanced matching techniques, making it more robust and sensitive to variations in word order and synonymy. In METEOR, the precision and recall are combined into an F-score, with recall typically weighted more heavily:

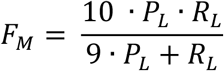

Then, a penalty is applied to account for disjoint word matches, penalizing cases where matched words are far apart or unordered. The penalty is calculated as follows:

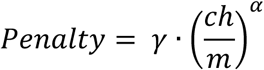

where *ch* is the number of chunks (continuous sequences of matched words), *m* is the total number of matches, and *γ* and *α* are hyperparameters. The final formula is:

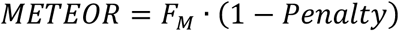

### Visualization of saliency maps

The Grad-CAM^72^ technique is harnessed to craft the Saliency Map for the input image model. Initially, the activation feature maps of the convolutional layers are derived via forward propagation, and subsequently, the gradients of these feature maps concerning the target class are computed through backpropagation. Following this, global average pooling is applied to these gradients to acquire the channel weights. These weights are then utilized to modulate the activation feature maps of the convolutional layers, culminating in a two-dimensional heat map of weighted summation, elucidating the significance of distinct regions within the input image for the target category. Following this, the heat map undergoes upscaling to match the input image’s dimensions using bilinear interpolation. Lastly, the heat map is rendered visually through color mapping to exhibit the areas of interest identified by the model. The contour map delineates lines of uniform value within a saliency map.

## Data availability

All relevant data that support the findings of this study are available from the corresponding authors upon reasonable request.

## Code availability

Our Brainfound can be found at https://github.com/gingerbread000/Brainfound.

## Data Availability

All data produced in the present study are available upon reasonable request to the authors

## Acknowledgments

This work was supported by NSFC (No. 62088102, 62222508, 62071272, 82327803, 82441014, and 82202133) and MOST(No.2020AA0105500).

## Author contributions

Q.D., X.L., and Y.G. conceived the Brainfound project and revised the manuscript. G.Z. and Z.G. implemented the Brainfound framework, trained the multimodality model, completed the fine-tuning of downstream tasks, organized the experimental results, and composed the manuscript. C. D. collected data and established the BrainCT-3M and BrainMRI-7M datasets. J.L. completed the saliency visualization of Brainfound attention. T.W., Y.G., and Y.C. established the BrainInstru-1M instruction dataset. L. W. collected 5T MRI brain data. Y. L., Y. L., Q. C., K. F., and L. W. completed the human-machine evaluation of automatic report generation. Q. C., K. F., and Y. L. Q. C., L. W., and Y. L. completed the answering of BrainMCQ.

