## Supplementary Materials for "A Multimodal Vision-text AI Copilot for Brain Disease Diagnosis and Medical Imaging"

### 3 Supplementary Figures

| | |
|---|---|
| **Supplementary Figure 1** | The fundamental details of BrainCT-3M used for pre-training Brainfound. |
| **Supplementary Figure 2** | The fundamental details of BrainMRI-7M used for pre-training Brainfound. |
| **Supplementary Figure 3** | The network structure of the visual module in Brainfound. |
| **Supplementary Figure 4** | The pre-training and fine-tuning process of the image encoder-decoder in Brainfound. |
| **Supplementary Figure 5** | The generated brain images from the vision module of Brainfound during the pre-training process. |
| **Supplementary Figure 6** | The visualization of features extracted by Brainfound and clustered by k-means. |
| **Supplementary Figure 7** | The alignment process between the text encoder and the image encoder of Brainfound. |
| **Supplementary Figure 8** | The training process for human-AI conversations. |
| **Supplementary Figure 9** | Performance of Brainfound on the brain hemorrhage classification task at RSNA. |
| **Supplementary Figure 10** | Saliency maps produced by Brainfound for cerebral hemorrhage classification in RSNA. |
| **Supplementary Figure 11** | Saliency maps generated by Brainfound for cerebral hemorrhage segmentation in RSNA. |

| | |
|---|---|
| **Supplementary Figure 12** | Saliency maps generated by Brainfound for brain midline segmentation. |
| **Supplementary Figure 13** | The zero-shot denoising mechanism of Brainfound. |
| **Supplementary Figure 14** | Comparison of the denoising results of Brainfound on 3T MRI images with simulated noise. |
| **Supplementary Figure 15** | The enhancement of Braindfound on T2WI captured by the 5T MRI. |
| **Supplementary Figure 16** | The enhancement of Braindfound on T1WI captured by the 5T MRI. |
| **Supplementary Figure 17** | The enhancement of Braindfound on real-world images captured by the 5T MRI in the external center. |
| **Supplementary Figure 18** | The comparison of zero-shot enhancement results on 0.3T FLAIR. |
| **Supplementary Figure 19** | The comparison of zero-shot enhancement results on 0.3T T1WI. |
| **Supplementary Figure 20** | The comparison of zero-shot enhancement results on 0.3T T2WI. |
| **Supplementary Figure 21** | The performance of Brainfound in translation MRI T1WI to T2WI. |
| **Supplementary Figure 22** | The performance of Brainfound in translation MRI T1WI to FLAIR. |

| | |
|---|---|
| **Supplementary Figure 23** | The performance of Brainfound in translation MRI T1WI to standard -b-value DWI. |
| **Supplementary Figure 24** | The performance of Brainfound in translation MRI T1WI to low-b-value DWI. |
| **Supplementary Figure 25** | The prompts utilized during the report generation process. |
| **Supplementary Figure 26** | The report generation result of a normal brain CT scan. |
| **Supplementary Figure 27** | The report generation result of an ischemic brain CT scan. |
| **Supplementary Figure 28** | The report generation result of a hemorrhage brain CT scan. |
| **Supplementary Figure 29** | The report evaluation system and MCQ answering system based on 3D Slicer. |
| **Supplementary Figure 30** | The results of scoring by experienced doctors on reports generated from four methods. |
| **Supplementary Figure 31** | The results of scoring on reports generated by four methods using GPT-4. |
| **Supplementary Figure 32** | The results of scoring on reports generated by four methods using GPT-4o. |
| **Supplementary Figure 33** | The zero-shot classification results of Brainfound on the external test set. |

| | |
|---|---|
| **Supplementary Figure 34** | The zero-shot classification results of aligned image encoder and text encoder in Brainfound. |
| **Supplementary Figure 35** | Saliency maps generated by Brainfound for zero-shot classification. |
| **Supplementary Figure 36** | The responses of Brainfound on several multiple-choice questions about brain imaging, Part I. |
| **Supplementary Figure 37** | The responses of Brainfound on several multiple-choice questions about brain imaging, Part II. |
| **Supplementary Figure 38** | Schematic diagram summarizing the evaluation results of Brainfound. |

**Video captions**

| | |
|---|---|
| **Supplementary Video 1** | The demo of Brainfound on free conversation around brain CT images. Two cases are presented: case 1 discusses what a cerebral infarction is and what a high-density linear signal shadow is. Case 2 is about cerebral hemorrhage, discussing what cerebral hemorrhage is and its effects on other areas. |

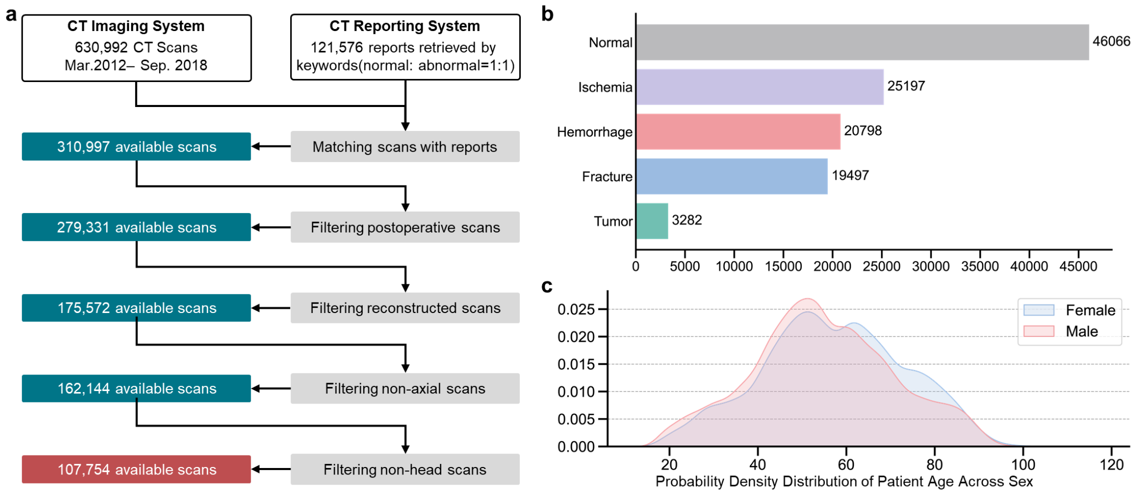


**Supplementary Figure 1**

**The fundamental details of BrainCT-3M used for pre-training Brainfound**. **a,** Data preprocessing procedure. A total of 630,992 brain CT scans and 121,576 diagnostic reports were collected as the foundational database. These were subsequently screened and refined by the image quality and a natural language processing (NLP) model combined with reports (Methods), culminating in the pre-training dataset BrainCT-3M. BrainCT-3M comprises 107,754 brain CT scans and corresponding reports, totaling over 3 million images. **b,** The number of CT scans for each type contained in BrainCT-3M. **c,** The age distribution of patients in BrainCT-3M.

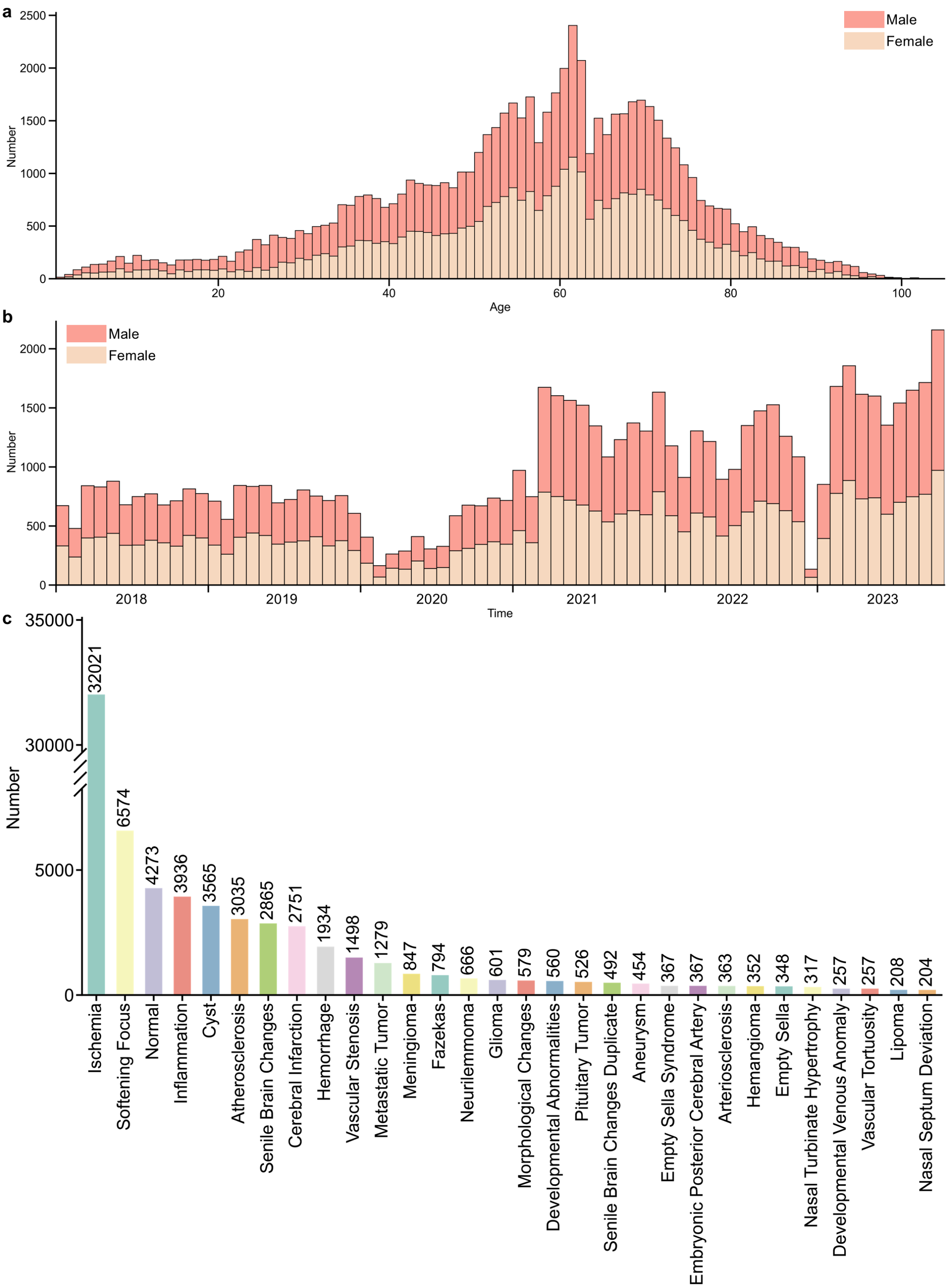


**Supplementary Figure 2**

**The fundamental details of BrainMRI-7M used for pre-training Brainfound**. **a**, Age distribution of patients collected in BrainMRI-7M. BrainMRI-7M consists of brain MRI scans from 68,653 patients, totaling approximately 7 million images. **b**, Distribution of admission times in BrainMRI-7M. The dataset includes MRI images over five years, from 2018 to 2023. **c**, Token statistics in corresponding BrainMRI-7M reports. We utilized GPT-4, a robust automated tool for analyzing MRI reports, to perform tokenization on BrainMRI-7M. After analyzing 68,653 MRI reports, the top 30 most frequent clinical medical terms were identified and presented.

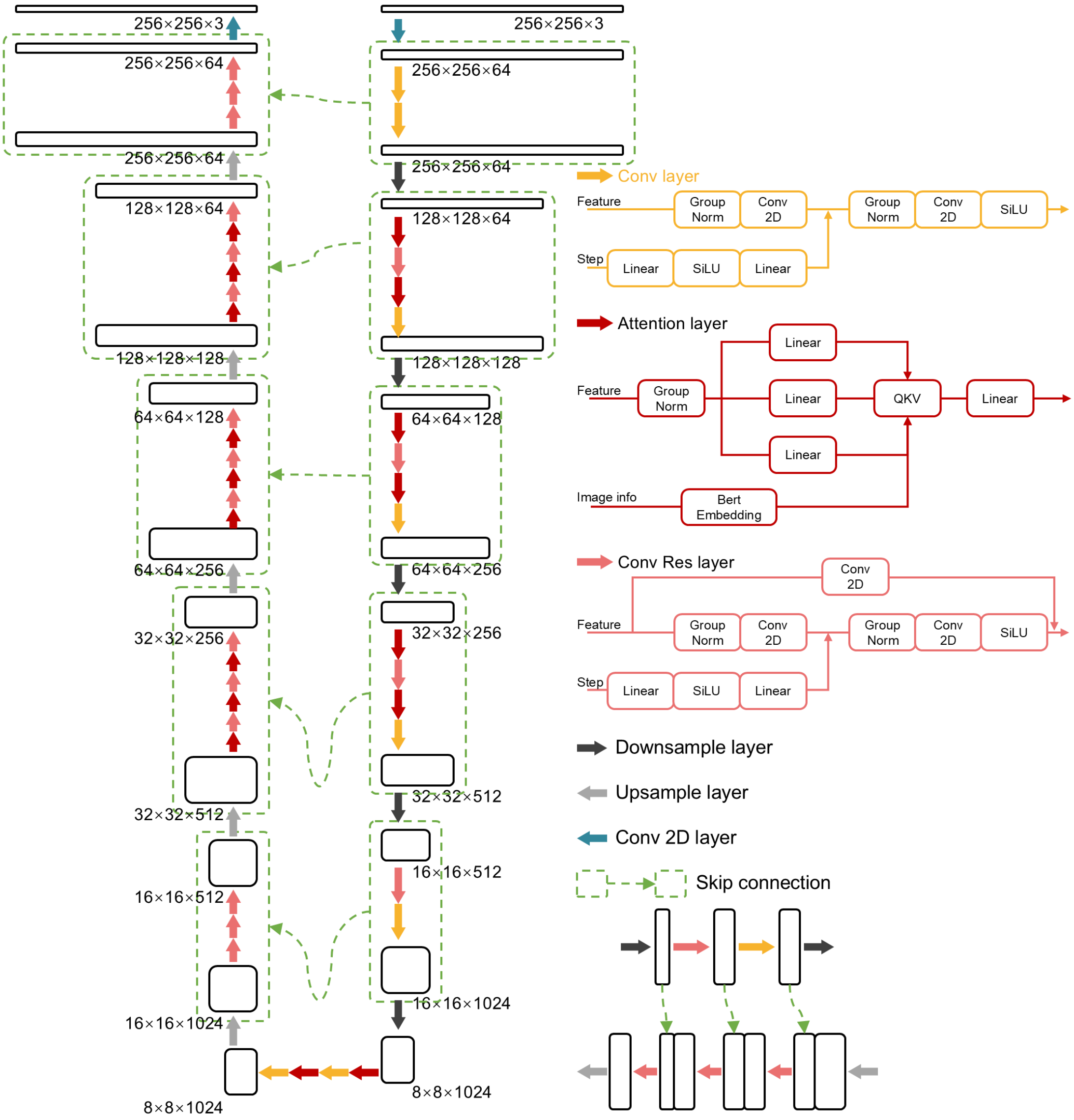


**Supplementary Figure 3**

**The network structure of the visual module in Brainfound**. The visual module of Brainfound features a U-shaped network. Input images are pooled progressively layer by layer to capture more global features. Basic image information is embedded into the neural network using the BERT module, and the final image reconstruction is controlled via cross-attention. On the left is the network architecture diagram, and on the right is a detailed explanation of each icon.

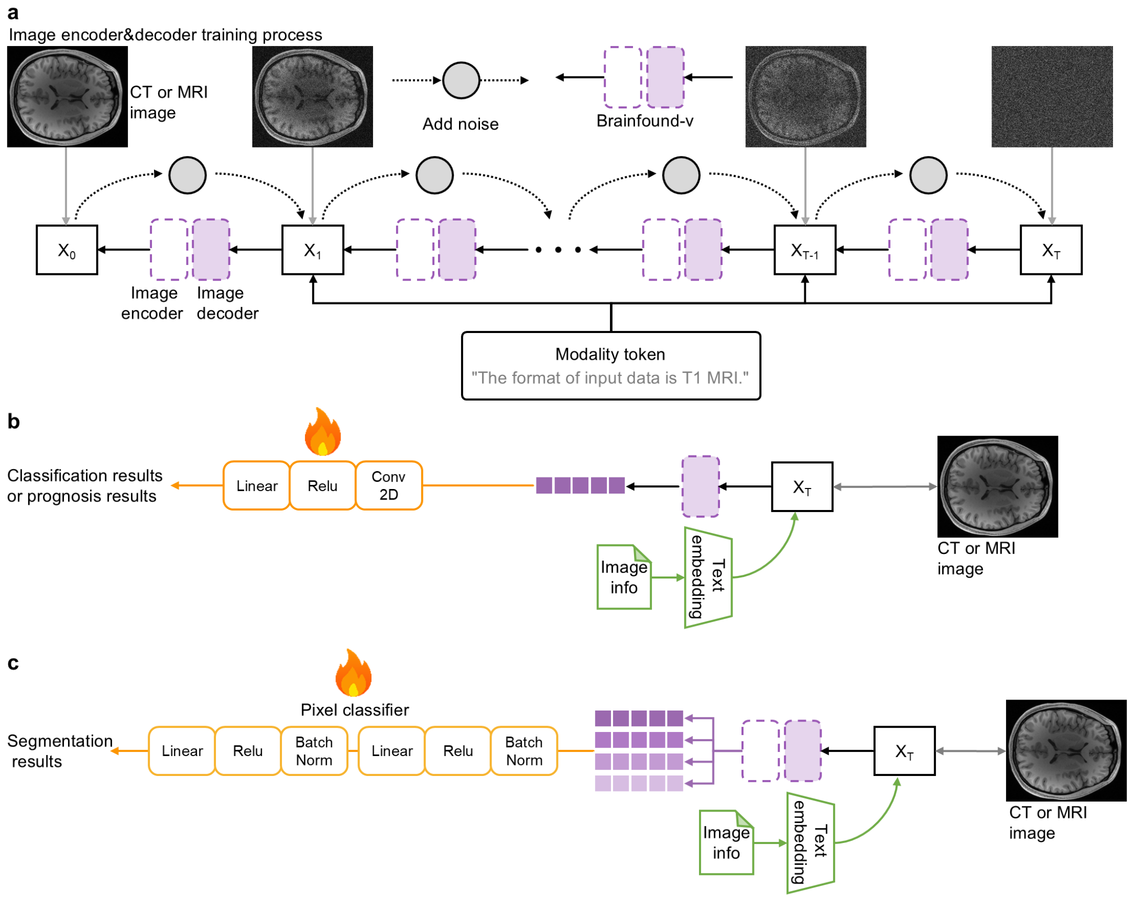


**Supplementary Figure 4**

**The pre-training and fine-tuning process of the image encoder-decoder in Brainfound**. **a**, The image encoder and decoder of Brainfound are pre-trained following the DDPM strategy. Clear brain CT or MRI images are gradually degraded by noise until a completely noisy image is obtained. The image encoder and decoder systematically learn the denoising process, reconstructing a pristine brain CT or MRI image from pure noise. The fundamental characteristics of the image, such as modality, is randomly selected as a token input into the visual network to facilitate the final reconstruction of the image. **b**, The features obtained by the Brainfound image encoder are input into the MLP, which outputs classification labels. When using the image encoder of Brainfound for diagnosing or classifying brain diseases, only a small amount of labeled training data is needed to fine-tune the MLP module at the end of the whole network. **c**, The features extracted by the Brainfound image encoder and decoder are concatenated and input into the pixel classifier to obtain the lesion localization or segmentation mask. The pixel classifier needs a small quantity of labeled data for fine-tuning.

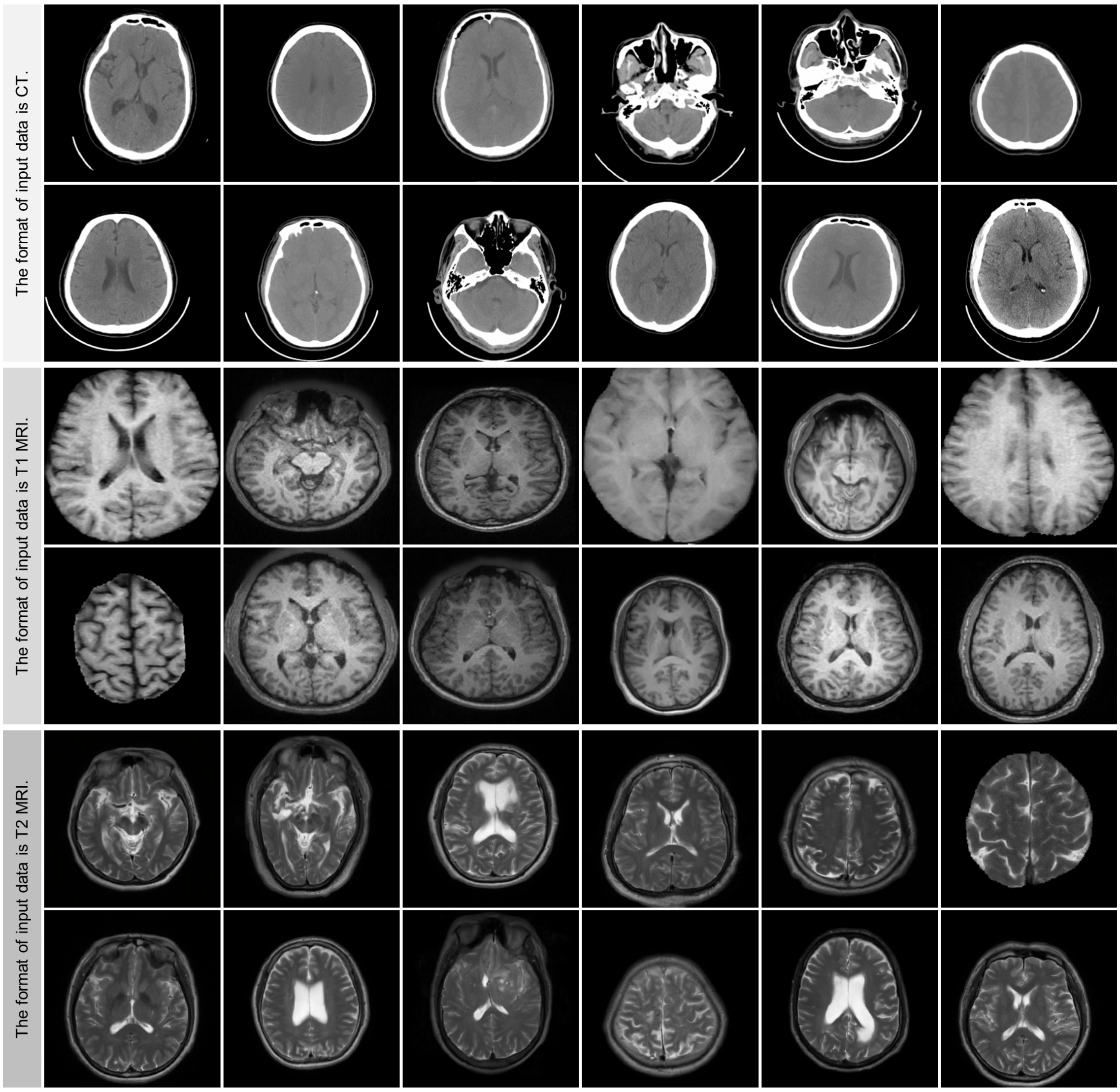


**Supplementary Figure 5**

**The generated brain images from the vision module of Brainfound during the pre-training process**. Images are generated guided by modal information. Brain CT images, brain MRI T1WI, and brain MRI T2WI are displayed separately. Each modality showcases 12 images.

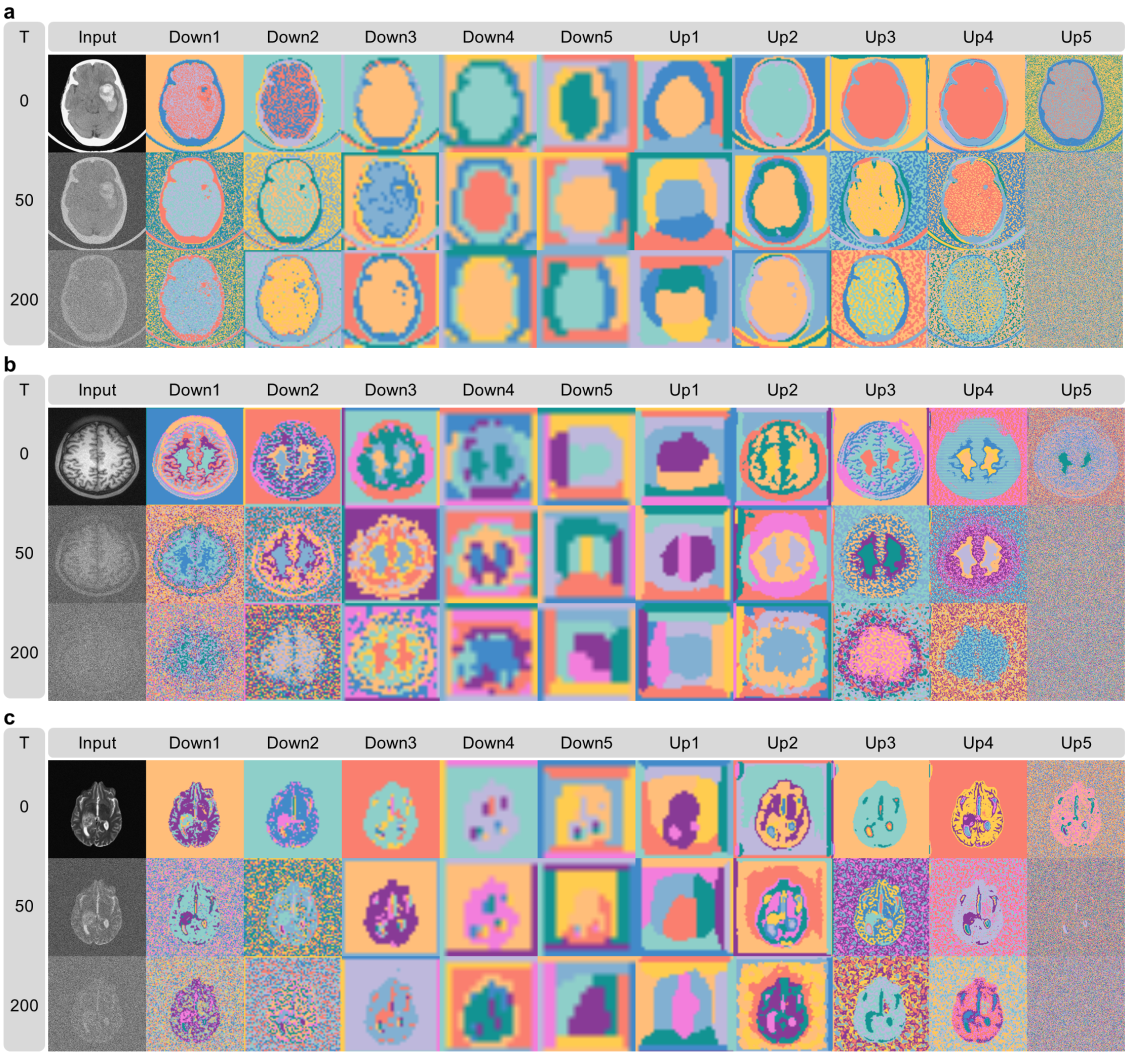


**Supplementary Figure 6**

**The visualization of features extracted by Brainfound and clustered by k-means.** The display includes examples from the ten intermediate convolutional layers of both the encoder (Down1 to Down5) and decoder (Up1 to Up5) modules at specific diffusion steps (T=0, T=50, T=200). The first column shows input images with varying Gaussian noise levels. Subsequent columns showcase classification maps derived from the k-means clustering of features, each with 10 cluster centers, where each color represents a different cluster category. Comparative analysis of features across layers indicates that lower-level feature clustering provides a more precise distinction of anatomical structures. As features are processed in higher layers, they become more abstract and contain less noise. During upsampling, distinct anatomical structures are reconstructed. When comparing feature clustering at different time steps for high Gaussian noise levels, such as at T=200, Brainfound also successfully reconstructs the general structure of the brain, indicating its acquired prior knowledge from the data. **a**, using brain CT images as the input. **b**, using brain T1WI as the input. **c**, using brain T2WI as the input.

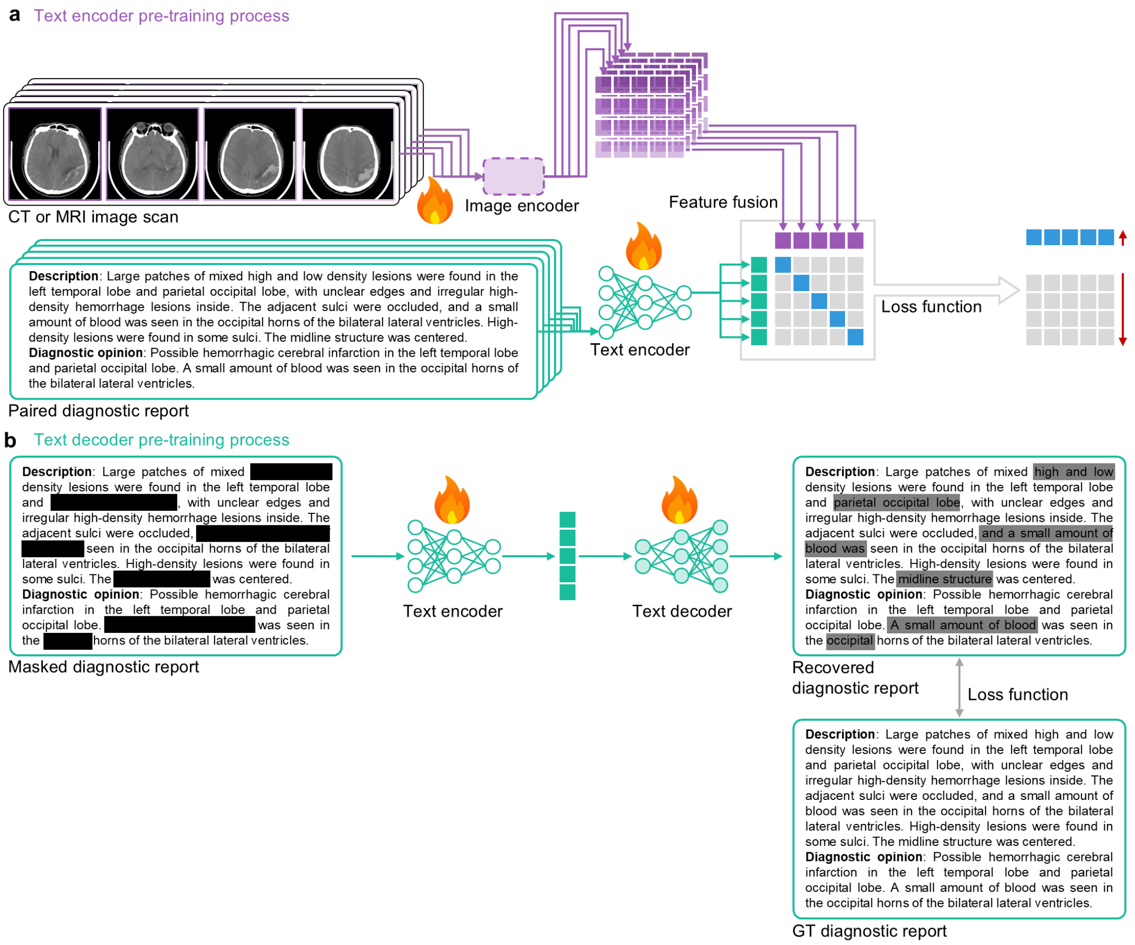


**Supplementary Figure 7**

**The alignment process between the text encoder and the image encoder of Brainfound. a,** The pre-training and alignment process of the text encoder of Brainfound. Brain imaging scans are input into the image encoder to obtain latent space features. Corresponding clinical reports are input into the text encoder to obtain textual latent space features. The two latent space features calculate cosine similarity for contrastive learning as a loss function. The alignment of the image encoder and text encoder is optimized by maximizing the loss function for paired images and reports while minimizing the loss function for unpaired images and reports. **b,** The pre-training process of text decoder of Brainfound. Clinical reports are randomly masked at the phrase or sentence level and input into the text encoder, followed by output through the text decoder, and then compared with the complete clinical reports to calculate the loss function. The parameters of the text encoder are frozen, while only the parameters of the text decoder are modified.

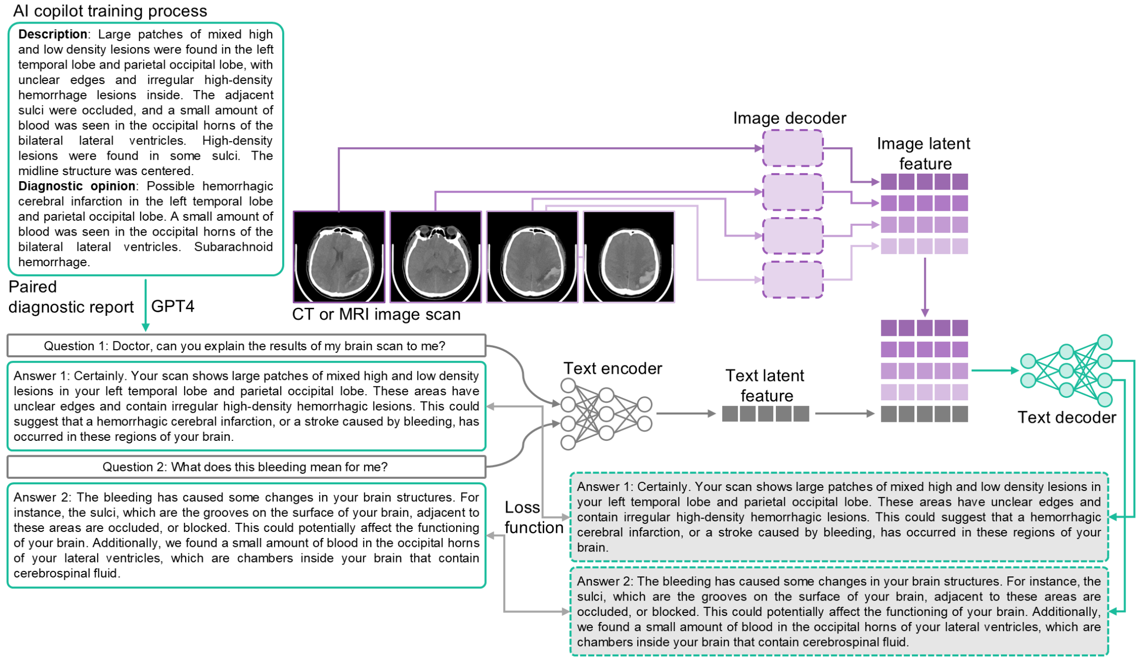


**Supplementary Figure 8**

**The training process for human-AI conversations**. For a diagnostic report, GPT-4 is used to create several rounds of open-ended conversation based on the content of the report. By employing prompt engineering, GPT-4 is instructed to formulate conversations across diverse prompt configurations, thereby enhancing the variety of conversation instructions and strengthening training robustness. In the generated conversations, questions are encoded through a text encoder to obtain text latent space features. The corresponding image scans are processed through the image encoder to extract image latent space features. These two latent space features are concatenated and fed into the text decoder. The output of the text decoder and the answers during the conversation are used to compute the loss function, which subsequently guides the optimization of both the text encoder and decoder.

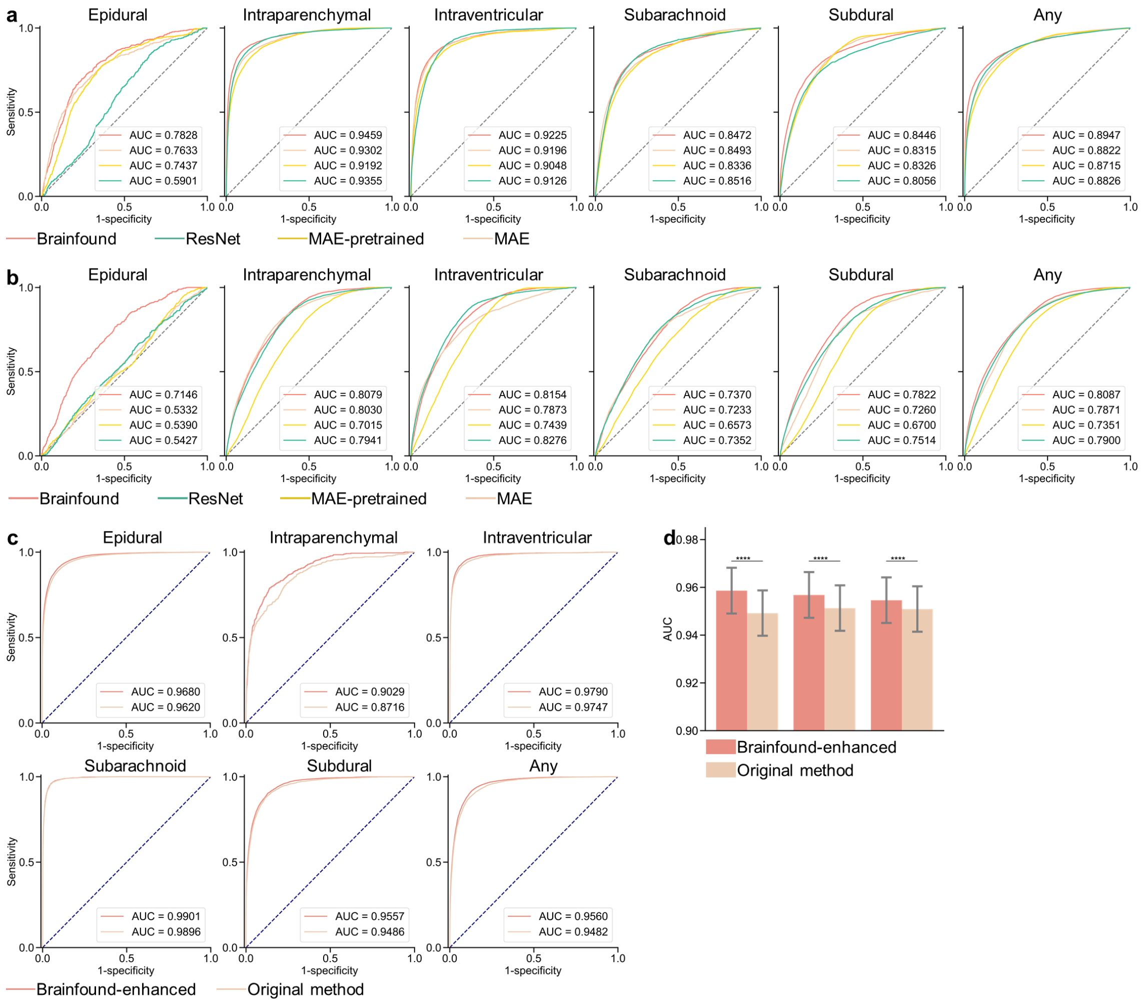


**Supplementary Figure 9**

**Performance of Brainfound on the brain hemorrhage classification task at RSNA**. **a**, The AUC curves for brain hemorrhage classification after full parameter fine-tuning of Brainfound, ResNet, MAE-pretrained, and MAE. MAE pre-trained refers to pre-training the MAE using BrainCT-3M, while MAE indicates pre-training the MAE using macro images. The categories, arranged sequentially from left to right, include Epidural, Intraparenchymal, Intraventricular, Subarachnoid, Subdural, and Any. **b**, The AUC curves for brain hemorrhage classification following fine-tuning the fixed feature extraction component of Brainfound, ResNet, MAE-pretrained, and MAE. Other details remain consistent with **a**. **c**, The AUC curve of replacing the backbone with the pre-trained Brainfound into the top-ranking solution of the RSNA classification competition. **d**, Results from substituting the backbone with the pre-trained Brainfound into the top-ranking solution of the RSNA classification competition. The testing experiments were conducted randomly three times.

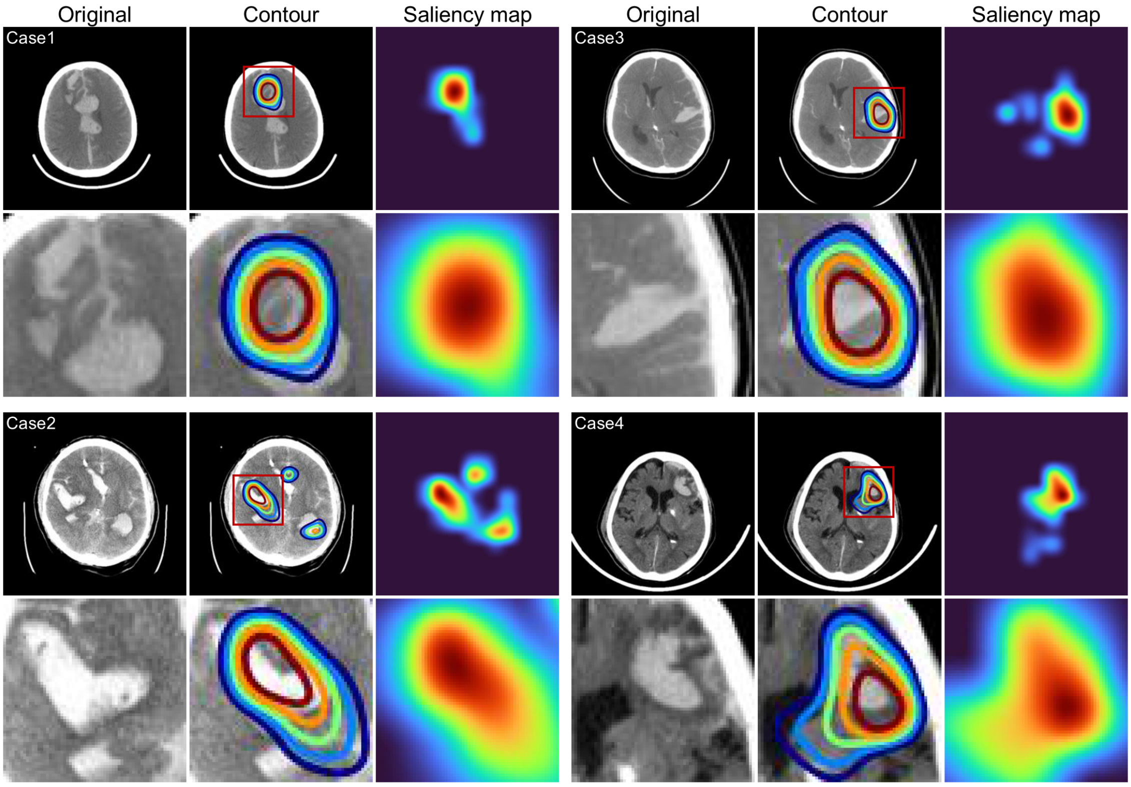


**Supplementary Figure 10**

**Saliency maps produced by Brainfound for cerebral hemorrhage classification in RSNA.** We present representative figures of saliency maps for four brain CT images from Brainfound. The brain hemorrhage CT images are located in the first and fourth columns. The second and fifth columns display the saliency contours. The saliency maps produced by Brainfound occupy the third and sixth columns. The imagery in the second and fourth rows provides an expanded view of the segments highlighted by red boxes in the first and third rows.

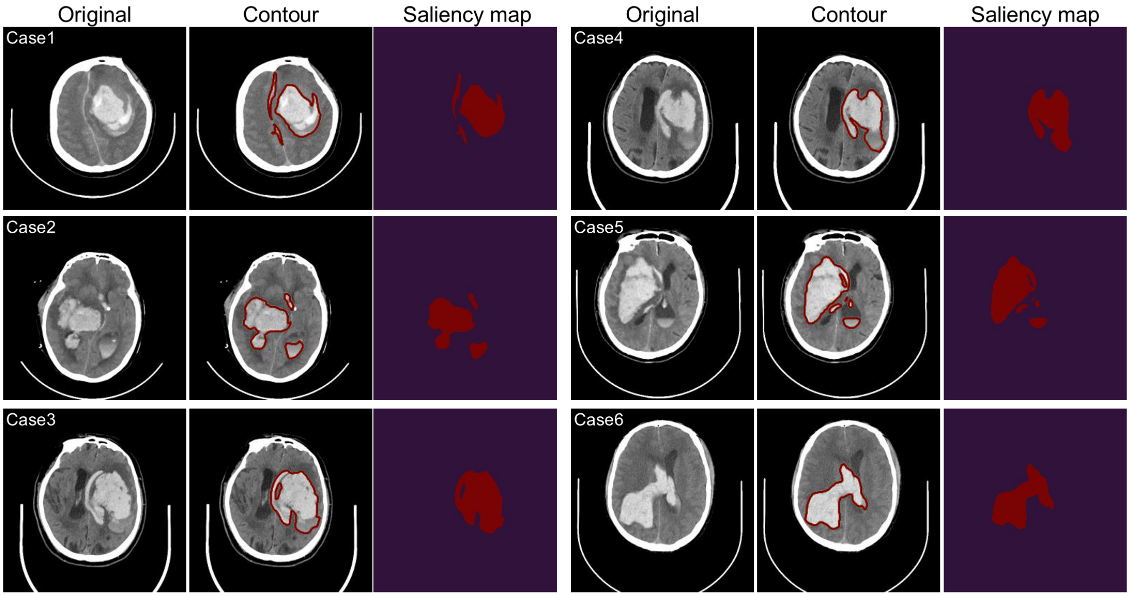


**Supplementary Figure 11**

**Saliency maps generated by Brainfound for cerebral hemorrhage segmentation in RSNA.** We presented several images in saliency maps for six brain CT images from Brainfound. The brain hemorrhage CT images appear in the first and fourth columns. The second and fifth columns display the saliency outlines. The saliency maps generated by Brainfound are in the third and sixth columns.

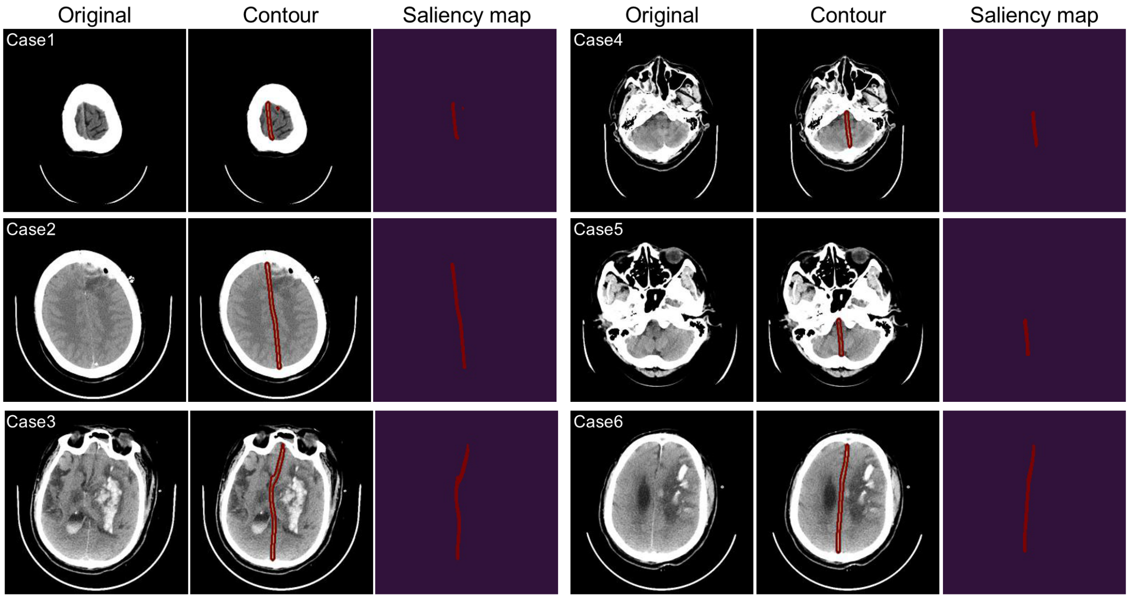


**Supplementary Figure 12**

**Saliency maps generated by Brainfound for brain midline segmentation.** We displayed a series of images in saliency maps for six brain CT scans from Brainfound. The brain CT images are located in the first and fourth columns. The second and fifth columns showcase the saliency outlines. The saliency maps created by Brainfound populate the third and sixth columns.

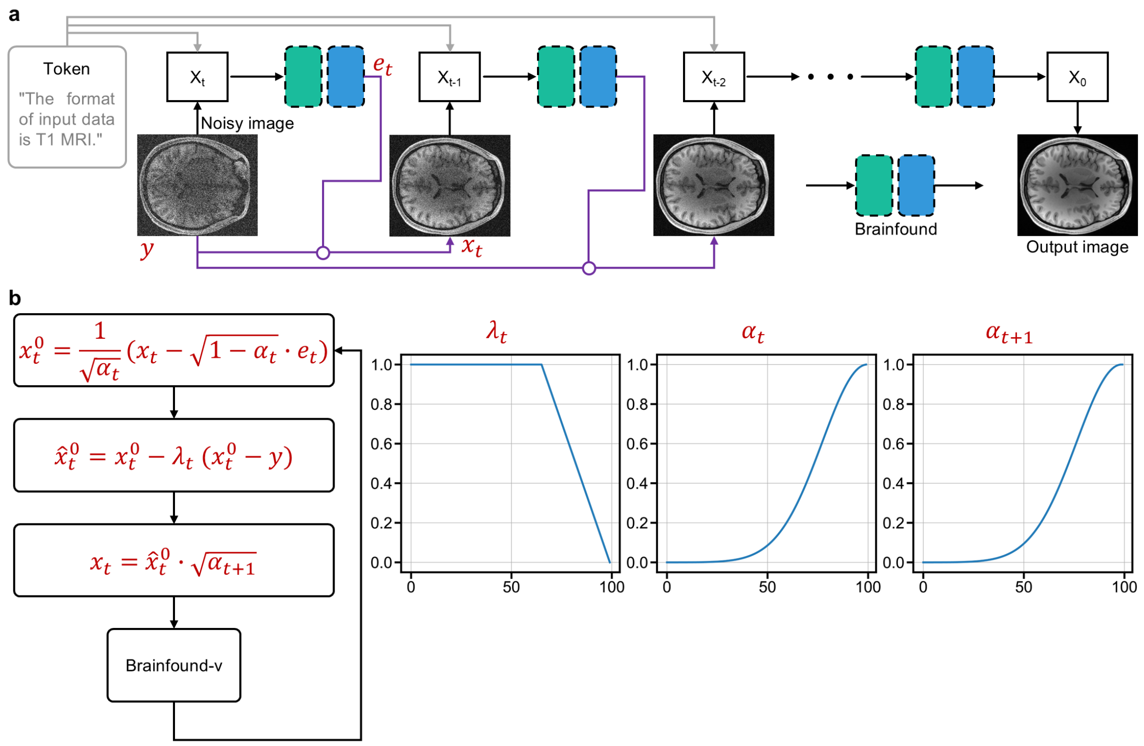


**Supplementary Figure 13**

**The zero-shot denoising mechanism of Brainfound. a**, Brainfound refines images through a step-by-step iterative process. The original noisy image is integrated into the DDPM image restoration, ultimately producing a clear image. **b**, Detailed computation of iterative steps. The three curves on the right indicate hyperparameter values throughout the iterations.

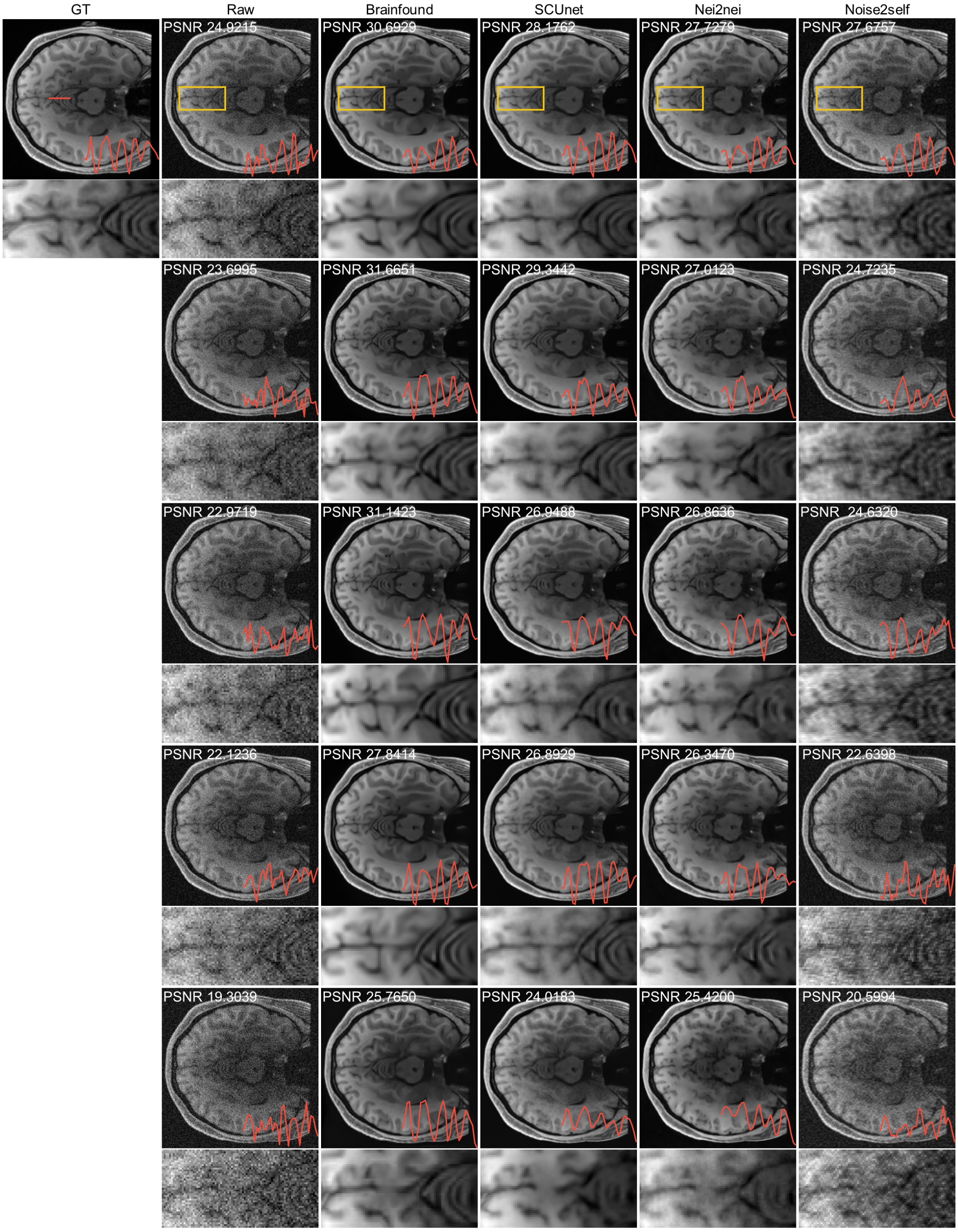


**Supplementary Figure 14**

**Comparison of the denoising results of Brainfound on 3T MRI images with simulated noise**. **a**, Arranged from left to right are the clear image, the original noisy image, the output of Brainfound, the output of SCUnet, the output of Nei2nei, and the output of Noise2self. From the first line to the fifth row, the noise intensity i progressively increases. The corresponding PSNR of the images is indicated in the upper left corner. The area within the yellow frame is magnified for display. The profile of the red line is illustrated.

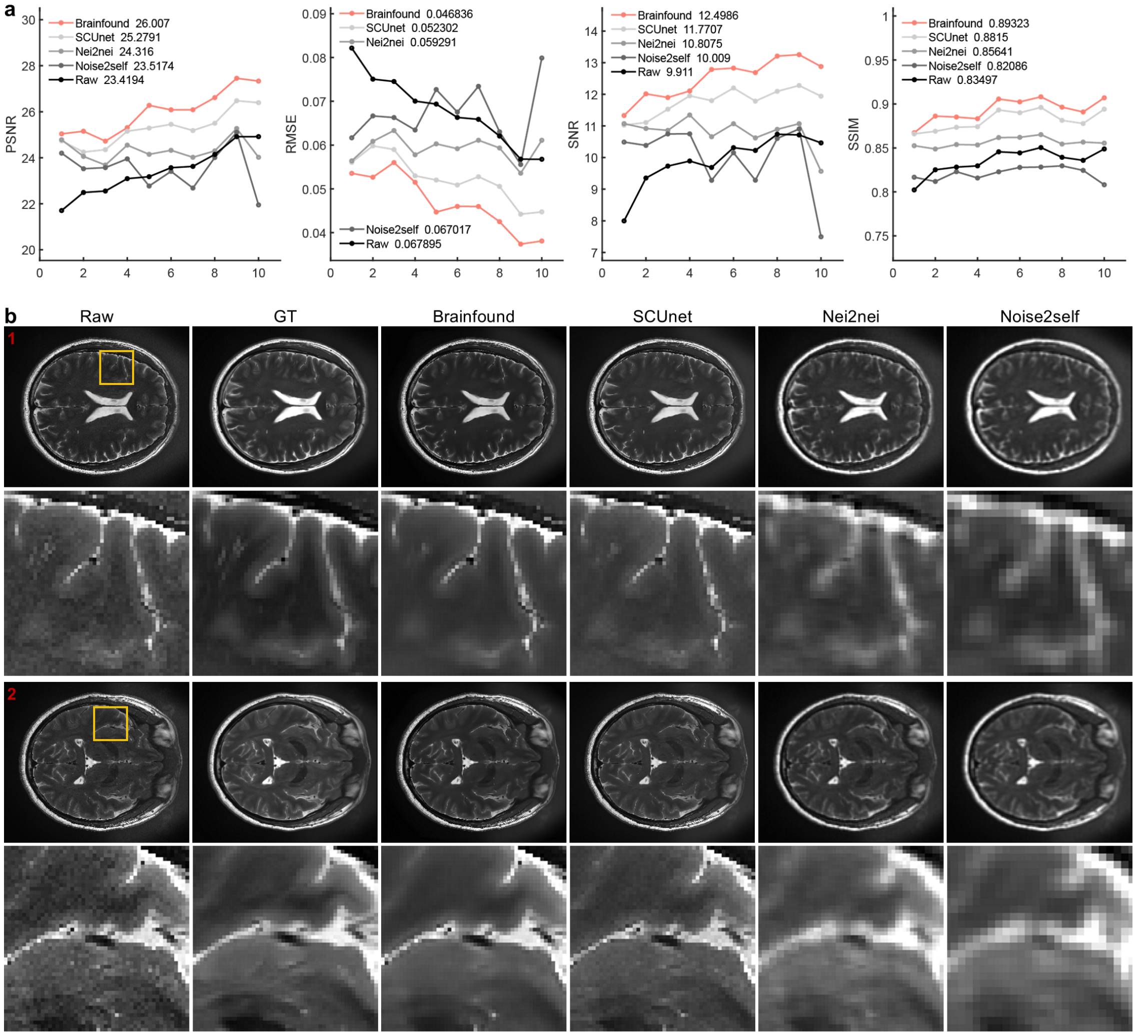


**Supplementary Figure 15**

**The enhancement of Braindfound on T2WI images captured by the 5T MRI**. **a,** The quantitative comparison of Brainfound, SCUnet, Nei2nei, and Noise2self for MRI image enhancement on the test set captured by 5T MRI (n=10) in Beijing Friendship Hospital. The scores listed from left to right are PSNR, RMSE, SNR, and SSIM. **b**, Two cases of 5T T2WI denoising. From left to right are: low SNR image, high SNR reference image, Brainfound-enhanced image, SCUnet-enhanced image, Nei2nei-enhanced image, Noise2self-enhanced image. The yellow-boxed areas in the first and third-row images are enlarged for display.

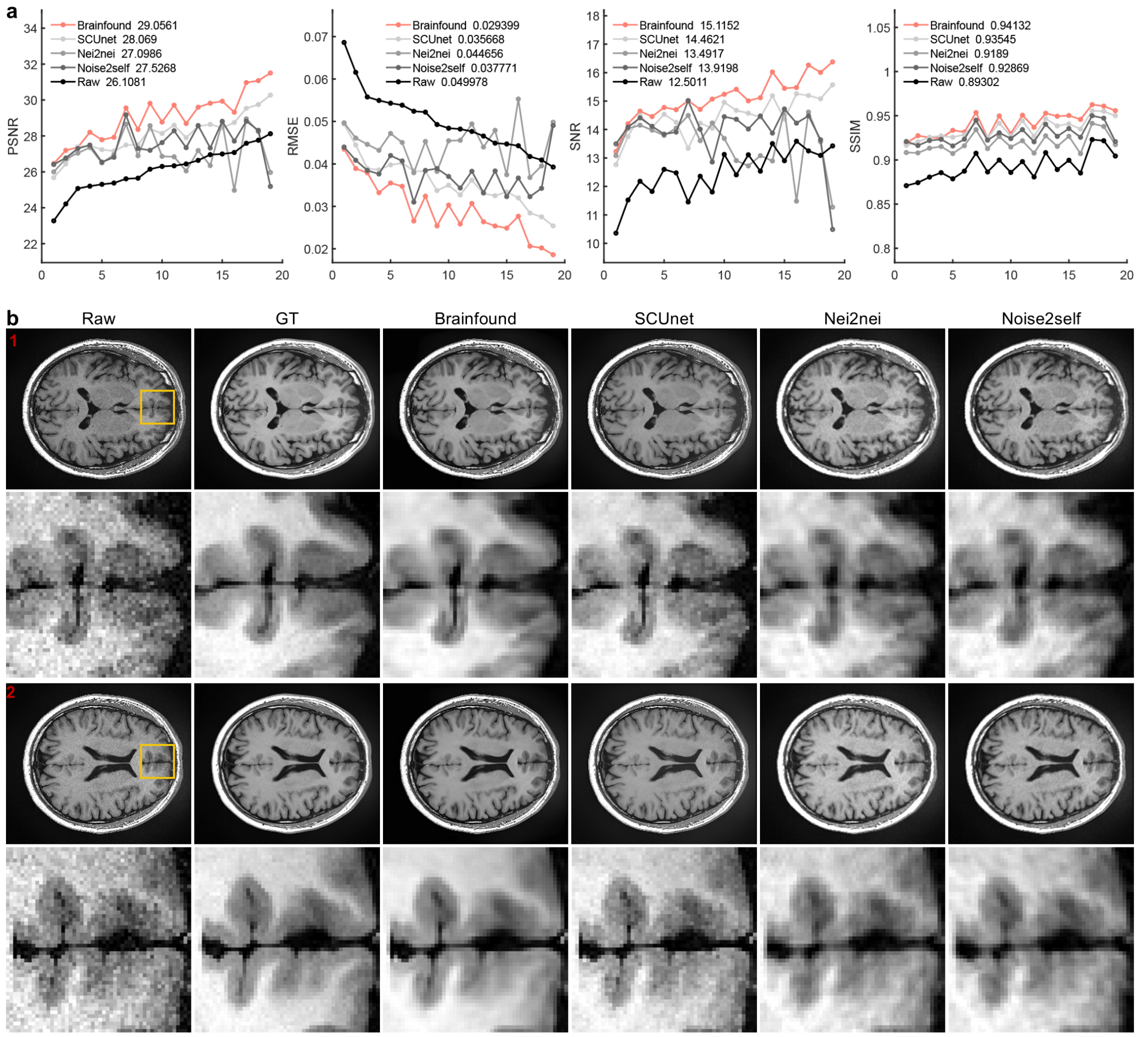


**Supplementary Figure 16**

**The enhancement of Braindfound on T1WI captured by the 5T MRI**. **a,** The comparison of Brainfound, SCUnet, Nei2nei, and Noise2self for MRI image enhancement on the external test dataset (n=19) captured by 5T MRI in Beijing Friendship Hospital. The scores displayed from left to right are PSNR, RMSE, SNR, and SSIM. **b**, Two 5T T1WI denoising cases. From left to right: low SNR image, high SNR reference image, Brainfound-enhanced image, SCUnet-enhanced image, Nei2nei-enhanced image, Noise2self-enhanced image. Enlargements of the yellow-boxed areas in the first and third-row images are provided.

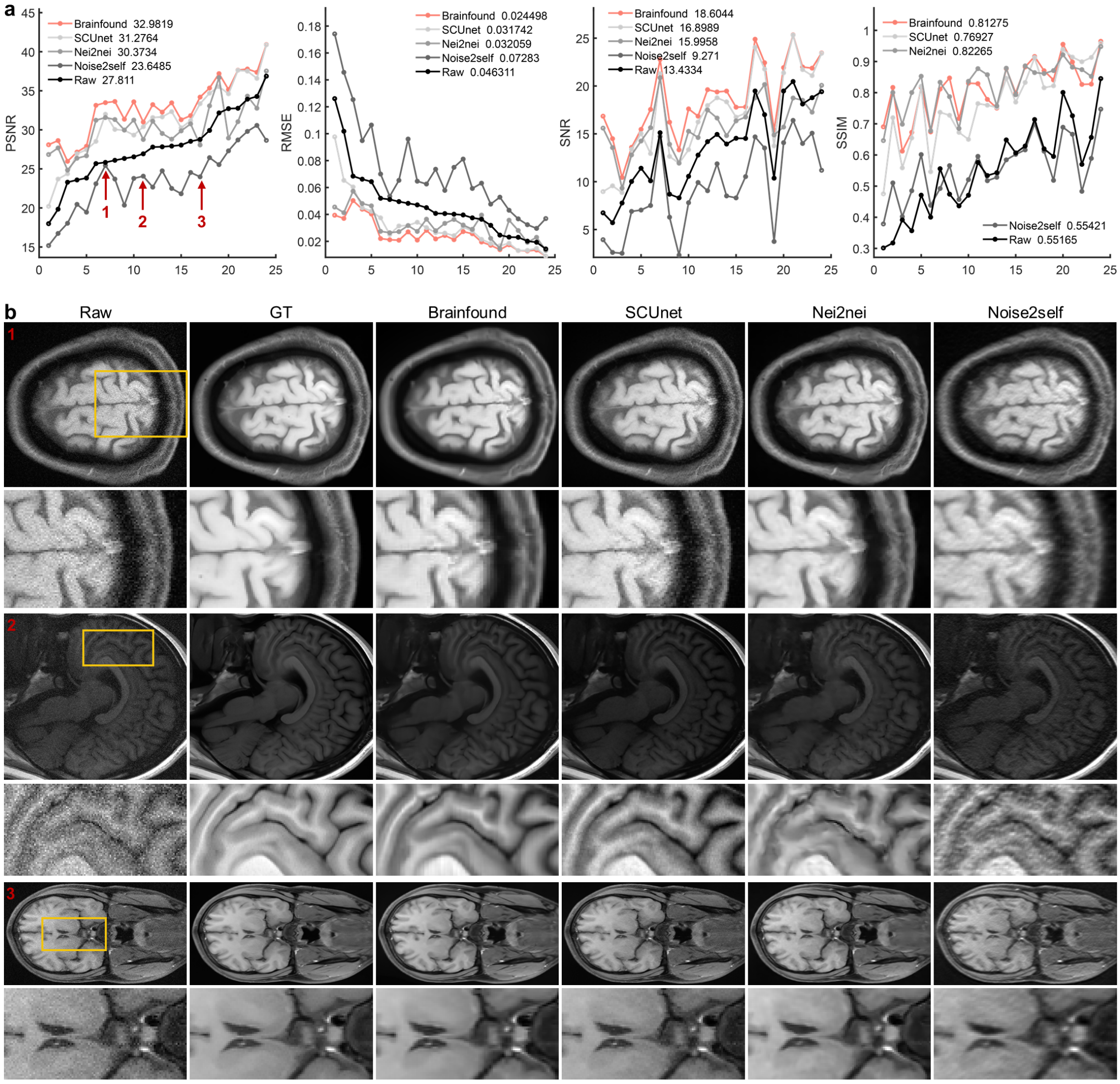


**Supplementary Figure 17**

**The enhancement of Braindfound on real-world images captured by the 5T MRI in the external center**. **a,** The quantitative comparison of Brainfound, SCUnet, Nei2nei, and Noise2self for MRI image enhancement on the external test dataset (n=25) captured by the United Imaging in Shanghai. The scores displayed from left to right are PSNR, RMSE, SNR, and SSIM. **b,** The images of the three cases marked with arrows on the curve in **a**. Displayed from left to right are the original noisy image, the high SNR GT image, the image enhanced by Brainfound, the image enhanced by SCUnet, the image enhanced by Neighbor2neighbor, and the image enhanced by Noise2self. The section outlined in yellow is enlarged for display.

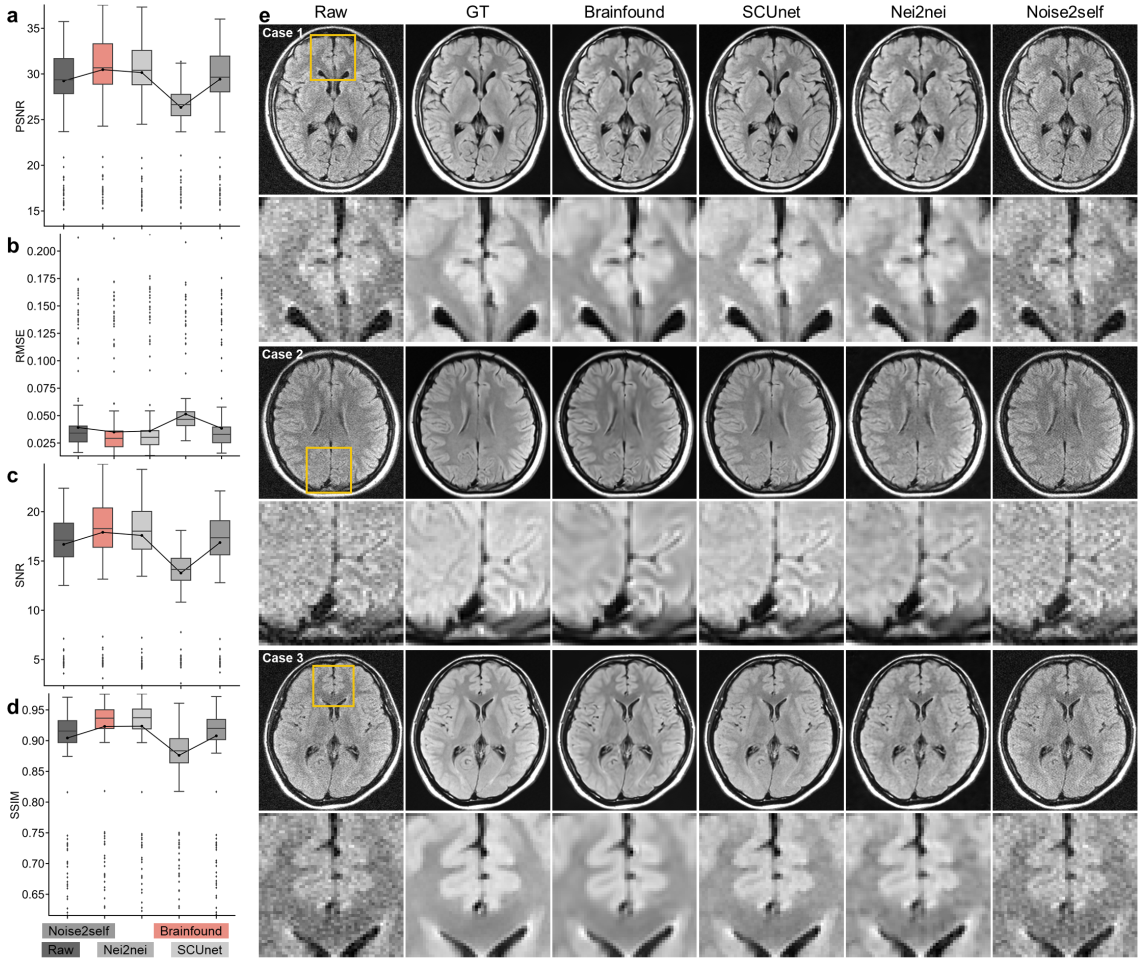


**Supplementary Figure 18**

**The comparison of zero-shot enhancement results on 0.3T FLAIR**. **a-d**, Quantitative comparison of image enhancement results using four methods: Brainfound, Noise2self, Neighbor2neighbor, SCUnet (with n=450). PSNR, RMSE, SNR, and SSIM were evaluated respectively. Brainfound achieved the best scores in all metrics except SSIM. **e**, Three typical denoising results are displayed. From left to right: the original image, high SNR reference image, Brainfound-enhanced image, SCUnet-enhanced image, Neighbor2neighbor-enhanced image, and Noise2self-enhanced image. The yellow-boxed area is enlarged for display.

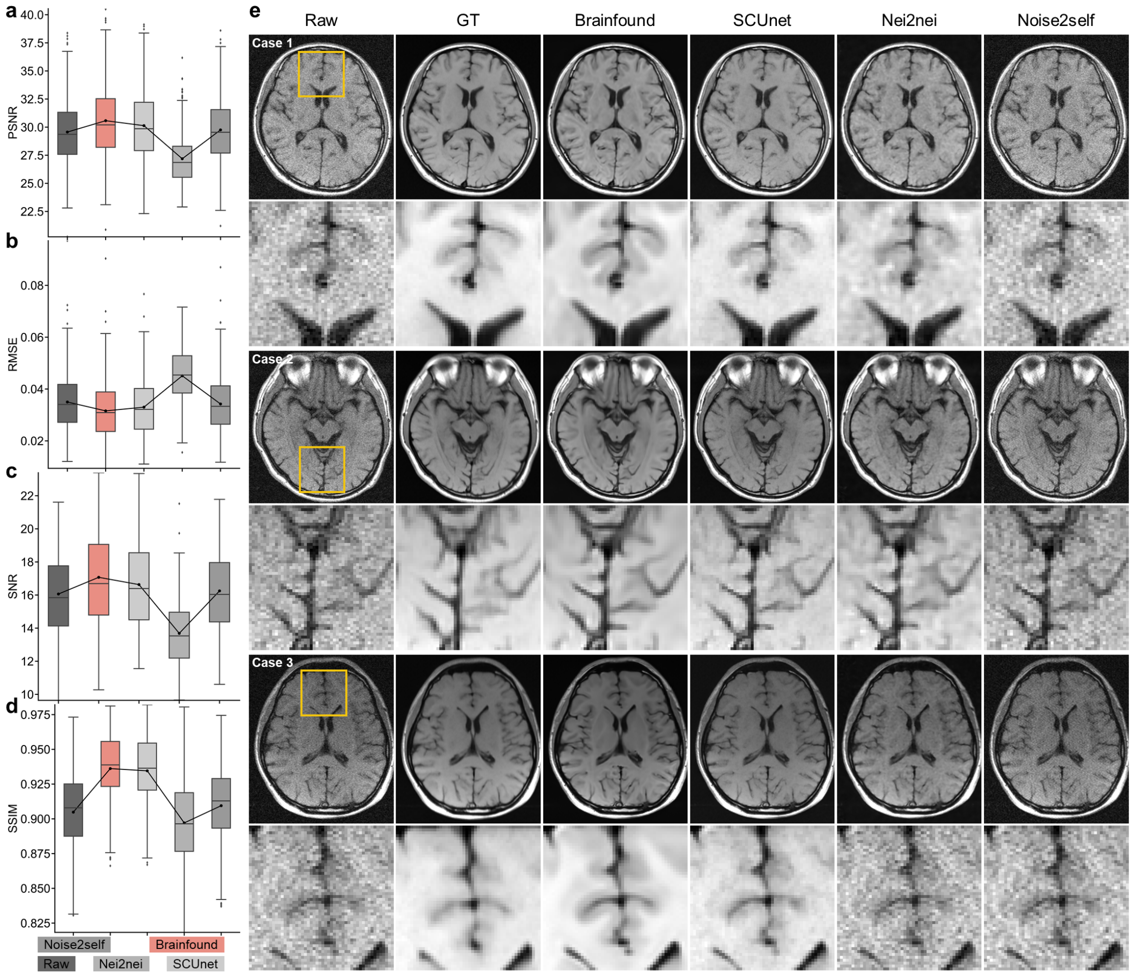


**Supplementary Figure 19**

**The comparison of zero-shot enhancement results on 0.3T T1WI**. **a-d**, Quantitative comparison of image enhancement results using four methods: Brainfound, Noise2self, Neighbor2neighbor, SCUnet (with n=450). PSNR, RMSE, SNR, and SSIM were computed respectively. Brainfound achieved the best scores in all metrics. **e**, Three typical denoising results are displayed. From left to right: the original image, high SNR reference image, Brainfound-enhanced image, SCUnet-enhanced image, Neighbor2neighbor-enhanced image, and Noise2self-enhanced image. The yellow-boxed area is enlarged for display.

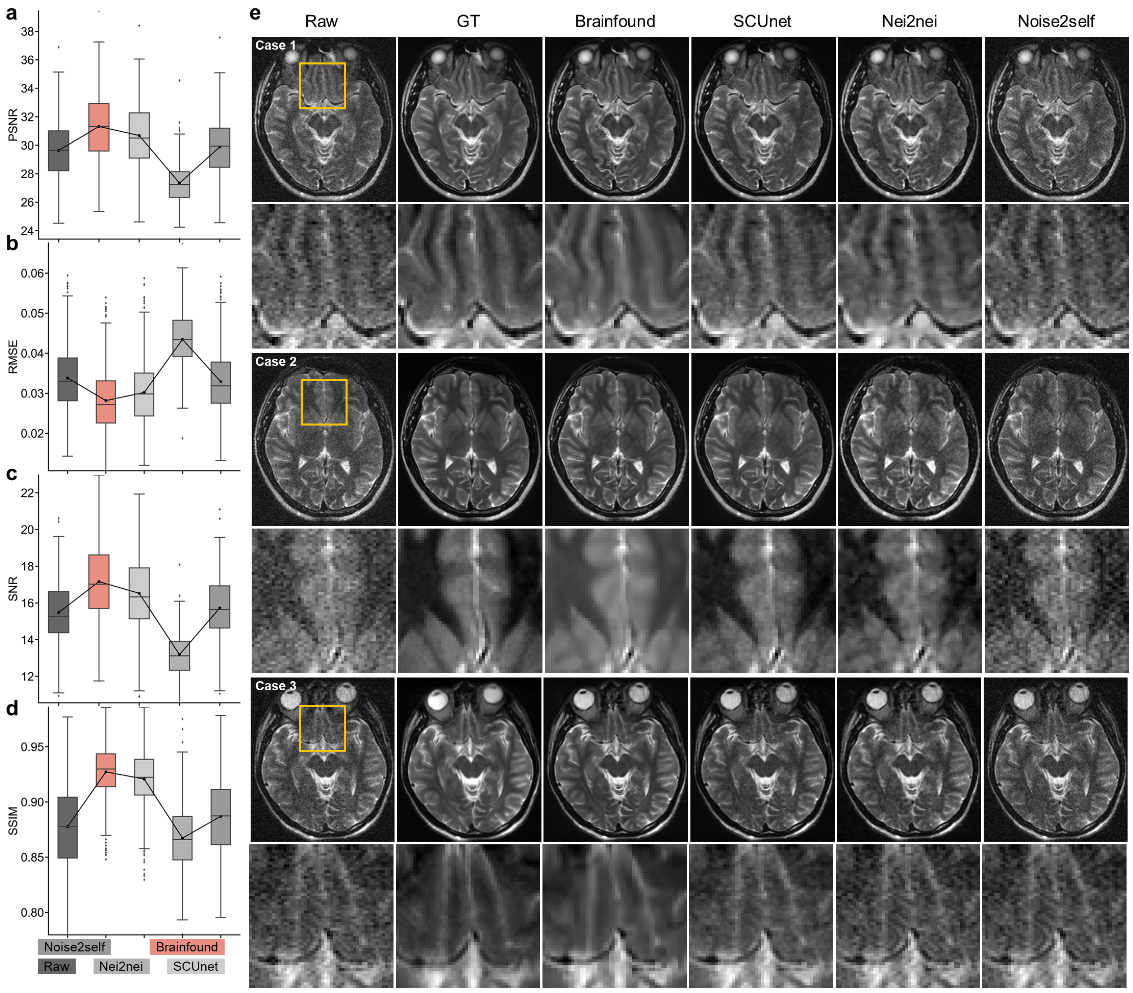


**Supplementary Figure 20**

**The comparison of zero-shot enhancement results on 0.3T T2WI**. **a-d**, Quantitative comparison of image enhancement results using four methods: Brainfound, Noise2self, Neighbor2neighbor, SCUnet (with n=450). PSNR, RMSE, SNR, and SSIM were calculated respectively. Brainfound achieved the best scores in all metrics. **e**, Three typical denoising results are displayed. From left to right: the original image, high SNR reference image, Brainfound-enhanced image, SCUnet-enhanced image, Neighbor2neighbor-enhanced image, and Noise2self-enhanced image. The yellow-boxed area is enlarged for display.

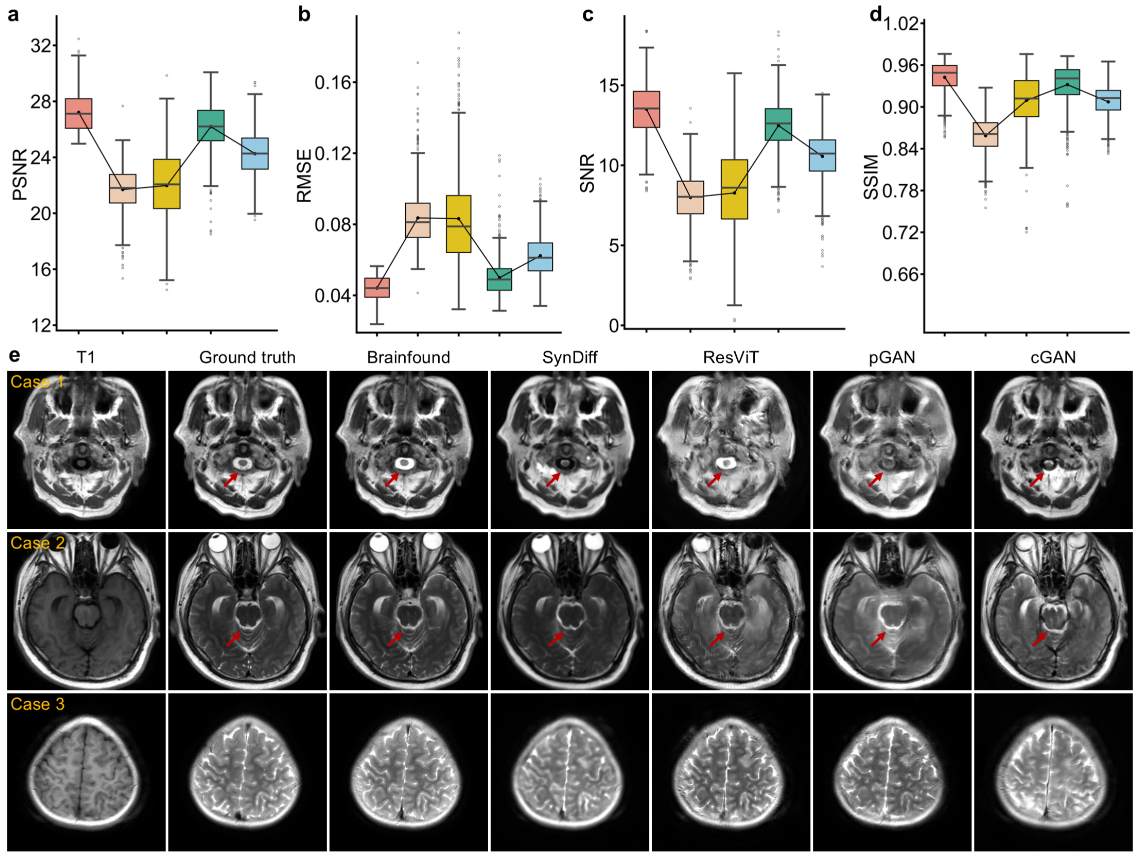


**Supplementary Figure 21**

**The performance of Brainfound in translation MRI T1WI to T2WI**. **a-d**, The quantitative evaluation of image modality translation with Brainfound, SynDiff, ResViT, pGAN, and cGAN (n=1936). Metrics including PSNR, RMSE, SNR, and SSIM were computed, and Brainfound consistently outperformed the other methods across all metrics. **e**, Three cases of T1WI-to-T2WI Image translation via five methods. From left to right: original T1WI, paired T2WI, results from Brainfound, SynDiff, ResViT, pGAN, and cGAN. In case 1, Brainfound accurately identifies the cerebrospinal fluid (CSF) around the spinal cord that shows low signal intensity on T1WI and converts these areas into high signal intensity on generated T2WI. In case 2, Brainfound generated a clearer image in which the pons and their surrounding structures look sharp and have good contrast. In case 3, Brainfound achieves a higher resolution in conversion tasks, while other methods result in slightly blurry images.

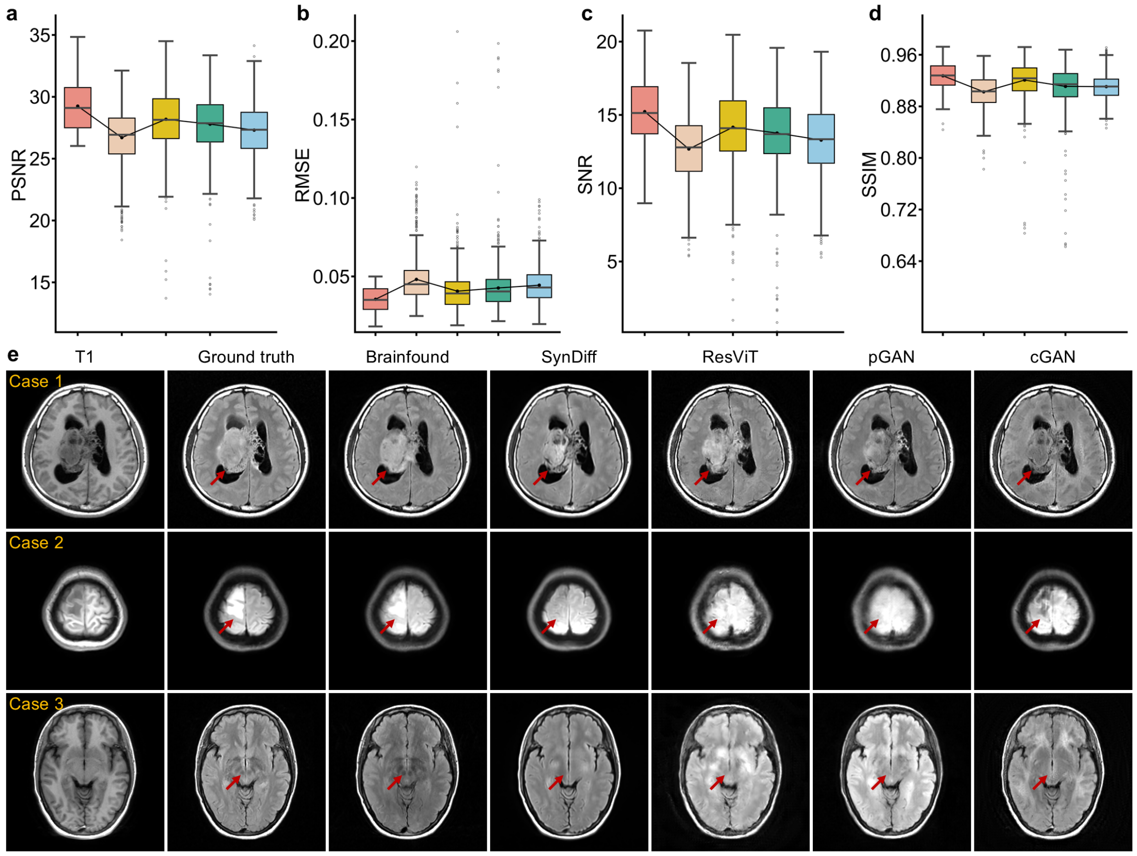


**Supplementary Figure 22**

**The performance of Brainfound in translation MRI T1WI to FLAIR**. **a-d**, Image modality translation was assessed quantitatively with Brainfound, SynDiff, ResViT, pGAN, and cGAN (n=1936). Brainfound showed superior performance across all metrics, including PSNR, RMSE, SNR, and SSIM. **e**, Three cases of T1WI-to-FLAIR image translation via five methods. In case 1, Brainfound more effectively transformed the situation of the tumor. In case 2, Brainfound accurately converted the edema lesion with low signal intensity on T1WI into high signal intensity on FLAIR. In the case 3, Brainfound accurately generated the distinct darkened areas of the red nucleus and substantia nigra on the FLAIR image.

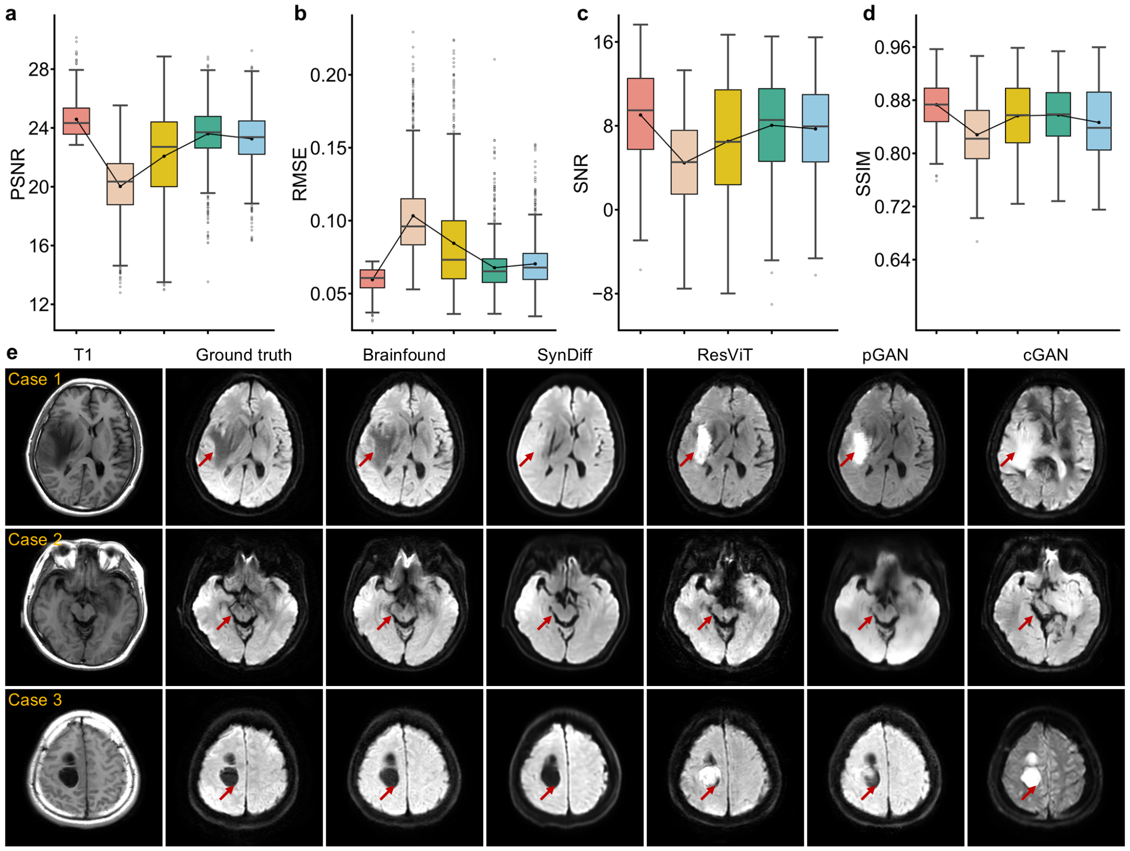


**Supplementary Figure 23**

**The performance of Brainfound in translation MRI T1WI to standard-b-value DWI**.

**a-d**, A quantitative assessment of T1WI to standard-b-value DWI image translation results was carried out using five methods: Brainfound, SynDiff, ResViT, pGAN, and cGAN (n=1936). The metrics PSNR, RMSE, SNR, and SSIM were calculated, with Brainfound obtaining the best performance in all metrics. **e**, Three cases of T1WI to standard-b-value DWI Image translation via five methods. For case 1 and case 3, the peritumoral vasogenic edema and the cystic lesions typically do not show restricted diffusion (high signal) on DWI images, and Brainfound accurately identified the edema and cystic regions and output the images with corresponding hypointense lesions. In case 2, the images generated by Brainfound exhibit less distortion in the slices near the base of the skull, and the depiction of the brainstem, ambient cisterns, and medial temporal lobes is clearer and matches the ground truth better than the other models.

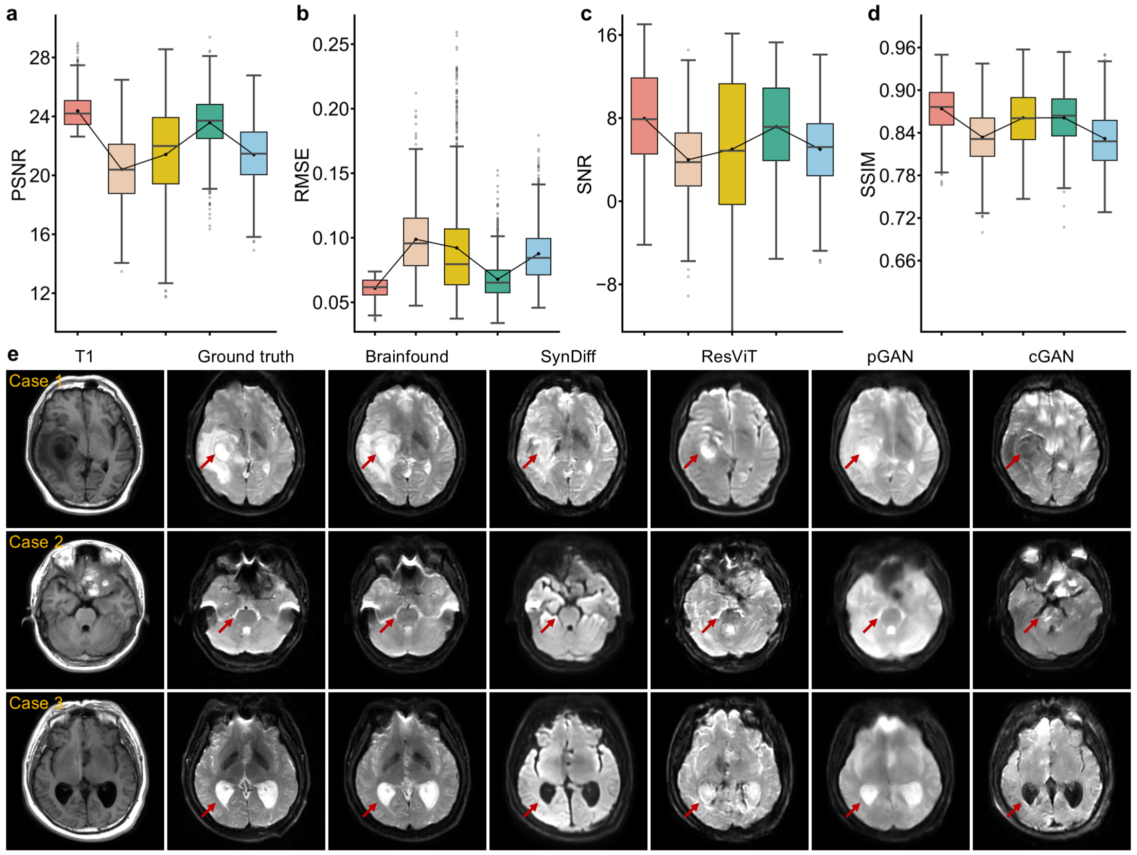


**Supplementary Figure 24**

**The performance of Brainfound in translation MRI T1WI to low-b-value DWI**. **a-d,** A quantitative comparison was made of T1WI to low-b-value DWI image translation results using five methods: Brainfound, SynDiff, ResViT, pGAN, and cGAN (n=1936). PSNR, RMSE, SNR, and SSIM were calculated individually. Brainfound achieved the highest scores in all metrics. **e**, Three cases of T1WI to low-b-value DWI Image translation via five models. As shown by the red arrows, the vasogenic edema and cystic lesions (case 1) and the CSF in the lateral ventricles (case 3) were accurately transformed into the high signal from the low signal on the original T1WI by Brainfound. In case 2, Brainfound provides a conversion that is closest to the ground truth.

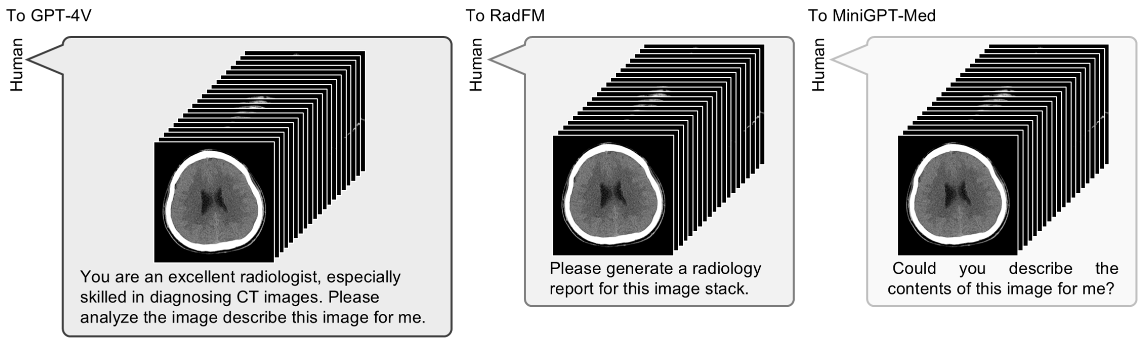


**Supplementary Figure 25**

**The prompts utilized during the report generation process**. From left to right: the prompt used for GPT-4V, the prompt used for RadFM, and the prompt used for MiniGPT-Med.

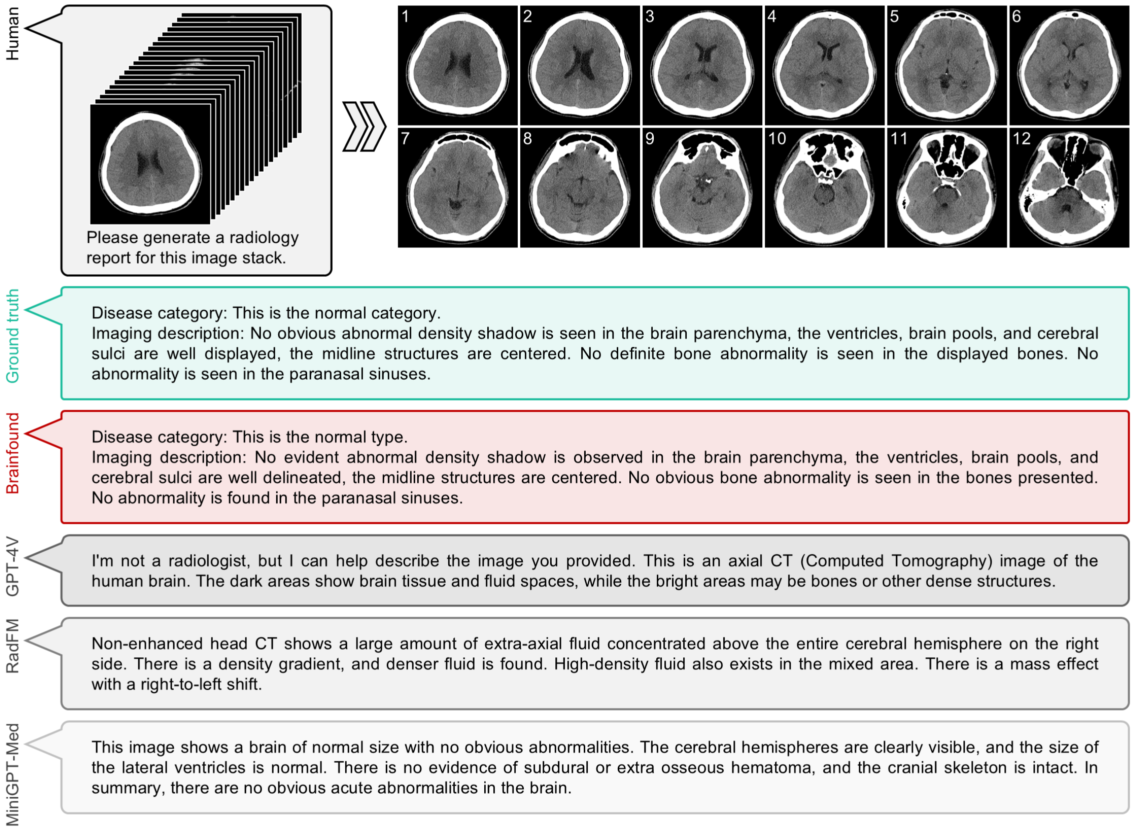


**Supplementary Figure 26**

**The report generation result of a normal brain CT scan**. The upper portion of the display illustrates the input CT scan and the prompt employed in the examination. On the right, there are 12 typical brain CT images from the scan. Beneath this, reports crafted by an experienced doctor, along with those formulated by Brainfound, GPT4-V, RadFM, and MiniGPT-Med, are showcased. The narrative of the CT scan in the report generated by RadFM is somewhat confusing. The report issued by GPT-4V is not in compliance with clinical standards.

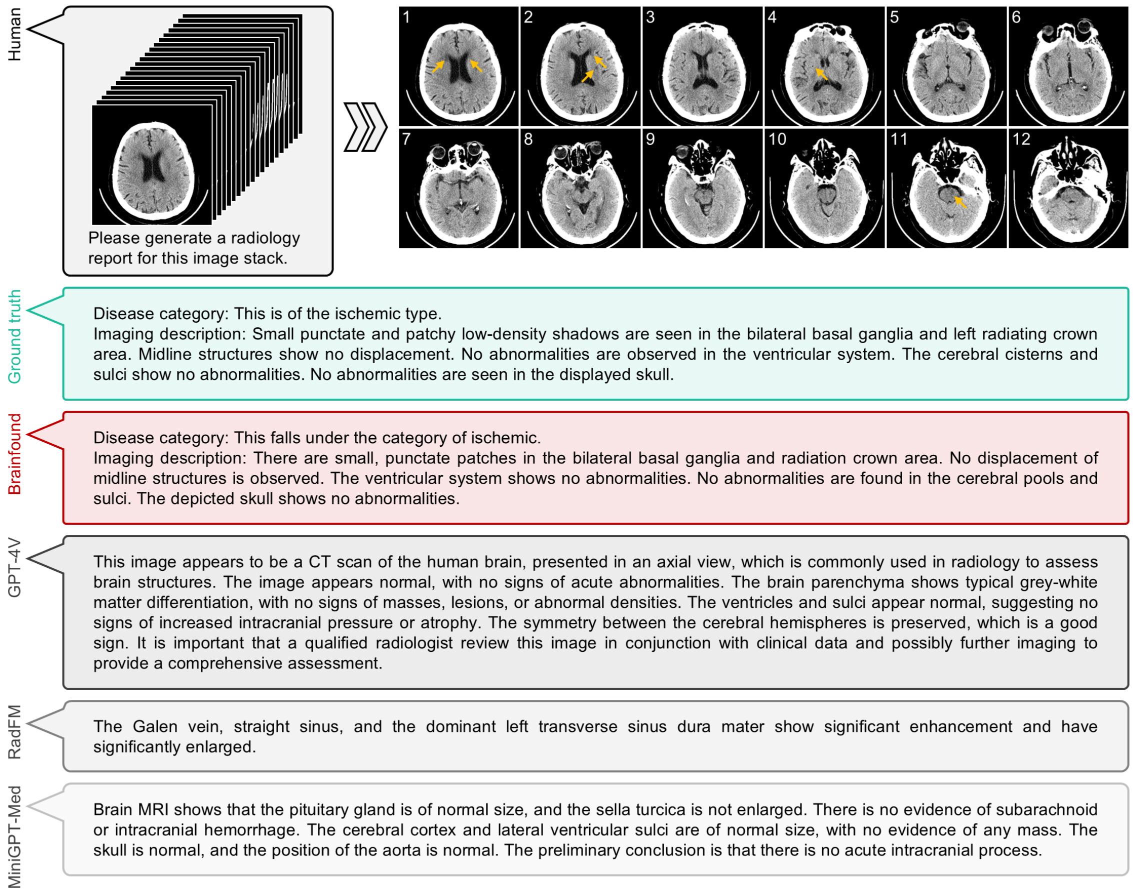


**Supplementary Figure 27**

**The report generation result of an ischemic brain CT scan**. The upper portion of the display illustrates the input CT scan and the prompt used in the examination. On the right, there are 12 typical brain CT images from the scan. Subsequently, reports crafted by an experienced doctor, along with those formulated by Brainfound, GPT4-V, RadFM, and MiniGPT-Med, are displayed. The report generated by RadFM is deficient in adequately describing the CT scan. The report issued by MiniGPT-Med incorrectly characterizes the images as an MRI sequence.

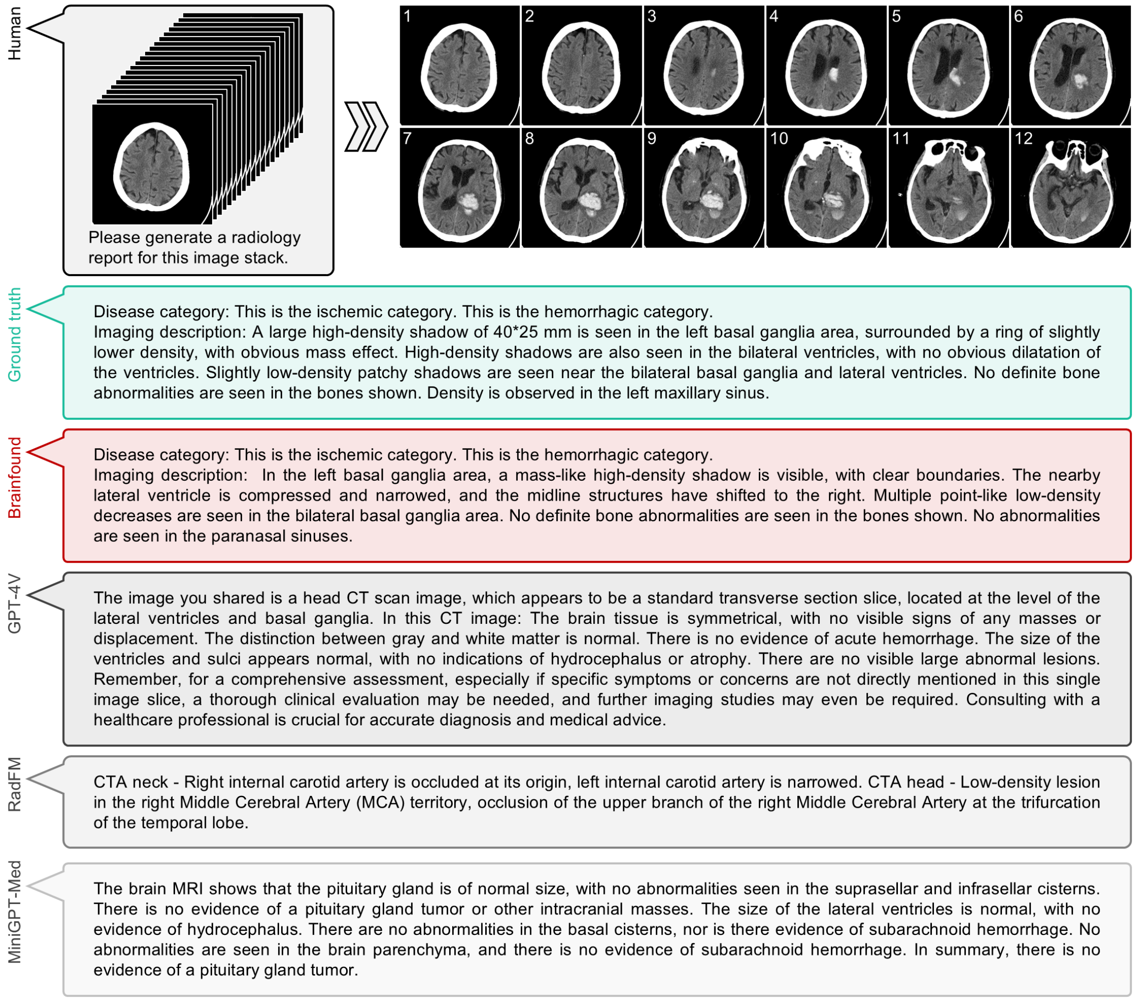


**Supplementary Figure 28**

**The report generation result of a hemorrhage brain CT scan**. The upper portion of the display illustrates the input CT scan and the prompt used in the examination. On the right, there are 12 typical CT images from the scan. Subsequently, reports crafted by an experienced doctor, along with those formulated by Brainfound, GPT4-V, RadFM, and MiniGPT-Med, are showcased. The report formulated by GPT-4V fails to enumerate the diseases present in the CT. The report generated by RadFM mischaracterizes the images as CTA imaging. The report rendered by MiniGPT-Med erroneously characterizes the CT sequence as an MRI sequence.

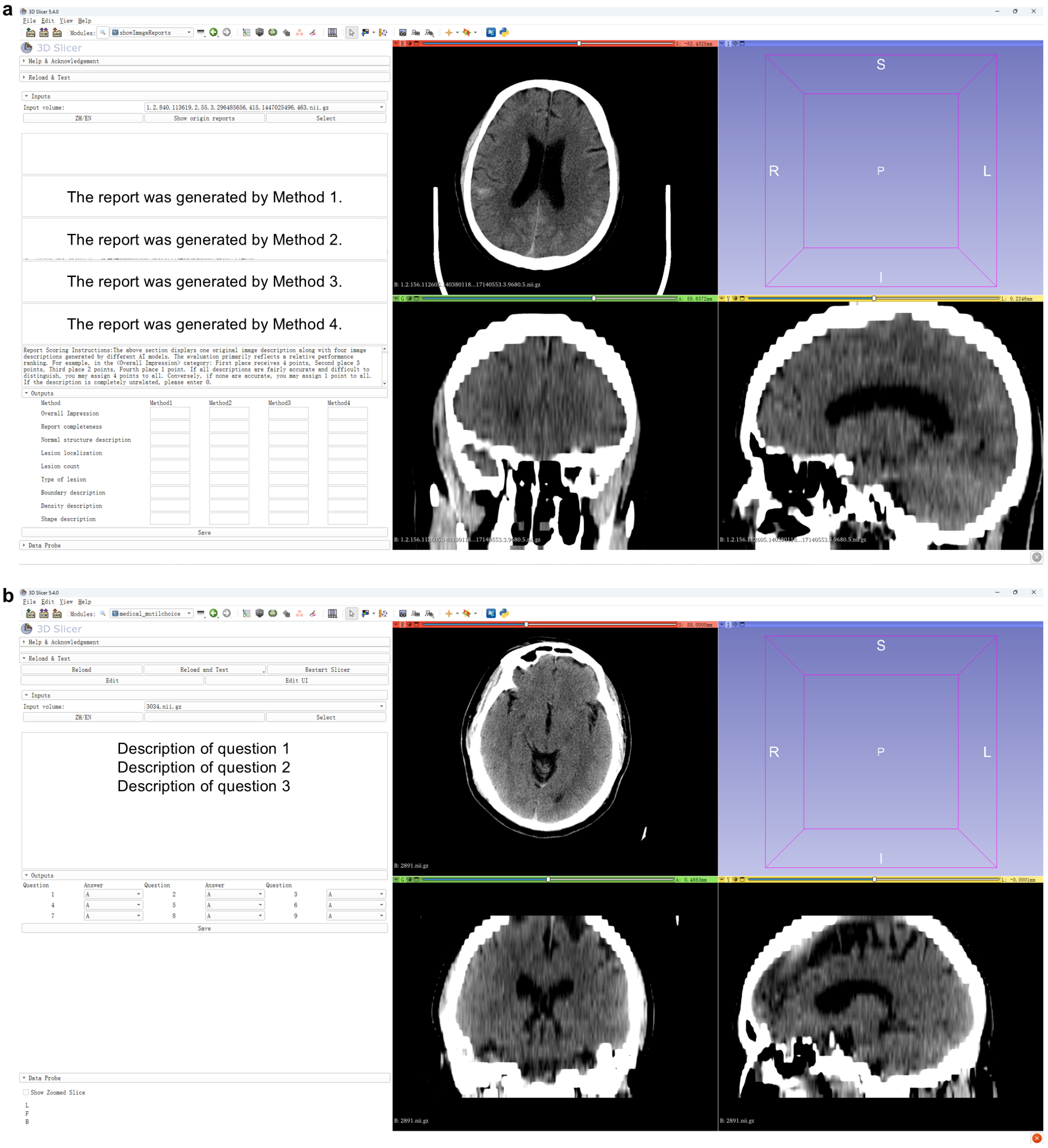


**Supplementary Figure 29**

**The report evaluation system and MCQ answering system are based on 3D Slicer.** **a**, On the right side of the graphical user interface (GUI), the brain CT scan that corresponds to the current reports is displayed in three dimensions. The left side of the interface presents medical reports generated by four methods awaiting evaluation. For reference, medical reports that have been authored by experienced doctors are available for displaying. The evaluative process involves a comprehensive assessment of report quality across nine distinct dimensions. **b**, MCQ answering system for doctors. This panel delineates three MCQs, each with multiple answer choices, where doctors are prompted to input their selections at the designated lower segment of the interface. For more details, refer to https://github.com/gingerbread000/SlicerMedicalReportGrading.

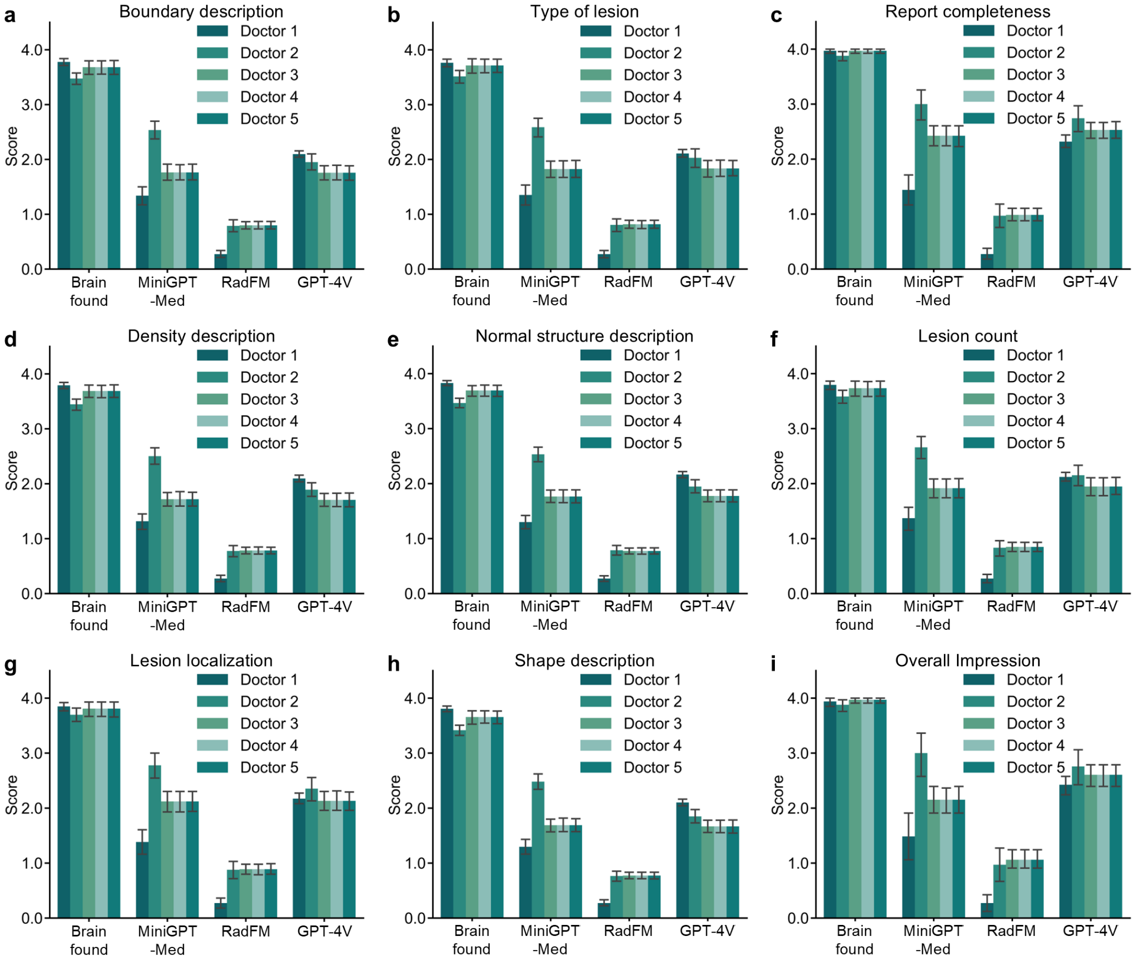


**Supplementary Figure 30**

**The results of scoring by experienced doctors on reports generated from four methods**. **a**, The scoring results of boundary description. **b**, The scoring results of the type of lesion. **c**, The scoring results of report completeness. **d**, The scoring results of density description. **e**, The scoring results of normal structure description. **f**, The scoring results of lesion count. **g**, The scoring results of lesion localization. **h**, The scoring results of shape description. **i**, The scoring results of the overall impression. The average tenure of the five doctors is 6.4 years.

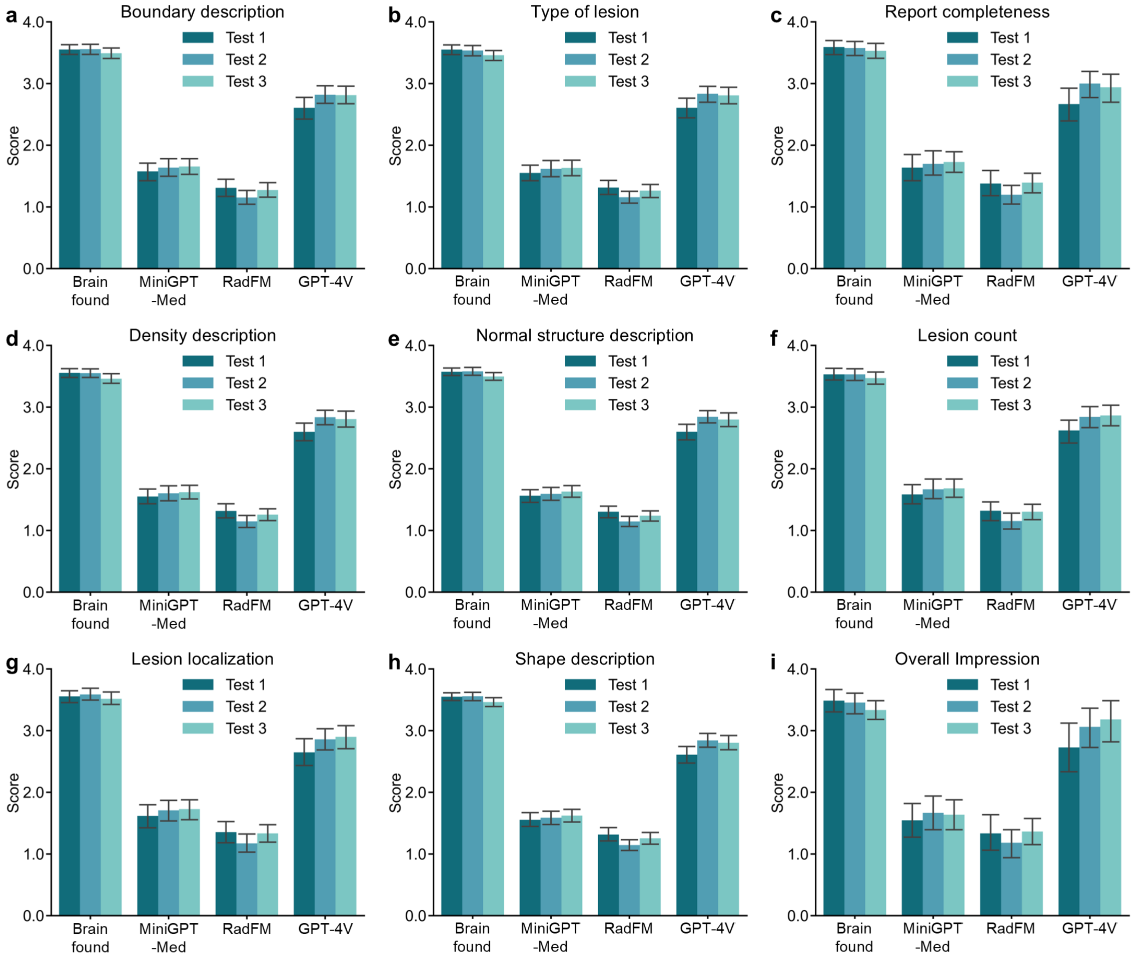


**Supplementary Figure 31**

**The results of scoring on reports generated by four methods using GPT-4**. **a**, The scoring results of boundary description. **b**, The scoring results of type of lesion. **c**, The scoring results of report completeness. **d**, The scoring results of density description. **e**, The scoring results of normal structure description. **f**, The scoring results of lesion count. **g**, The scoring results of lesion localization. **h**, The scoring results of shape description. **i**, The scoring results of the overall impression. The prompt for GPT-4 during the evaluation process is as follows: *You are an excellent radiologist, particularly skilled in determining whether a brain CT report is correct and compliant with standards. I will provide you with 5 reports, the first of which is written by a professional doctor after interpreting the CT. The other four reports are written by four different methods. I need you to score the other four reports based on the first report. Please score separately for* *the following aspects: overall impression, report completeness, normal structure description, lesion localization, lesion quantity, lesion type boundary description, density description, and shape description.*

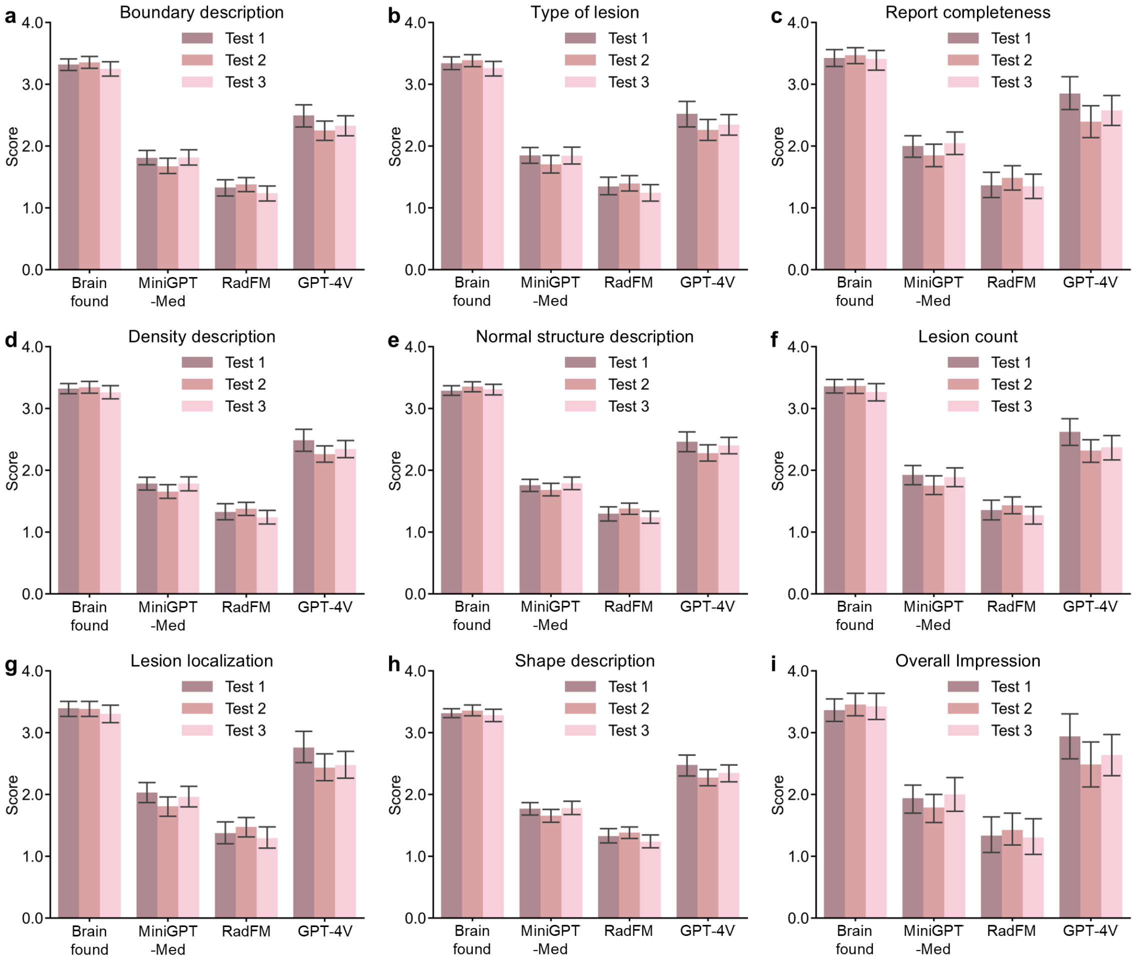


**Supplementary Figure 32**

**The results of scoring on reports generated by four methods using GPT-4o**. **a**, The scoring results of boundary description. **b**, The scoring results of type of lesion. **c**, The scoring results of report completeness. **d**, The scoring results of density description. **e**, The scoring results of normal structure description. **f**, The scoring results of lesion count. **g**, The scoring results of lesion localization. **h**, The scoring results of shape description. **i**, The scoring results of the overall impression. The prompt for GPT-4o during the evaluation process is the same as GPT-4 in Supplementary Fig. 31.

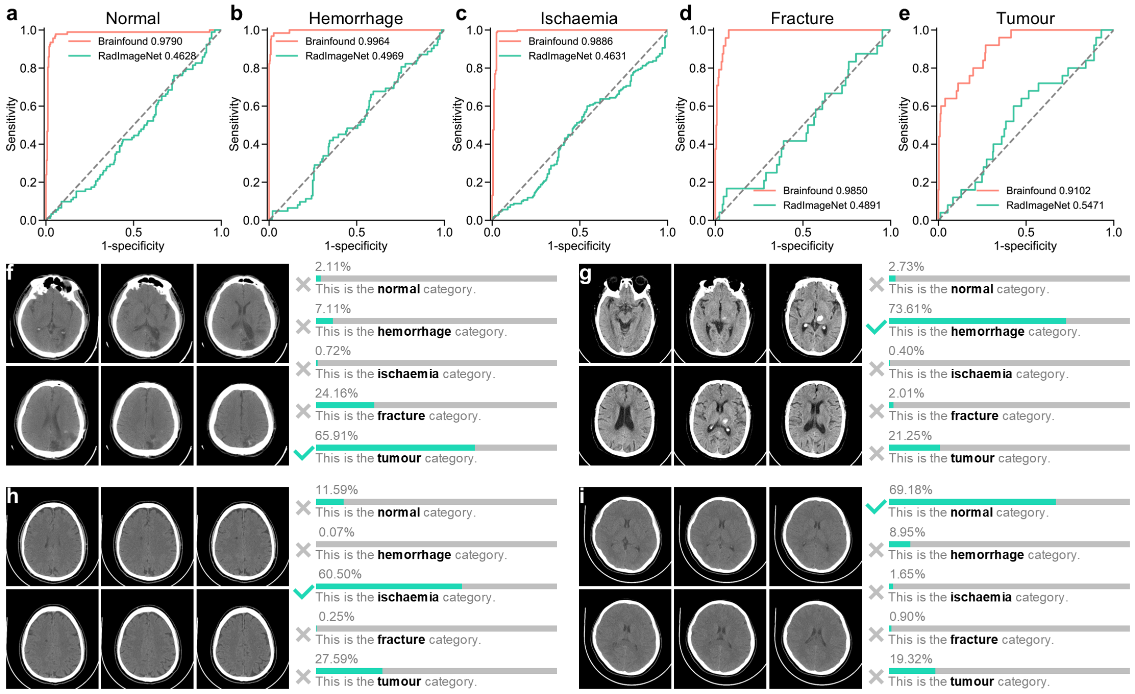


**Supplementary Figure 33**

**The zero-shot classification results of Brainfound on the external test set**. **a-e**, The zero-shot classification results of Brainfound, with RadImageNet serving as the comparison method on the external test set. The AUC curves, arranged from left to right, represent the categories of normal, hemorrhage, ischemia, fracture, and tumor. **f**, Probability outputs for tumor classification. **g**, Probability outputs for hemorrhage classification. **h**, Probability outputs for ischemia classification. **i**, Probability outputs for normal classification.

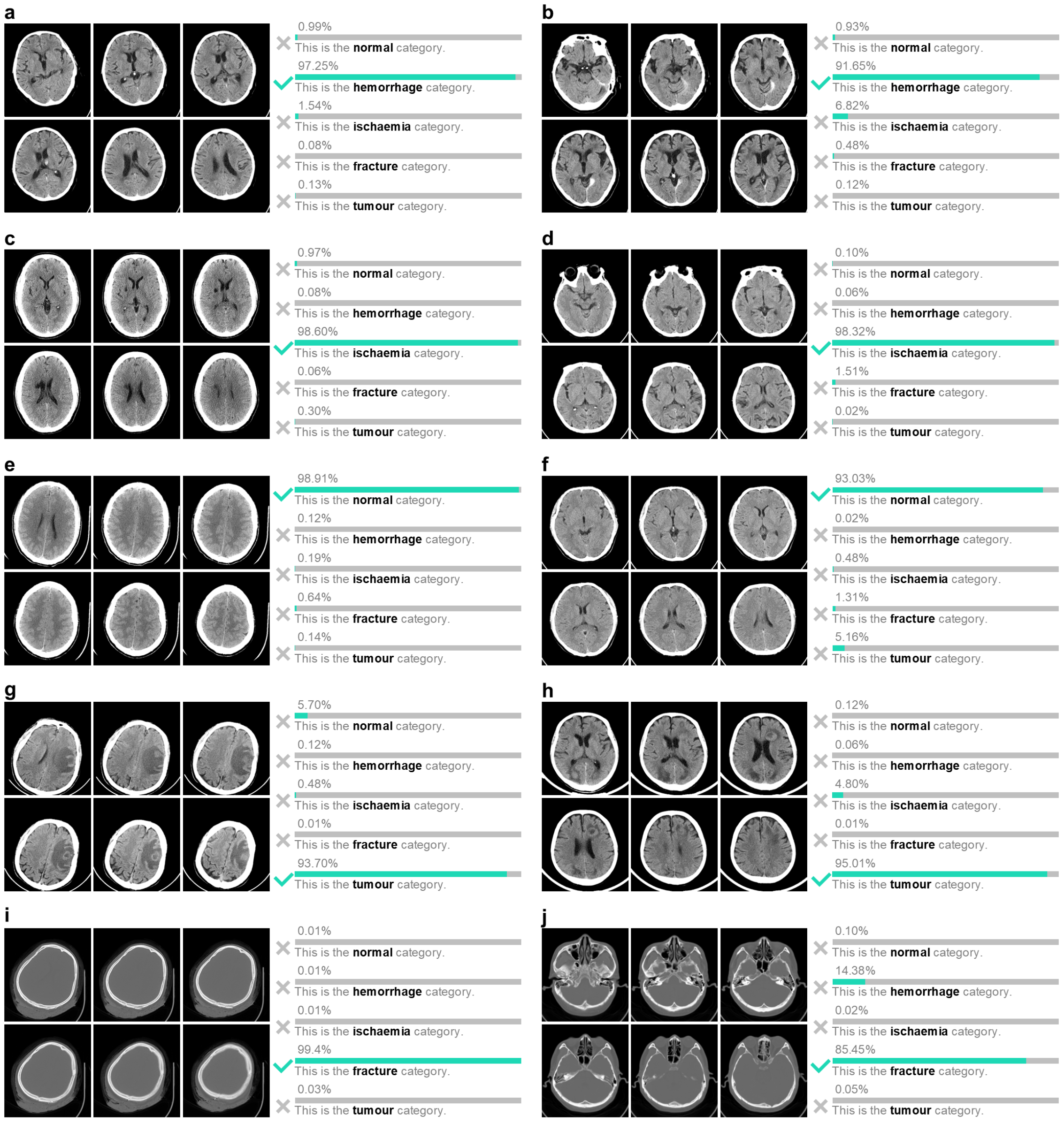


**Supplementary Figure 34**

**The zero-shot classification results of aligned image encoder and text encoder in Brainfound**. **a-b**, Two cases for the classification of brain hemorrhage types. The output probabilities are 97.25% and 91.65%, respectively. **c-d**, Two cases for the classification of brain ischemia types. The output probabilities are 98.60% and 99.35%, respectively. **e-f**, Two cases for the classification of brain normal types. The output probabilities of 98.91% and 93.03%, respectively. **g-h**, Two cases for the classification of brain tumor types. The output probabilities are 93.70% and 95.01%, respectively. **i-j**, Two cases for the classification of brain fracture types. The output probabilities are 99.40% and 85.45%, respectively.

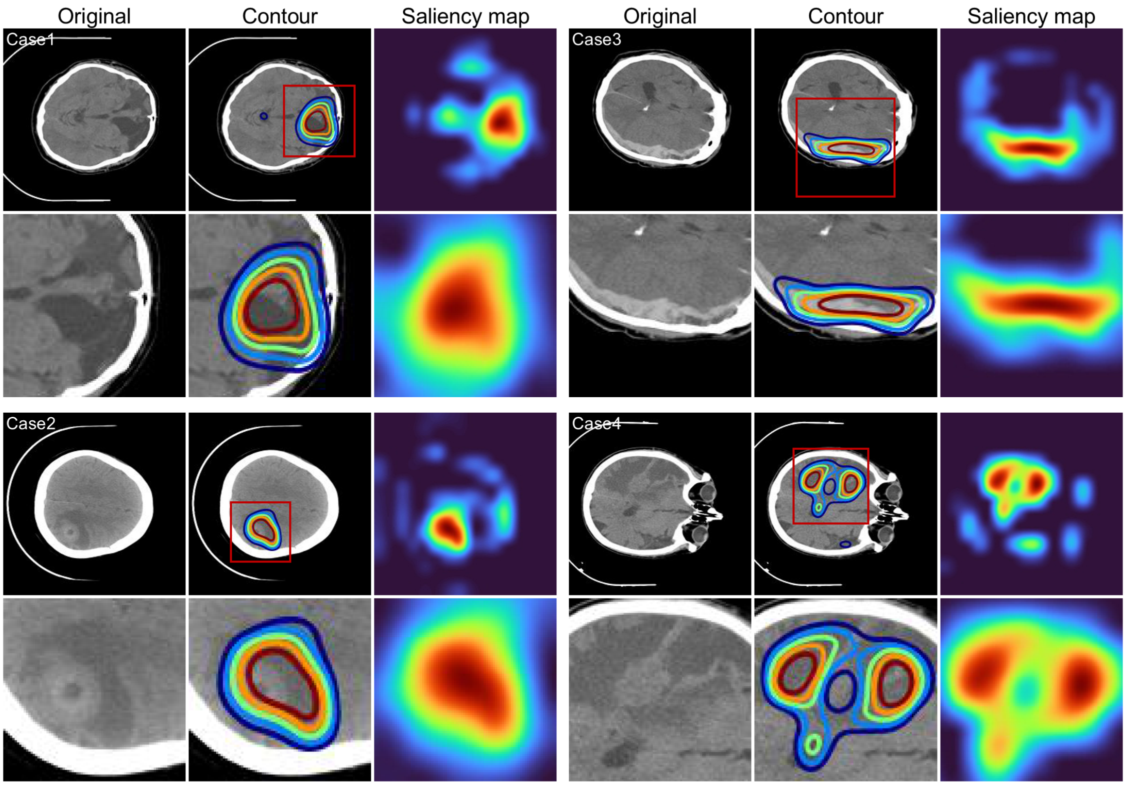


**Supplementary Figure 35**

**Saliency maps generated by Brainfound for zero-shot classification.** We showcased representative images of saliency maps for four brain CT images from Brainfound. The brain CT images are positioned in the first and fourth columns. The second and fifth columns show the saliency contours. The saliency maps developed by Brainfound are located in the third and sixth columns. The images in the second and fourth rows offer an enlarged perspective of the sections highlighted by red boxes in the first and third rows.

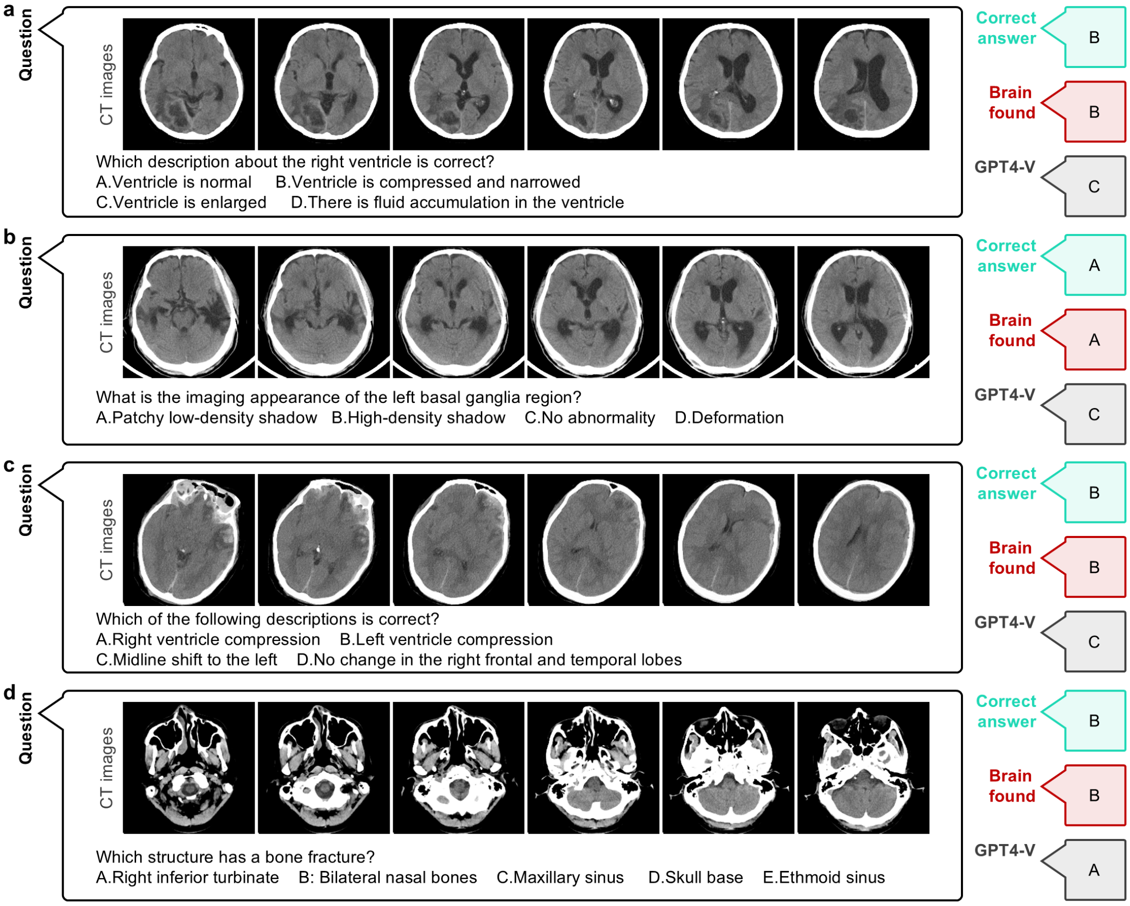


**Supplementary Figure 36**

**The responses of Brainfound on several multiple-choice questions about brain imaging, Part I**. **a**, With the CT images, Brainfound accurately identified the imaging characteristics of the left basal ganglia. **b**, Utilizing brain CT imaging, Brainfound determines which option is correct. **c**, Brainfound accurately selected the description of the right ventricle. **d**, Brainfound accurately identified the location of the fracture.

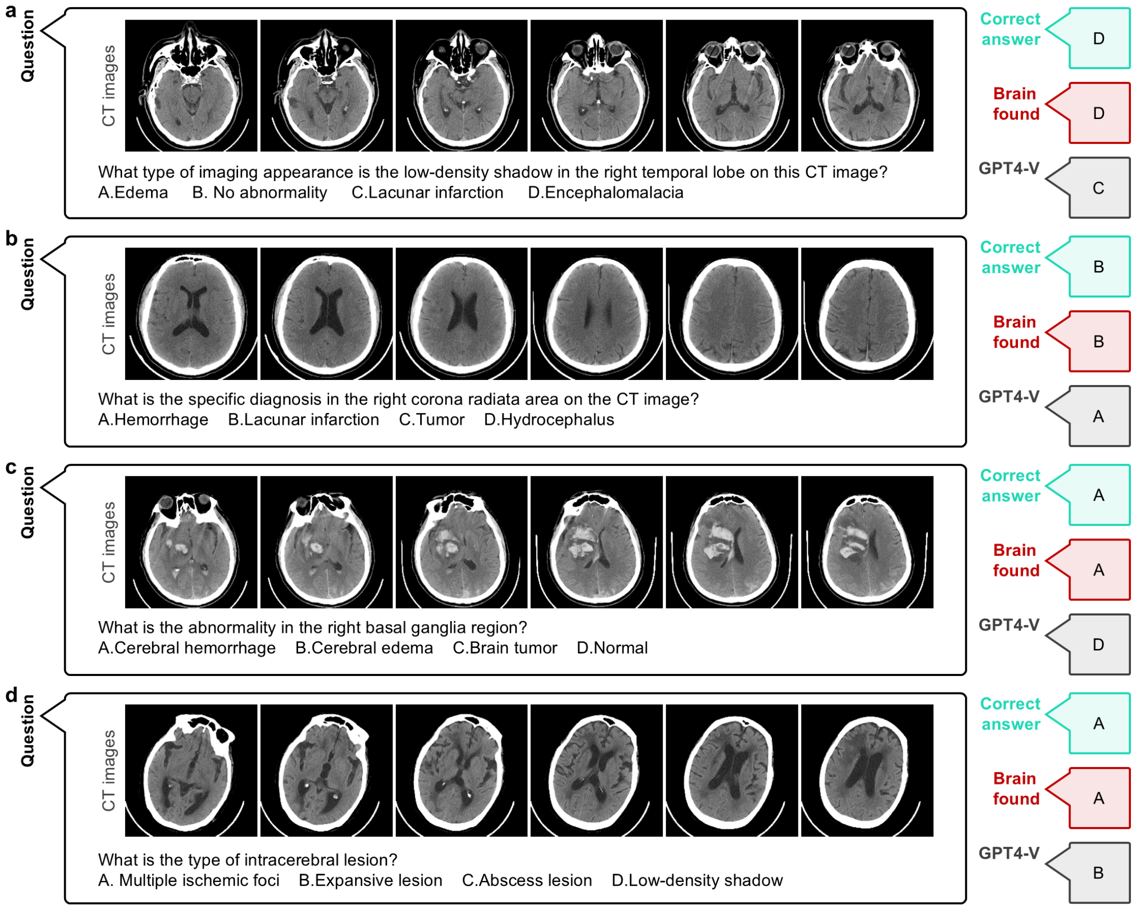


**Supplementary Figure 37**

**The responses of Brainfound on several multiple-choice questions about brain imaging, Part II**. **a**, Brainfound accurately determined the type of low-density shadow in the right temporal lobe depicted in the CT images. **b**, Brainfound accurately determined the specific diagnostic result for the right corona radiata area. **c**, Brainfound correctly identified the abnormality type in the right basal ganglia region. **d**, Brainfound accurately determined the type of brain lesion in the CT image.

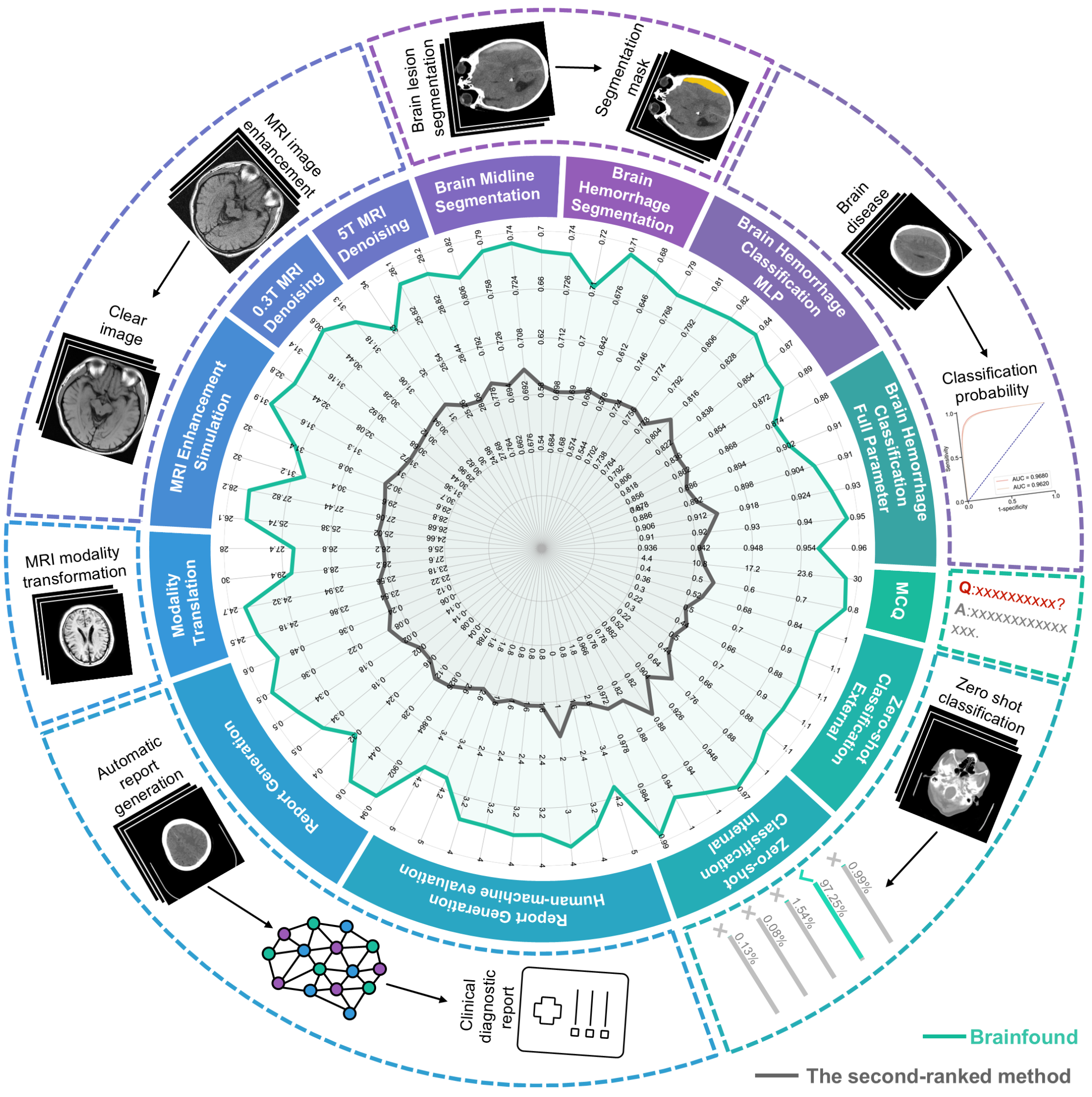


**Supplementary Figure 38**

**Schematic diagram summarizing the evaluation results of Brainfound.** The radar chart displays all the experimental results. Two curves are plotted, one representing the results of Brainfound and the other representing the results of the second-ranked method in all comparisons. The names of the task types are labeled on the outside of the radar chart.
